# COVID-19 Time-varying Reproduction Numbers Worldwide: An Empirical Analysis of Mandatory and Voluntary Social Distancing

**DOI:** 10.1101/2021.04.06.21255033

**Authors:** Alexander Chudik, M. Hashem Pesaran, Alessandro Rebucci

## Abstract

This paper estimates time-varying COVID-19 reproduction numbers worldwide solely based on the number of reported infected cases, allowing for under-reporting. Estimation is based on a moment condition that can be derived from an agent-based stochastic network model of COVID-19 transmission. The outcomes in terms of the reproduction number and the trajectory of per-capita cases through the end of 2020 are very diverse. The reproduction number depends on the transmission rate and the proportion of susceptible population, or the herd immunity effect. Changes in the transmission rate depend on changes in the behavior of the virus, re-flecting mutations and vaccinations, and changes in people’s behavior, reflecting voluntary or government mandated isolation. Over our sample period, neither mutation nor vaccination are major factors, so one can attribute variation in the transmission rate to variations in behavior. Evidence based on panel data models explaining transmission rates for nine European countries indicates that the diversity of outcomes resulted from the non-linear interaction of mandatory containment measures, voluntary precautionary isolation, and the economic incentives that gov-ernments provided to support isolation. These effects are precisely estimated and robust to various assumptions. As a result, countries with seemingly different social distancing policies achieved quite similar outcomes in terms of the reproduction number. These results imply that ignoring the voluntary component of social distancing could introduce an upward bias in the estimates of the effects of lock-downs and support policies on the transmission rates.

**JEL Classification:** D0, F6, C4, I120, E7

## 1 Introduction

The COVID-19 pandemic has claimed millions of lives and brought about very costly government interventions to contain it, with unprecedented and widespread economic disruption worldwide. China responded to the initial outbreak with strict and binding mandatory social distancing policies to contain the epidemic and it is widely credited to have been successful in eradicating the virus each time it resurfaced. At the other end of the spectrum, for example, Sweden initially attempted to let its epidemic run its course with only minimal interventions from the government. Other countries responded by adopting a mixture of policies, either by deliberate choice or due to popular opposition to the implementation of lock-downs or even milder forms of social distancing. Yet outcomes in terms of per capita cases and deaths do not always diverge or align with these priors.

The purpose of this paper is two-fold: (a) to compare time-varying estimates of COVID-19 effective reproduction numbers across a large number of diverse countries, and to highlight the wide disparities that exist across countries. (b) to model empirically the effects of mandatory and voluntary social distancing as well as incentives to comply on the evolution of the virus transmission rate in a sample of European countries.

Reproduction numbers are epidemiologic metrics to measure the intensity of an infectious disease. The basic reproduction number, denoted by *ℛ*_0_, is the number of new infections expected to result from one infected individual at the start of the epidemic. Within a classical susceptible-infective-removed (SIR) model the basic reproduction number is given by *ℛ*_0_ = *β*_0_*/γ*, where *β*_0_ is the initial (biological) transmission rate, and *γ* is the recovery rate. Since the transmissibility of a disease will vary over time due to changes in immunity, mitigation policies, or precautionary behavior, the effective reproduction number, which we denote by *ℛ*_*et*_, measures the *ℛ* number *t* periods after the initial outbreak. As we show in the paper, in the classical SIR model, we have *ℛ*_*et*_ = (1 − *c*_*t*_) *β*_*t*_*/γ*, where *c*_*t*_ is the per capita number of infected cases at time *t*. Moreover, in our framework, for a given value of *ℛ*_0_, social distancing, either voluntary due to precautionary behavior or mandatory due to non-pharmaceutical interventions (which we also call mitigation or containment policies for brevity), lead to time-variation in the transmission rate, *β*_*t*_, driven by changes in contacts or susceptibility to infection. As a result, in our model, one can separate changes in *ℛ*_*et*_ due the extent to which the susceptible population is shrinking, 1 − *c*_*t*_ (which we call herding), or due to social distancing whether mandated or voluntary.

To estimate the COVID-19 time-varying transmission rates, *β*_*t*_, we apply a new method proposed by Pesaran and Yang (2020), henceforth PY, based on a moment condition that can be derived from an agent-based stochastic network model of epidemic diffusion. As PY show, a linearized version of this moment condition can aggregate up to the classical deterministic SIR model. From this moment condition, we then derive reduced form regressions in confirmed cases that control for under-reporting due to unreported asymptotic cases and/or in absence of universal COVID-19 testing.

The estimation approach that we propose is applicable to any level of jurisdiction and could provide guidance on how to measure the health impact in causal studies of specific mitigating policies. For the sake of brevity we report the results for selected countries and regions, but compute rolling estimates of effective reproduction numbers for all jurisdictions for which Johns Hopkins University (JHU) reports case statistics.^1^ This is important since measuring health outcomes is challenging in studies that seek to establish causal effects of policies to address COVID-19.

Our method of moment estimation requires only data on infected cases, thus complementing estimation methods based on death statistics. The reported number of infected cases and deaths is problematic and different countries might have better quality data on either one or the other. For example, Spain has very good death statistics. In other countries, death statistics have undergone major revisions on several occasions. For example, the United Kingdom death toll was revised downward by 5,377 on August 12, 2020 after a review concluded that daily death figures should only include deaths which had occurred within 28 days of a positive COVID-19 test.

Our estimation method is not only simple to apply, but also fairly robust to the under-reporting of infected cases. Many existing estimation methods of reproduction numbers do not allow for measurement errors and might not be robust to the under-reporting problem. For instance, the seemingly unrelated regression estimates developed by Korolev (2020) may be biased downward if one neglects under-reporting of confirmed cases. Stock (2020) focuses on measurement errors and explores the benefits of randomly testing the general population to determine the asymptomatic infection rate.

Following the medical evidence in Gibbons et al. (2014), we use a multiplication factor to allow for under-reporting in a way that will be elaborated in later sections of the paper. This is particularly important given that it has been widely acknowledged that number of reported infected cases may suffer from considerable under-reporting, especially during the early stages of the epidemic. For example, Li et al. (2020) estimate that only 14 percent of all infections were documented in China prior to the January 23, 2020 travel restrictions. This translates to a multiplication factor of 1*/*0.14 *≈* 7.14. Jagodnik et al. (2020) estimate that the recorded cases were under-reported by a multiplication factor in the range of 3 to 16 times in the seven countries that they considered as of March 28, 2020.^2^ According to the Centers for Disease Control and Prevention study of Havers et al. (2020), in the United States, the number of infected cases is likely to be 10 times more than reported based on antibody tests from March through May, 2020. More recently, Rahmandad, Lim, and Sterman (2020) estimate that the cumulative cases across 86 countries through July 10, 2020 are 10.5 times the number of officially reported cases, with a 10^*th*^ − 90^*th*^ percentile range of 3.35 − 23.81.

We find that for China the effective reproduction number drops below 1 within 30 days of the lock-downs. The reproduction number estimates obtained for other countries initially also fall, as found for instance by Atkeson, Kopecky, and Zha (2020a). Contrary to evidence based on COVID death statistics, however, the pace and the magnitude of the decline varies significantly across countries. For example, we estimate it took about three times longer for the southern hemisphere to bring the reproduction number down to one compared with the northern hemisphere, excluding China.

The critical difference between China and all other countries is not only the initial slower decline of the reproduction number, but also the fact that, with very few exceptions, the epidemic was never eradicated completely and hence it resurfaced every time restrictions were loosened. This is important because without herd immunity, community transmission resumes as soon as social interaction resumes. Indeed, our estimates show that, in most cases, the estimated reproduction number does not remain permanently below one and most countries experience more than one wave, with the second wave typically larger than the first one.

The paper also finds that, whilst in China and Asia the reproduction number is driven entirely by a reduction in contacts and vulnerability to COVID-19, in the United States, the United Kingdom, Brazil, and several other countries, community transmission drops also due to the rise in the proportion of population becoming infected, and the rise in the relative importance of herd immunity. Critically, our results show that countries with seemingly different social distancing policies achieved quite similar outcomes in terms of the reproduction numbers.

For a better understanding of the factors behind the evolution of effective reproduction numbers, we separate the herding component from the *transmission rate* and empirically model the latter for a sample of European countries with a similar start to the outbreaks in March 2020, but with differing outcomes subsequently. In line with the agent-based stochastic epidemic network model of PY, we argue that it is the transmission rate, and not the *ℛ* number, that depends on behavioral changes. The agent-based model shows the transmission rate to depend on the average number contacts *multiplied* by individual-specific susceptibility to become infected which in turn depends on average duration of contacts, wearing of face masks, and other recommended precautions. Accordingly, in our empirical analysis we assume that a country’s time-varying country-specific transmission rate depends on three factors. Consistent with a simple decision theoretic model presented in the paper and a large body of empirical evidence, we distinguish between government-mandated social distancing policies and voluntary self-isolation. We also control for government economic support that affects the incentive to comply as reported in survey data (e.g., Papageorge et al., 2021; Hamermesh, 2020). To measure mandated-social distancing and incentives to comply with these policies we use the stringency and support indices compiled by Oxford COVID-19 Government Response Tracker project.^3^ To assess the impact of voluntary social distancing, we allow for a threshold effect capturing the impact of fear of becoming infected arising from news of rising cases on individual precautionary behavior. The importance of these factors in controlling the effective rate of transmission is jointly estimated within the context of the epidemic model, allowing for a lag of two or three weeks between the policy or behavioral changes and the infection outcomes.

We find that all three determinants of the transmission rate are statistically highly significant and have the expected signs. However, consistent with the heterogeneity documented in the first part of the empirical analysis, we also find that when we control for voluntary behavior by adding the threshold effect, the magnitudes of estimated coefficients on stringency and compliance indicators decline markedly. This is clear evidence suggesting that the role of mandatory policies might be overestimated in studies that do not explicitly allow for voluntary self-isolation and herding. In addition, our estimates suggest that voluntary social distancing alone would not have been sufficient to bring the *ℛ* number below one in Europe and to keep it there without substantial contributions from herd immunity and/or mass vaccination.

To summarize the main message, our empirical analysis shows that mandatory social distancing is critical, as voluntary social distancing alone does not seem capable of bringing the reproduction number below one. Draconian mandatory social distancing, as in the case of China, can succeed. However, in light of the economic and social costs of such an approach, other countries attempted to pursue alternative strategies. As a result, epidemic curves show a great deal of heterogeneity. They show that protracted, albeit not full, China-style lock-downs alone are not sufficient to contain the epidemic, although they are a necessary ingredient of a policy response aimed at bringing the effective reproduction number below one. Similarly, it does not seem plausible for voluntary social distancing to do the job on its own, without relying on herd immunity and/or mass vaccination.

### Related Literature

A very large body of research investigates the COVID-19 outbreak and the policies to contain its spread.^4^ For example, Fang, Wang, and Yang (2020) analyze efforts to contain the COVID-19 outbreak in China, measuring the effectiveness of the lock-down of Wuhan and showing that these policies also contributed significantly to reducing the total number of infections also outside of Wuhan. Similarly, there is ample reduced form evidence on the impact of mandatory social distancing using state and county level data in the case of the United States, and for a few other countries. However, there are not many studies on the relative importance of mandatory and voluntary social distancing, especially for large cross sections of countries.

Caselli et al. (2020) find that both lock-downs and voluntary social distancing helped contain the first wave of COVID-19, but mandatory interventions have been critical. Jinjarak et al. (2020) find that more stringent policies are associated with lower mortality growth rates in a large cross section of countries but with some heterogeneity depending on demographics, the degree of urbanization and political freedom, as well as the international travel flows. In general, however, countries with more stringent policies at the onset of the epidemic realized lower peak mortality rates and exhibited lower duration during the first epidemic wave. We distinguish not only between mandatory and voluntary social distancing, but also consider the role of herd immunity in lowering the reproduction number. To our knowledge, no study which considers voluntary or government-mandated social distancing also controls for the possibility of herd immunity and distinguishes its impacts on effective reproduction numbers from the influence of policy and/or behavioral factors.

A number of studies consider the effects of different intervention strategies – such as isolating the elderly, closing schools and/or workplaces, and alternating work/school schedules – which should lower the average number of contacts of specific age groups, contact locations, or time windows relative to normal (pre-COVID) patterns using calibrated behavioral SIR or compartmental models.^5^ We take an empirical/econometrics approach calibrating only the recovery rate, *γ*; a parameter on which we have much more precise clinical information.

Various methods are available in the epidemiological literature to estimate the reproduction numbers at the beginning and/or in real time during epidemics, but there is no uniform framework. Estimation approaches that are data-driven and which involve simplifying assumptions include the use of the number of susceptibles at endemic equilibrium, the average age at infection, the final size equation, and calculation from the intrinsic growth rate of the number of infections (Heffernan, Smith, and Wahl 2005). Estimation of reproduction numbers based on different models are reviewed by Chowell and Nishiura (2008), Obadia, Haneef, and Boëlle (2012), and Nikbakht et al. (2019). More recent contributions, focusing on estimation of reproduction numbers for the COVID-19 pandemic based on death statistics include Atkeson, Kopecky, and Zha (2020b), Baqaee et al. (2020), Korolev (2020) and Toda (2020).

Three closely related papers are Fernández-Villaverde and Jones (2020), Atkeson, Kopecky, and Zha (2020a), and Cakmakli and Simsek (2020). Fernández-Villaverde and Jones (2020) estimate transmission rates (*β*_*t*_*/γ*) for many jurisdictions as we do based on the number of observed deaths to infer the number of infections. The rationale is that confirmed infections are subject to significant measurement errors due to limited testing. Our estimates require only case data and correct for measurement errors due to under-reporting associated with testing capacity. Atkeson, Kopecky, and Zha (2020a) report a set of stylized facts on death rates for a large cross section of countries and conclude that mortality falls rather uniformly across countries within the first 20-30 days after the first 25 cumulative deaths and remain low through the summer of 2020. As the paper notes, this implies that both the effective reproduction numbers and the transmission rates of COVID-19 fall uniformly from high and heterogeneous initial levels and remain relatively low thereafter. We document much more heterogeneity during both the first and subsequent waves. Cakmakli and Simsek (2020) estimate a Susceptible-Infected-Recovery-Death (SIRD) model allowing for unreported cases as we do for six large countries. But as noted above, our method of moment estimation only uses the number of infected cases and does not use data on removed (recoveries plus deaths) that are particularity unreliable.

The rest of the paper is organized as follows. Section 2 discusses our SIR model with social distancing. Section 3 discusses the econometric estimation based on a moment condition derived from an agent-based model. Section 4 reports our estimates of the reproduction number for a selection of key countries and regions worldwide. Section 5 analyzes the relative importance of mandatory and voluntary social distancing. Section 6 concludes. An online appendix reports technical details and supplemental results.

## 2 SIR model with social distancing

There are many approaches to modelling the spread of epidemics. The basic mathematical model widely used by researchers is the susceptible-infective-removed (SIR) model advanced by Kermack and McKendrick (1927). This model and its various extensions have been the subject of a vast number of studies, and have been applied extensively to investigate the spread of COVID-19. A comprehensive treatment is provided by Diekmann and Heesterbeek (2000) with further contributions by Metz (1978), Satsuma et al. (2004), Harko et al. (2014), Salje et al. (2016), amongst many others.

The basic SIR model considers a given population of fixed size *n*, composed of three distinct groups, those individuals in period *t* who have not yet contracted the disease and are therefore susceptible, denoted by *S*_*t*_; the ‘removed’ individuals who can no longer contract the disease, consisting of recovered and deceased, denoted by *R*_*t*_; and those who remain infected at time *t* and denoted by *I*_*t*_. Thus,

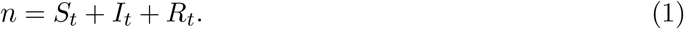

As it stands, this is an accounting identity, and it is therefore sufficient to model two of the three variables (*S*_*t*_, *I*_*t*_, and *R*_*t*_) to obtain the third as the remainder.

The classic SIR model is deterministic. It is cast in the following set of difference equations (for *t* = 1, 2,*…, T*)

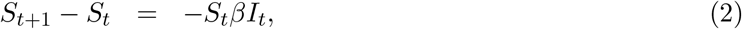

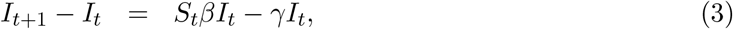

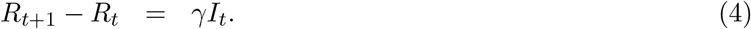

The parameter *β* is the rate of transmission, while *γ* is the recovery rate. For a given non-zero initial values *S*_1_ and *I*_1_, and the parameter values for *β* and *γ*, the evolution of the number of infected and recovered individuals is deterministic and given by the recursive solution of (2)-(4) for given initial values.

The evolution of the epidemic crucially depends on the two key parameters *β* and *γ*. It is easy to see from equation (2)-(4) that, without any mitigating intervention, the epidemic will spread if *β/γ* = *ℛ*_0_ *>* 1 and will cease only after infecting (*ℛ*_0_ − 1)*/ℛ*_0_ of the population. The parameter ratio *β/γ* = *ℛ*_0_ *>* 1 is referred to as the basic reproduction number, also defined in a stochastic context as the expected number of secondary cases produced by a single infected individual in a *completely* susceptible population. The terminal condition (*ℛ*_0_ − 1)*/ℛ*_0_ is the herd immunity threshold. In the case of COVID-19, a number of different estimates have been suggested in the literature, placing *ℛ*_0_ somewhere in the range of 2.4 to 3.9.^6^ So, the classical SIR model predicts that in the absence of intervention as much as 2/3 of the population could eventually become infected before herd immunity is reached.

Such an outcome would involve unbearable strain on national health care systems and a significant loss of life. This well understood possibility triggered unparalleled mitigation and containment interventions, first by China and South Korea, then Europe, the US and all other countries around the world. Such interventions, which broadly speaking we refer to as “social distancing” include case isolation, mandated face mask wearing, banning of gatherings, closures of schools and universities, and even local and national lock-downs; all aimed at slowing down the transmission rate of the virus. It is clear that these policies, together with voluntary changes in behavior in response to the epidemic, make it harder for the virus to transmit between individuals. One way to capture the impact of social distancing in the above model is to allow the transmission rate parameter, *β*, to be time-varying. In the remainder of the paper we will treat *β* as time-varying, whilst we assume the recovery rate *γ* is time invariant based on clinical evidence discussed below. We will refer to *β*_*t*_*/γ* as the “effective transmission rate”.

Following Pesaran and Yang (2020), hereafter PY, the transmission rate *β* and its time-varying version *β*_*t*_ can be represented by *β*_*t*_ = *τ*_*t*_*κ*_*t*_, where *τ*_*t*_ is the individual vulnerability to infection given contact (or exposure intensity) and *κ*_*t*_ is the average contacts per day. Social distancing, be it mandated and/or voluntary, can clearly influence both *τ*_*t*_ and/or *κ*_*t*_ and will result in time variation in the transmission rate.

PY also show that the classic aggregate SIR model (2)-(4) with time-varying transmission rate can be obtained as an approximation (for a large population *n*) to an individual-based stochastic network model of epidemic, where individuals randomly interact with each other. Stochastic simulation results obtained by PY also show that a single group model provides a good approximation to a multi-group alternative. In PY’s setting, the effective reproduction number is given by

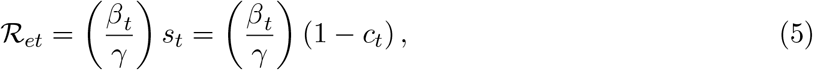

where *β*_*t*_*/γ* is the effective transmission rate, *c*_*t*_ = (*n* − *S*_*t*_)*/n* = 1 − *s*_*t*_ is the fraction of population that have been infected (cumulation of new infected cases), and 1 − *c*_*t*_ is the herd-immunity component of *ℛ*_*et*_. In this setting, *ℛ*_*et*_ is the expected number of secondary cases produced by one infected individual in a population that includes both susceptible and non-susceptible individuals at time *t*. It is also worth bearing in mind that at the outset of epidemic outbreak, assuming a fully susceptible population, we have *s*_0_ = 1 (*c*_0_ = 0), which in turn ensures that *ℛ*_*e*0_ = *β*_0_*/γ* = *ℛ*_0_.

As the epidemic evolves, the average number of secondary cases caused by a single infected individual will vary over time as a result of decline in the number of susceptible individuals (due to immunity or death) and/or changes in behavior due to social distancing. In our empirical analysis, we provide country-specific estimates of *ℛ*_*et*_ together with the effective transmission rate *β*_*t*_*/γ* which permits assessing the influence of herding. In the last section of the paper, we estimate the key determinants of *β*_*t*_ across a number of European countries. Note that the herding component of *ℛ*_*et*_, namely 1 − *c*_*t*_, is given and can not be affected by changes in mitigation policies.

## 3 Estimating time-varying transmission rates

In order to estimate country-specific time-varying transmission rates, *β*_*t*_, we utilize the following aggregate (non-linear) moment condition derived from the agent-based stochastic model of PY:

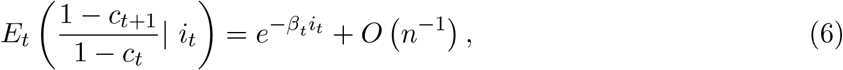

where *i*_*t*_ = *I*_*t*_*/n* is the number of infected individuals (scaled by population), and the last term is the approximation error in *n*, which we ignore since *n* is large. Here, it is important to note that (in large populations as in our data) the transmission rate can change only if the biology of virus changes (mutations), a vaccine is introduced, or people change behaviors voluntarily or due to policy interventions. Transmission rate *β*_*t*_ in (6) reflects all these factors. However, for most of our sample, virus mutations are likely to be of secondary importance perhaps with the exception of the last few weeks. Similarly, vaccinations did not start until late 2020 or early 2021 on all countries, with only the United Kingdon and Israel making some progress in January 2021. Hence, we attribute the change in the transmission rate in 2020 primarily to the change in behaviour. In other words, under the identification assumptions of constant biology of the virus and no vaccinations, time variations in *β*_*t*_ are caused by interventions alone. In contrast, in January 2021 and in the subsequent months, global roll-out of inoculations can be a gradually more important additional factor in reducing the transmission rate. Moreover, mutations might also play a more significant role in the moths ahead.

In estimating *β*_*t*_ we face two data-related difficulties. The first difficulty is in obtaining data on active cases, *I*_*t*_ = *C*_*t*_ − *R*_*t*_, where *R*_*t*_ is the number of recoveries and/or dead. This is not straight-forward because data on *R*_*t*_ either do not exist or are unreliable due to considerable measurement difficulties. Consider for example Europe. The recorded data on recoveries are unavailable for Spain and UK; they are of poor quality for France and Italy; and they are relatively close to our estimated recovery for Austria and Germany. To overcome this problem, we use the SIR model’s recovery equation to impute data on recoveries based on the confirmed cases *alone*, assuming a recovery rate *γ* = 1*/*14. We obtain very similar results if we use *γ* = 1*/*21.^7^ The choice *γ* = 1*/*14 is consistent with the assumptions made in designing quarantine policies based on clinical evidence and also used in calibrated behavioral epidemic models.^8^

The second difficulty is with the measurement of confirmed cases, which are likely to be under-reported, in part due to fact that a non-negligible portion (perhaps about a half) of the cases is asymptomatic and therefore unlikely to be detected without large-scale testing. To mitigate the problem of under-reporting, we follow the epidemiological literature (see, for example, Gibbons et al., 2014) and assume that the magnitude of under-reporting is measured by the multiplication factor (MF - the ratio of true cases to reported cases). Denoting the observed values of *c*_*t*_ and *i*_*t*_ by 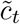 and *ĩ*_*t*_, we have

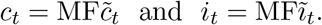

Then the moment condition in terms of observed values (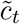 and *ĩ*_*t*_) can be written as

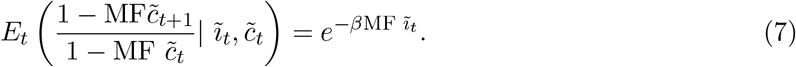

We do not know the true *MF*. We allow for the multiplication factor to be larger than one. We abstract from time variation in *MF*, and note that the magnitude of *MF* will not matter when only a relatively small fraction of the population has been infected as is true currently for COVID-19 worldwide. Estimation results in PY suggest a *MF* value of about 5 for most countries, declining slowly to about 2.5 towards the end of the sample. In figures reported below we use *MF* = 3 (a conservative value for the end of the sample, where herd immunity plays the most important role). In the online Appendix, we compare these results with the ones obtained for *MF* = 5. Estimates of the reproduction numbers are not sensitive to the choice of *MF*, whereas estimates of the effective transmission rate can be sensitive to the choice of *MF* towards the end of the sample in those countries where the reported share of infected population is relatively large. In our panel estimation results for a sample of European countries in Section 5, we report estimates for *MF* = 3 and 5, but the panel estimation results are very similar also for *MF* = 2 and 7.^9^

One additional challenge is that the reported daily data are subject to weekly distortions (e.g., the reported number of cases on Sundays is usually lower compared with the infected cases reported for other days). To deal with this calendar distortion, as is common practice, we take seven-day moving averages of the reported data used in estimation. But again we note that our results are robust if we use reported daily cases without averaging. Using the moment condition (7), we compute rolling-window estimates of the transmission rate as

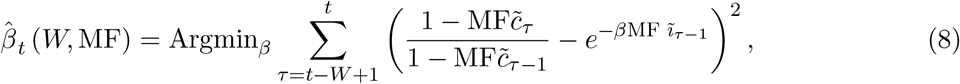

where *W* is the rolling window size, which we set to 14 days.

To estimate *β*_*t*_ following (8), we need observations on per capita infected and active cases, *c*_*t*_ and *i*_*t*_. Using the recorded number of infected cases, *C*_*t*_, and population data, *c*_*t*_ is readily available.^10^ Since *I*_*t*_ = *C*_*t*_ − *R*_*t*_, we can obtain *i*_*t*_ if the number of removed (recovered) cases, *R*_*t*_, is available. For all countries we estimate the number of removed (including recoveries and deaths) by *R*_*t*_ = (1 − *γ*) *R*_*t*−1_ + *γC*_*t*−1_, for *t* = 2, 3,*…*., where the recovery rate *γ* is set to 1*/*14, and the process starts from *R*_1_ = 0, and reported *C*_1_. We then compute *I*_*t*_ by subtracting the estimated *R*_*t*_ from the recorded *C*_*t*_.

## 4 COVID-19: A global pandemic with heterogeneous time-varying transmission

We are now ready to report our model estimates. In this section we report country-specific estimates of the reproduction number, *ℛ*_*et*_. We refer to it below as simply the “*ℛ* number”. We plot it alongside the effective transmission rate, *β*_*t*_ *×* 14 = *β*_*t*_*/γ*, to separately assess the influence of herding from social distancing, for a large sample of countries. In the next section of the paper, we will identify and estimate factors that contribute to the evolution of *β*_*t*_*/γ*, distinguishing between voluntary and mandatory social distancing for a selected group of European countries that have experienced similar starts to the outbreaks in March 2020, but with differing outcomes.

While we estimate the two parameters of interest for all jurisdictions for which JHU reports case statistics, in this section we report only the results for selected countries and regions.^11^ Figures 1-8 plot the results. Figure 1 reports results for China and the rest of the world. Figures 2-4 show results by geographic regions (excluding China): the Northern and Southern Hemispheres (Figure 2) and all main regions of the world (Figures 3-4), including East Asia and Pacific, South Asia, Eastern Europe and Central Asia, Western Europe, North America, Latin America and Caribbean, Middle East and North Africa, and Sub-Saharan Africa. Figures 5-6 report results for selected large economies. Finally, Figures 7-8 give results for the selected European economies also analyzed in Section 5 below. The estimates of transmission rates and *ℛ*-numbers for the regions are based on aggregate region-specific case statistics rather than averages across country specific estimates.

**Figure 1:**
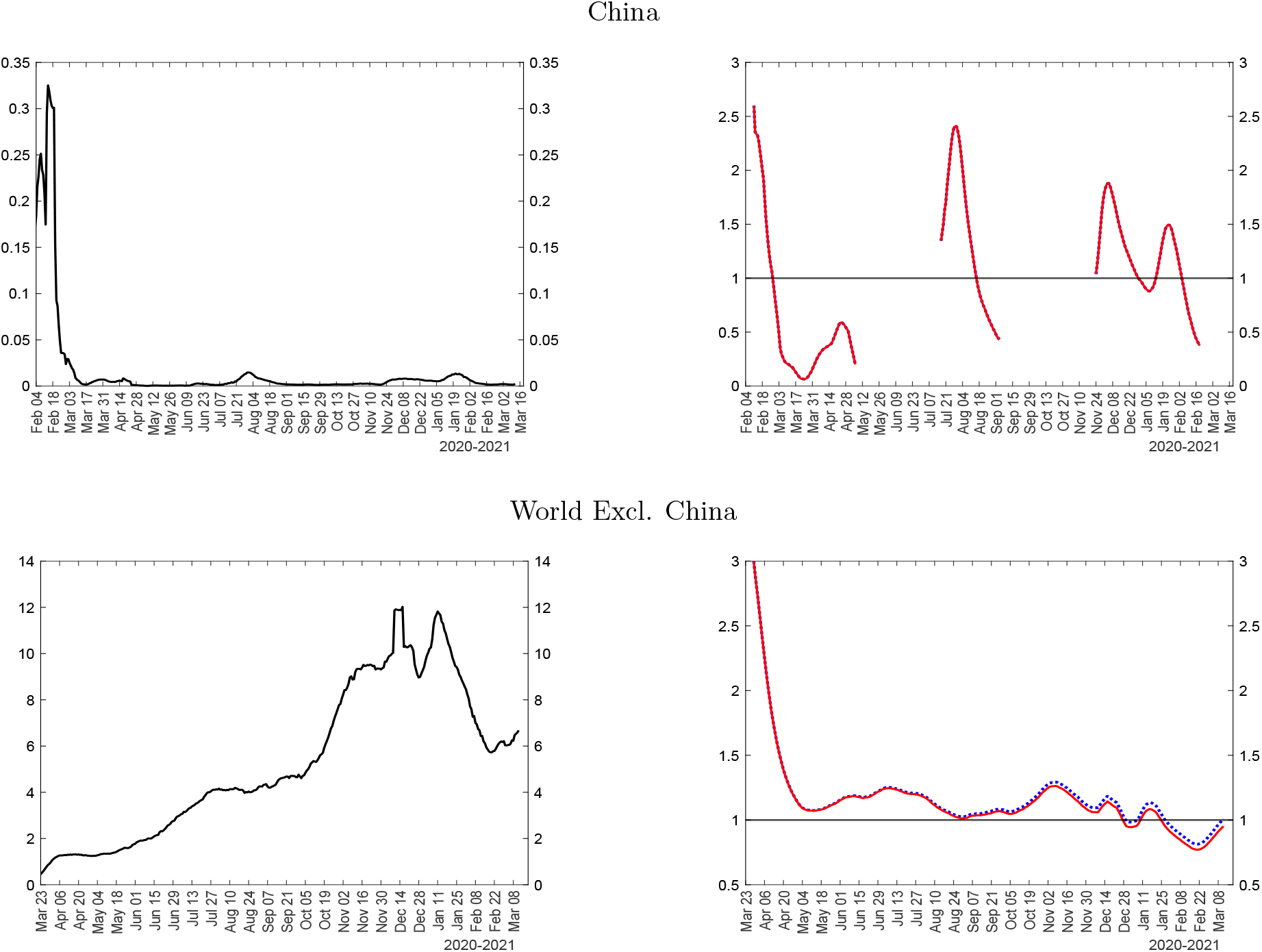
New cases (left) and *ℛ* numbers (right) for China and the rest of the world Notes: The figure plots a seven-day moving average of the number of reported new cases per 100k population (left charts), the *ℛ* number, 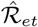 (right charts, solid red line), and the effective transmission rate, 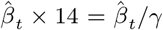 (right charts, dotted blue line). 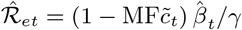, where *γ* = 1*/*14, and MF=3 for each country. 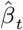 is estimated using (8), where the number of active infections is computed using the data on confirmed cases minus imputed removed cases. The number of removed (recoveries + deaths) is imputed recursively using *R*_*t*_ = (1 − *γ*) *R*_*t*−1_ + *γC*_*t*−1_ for all countries.

**Figure 2:**
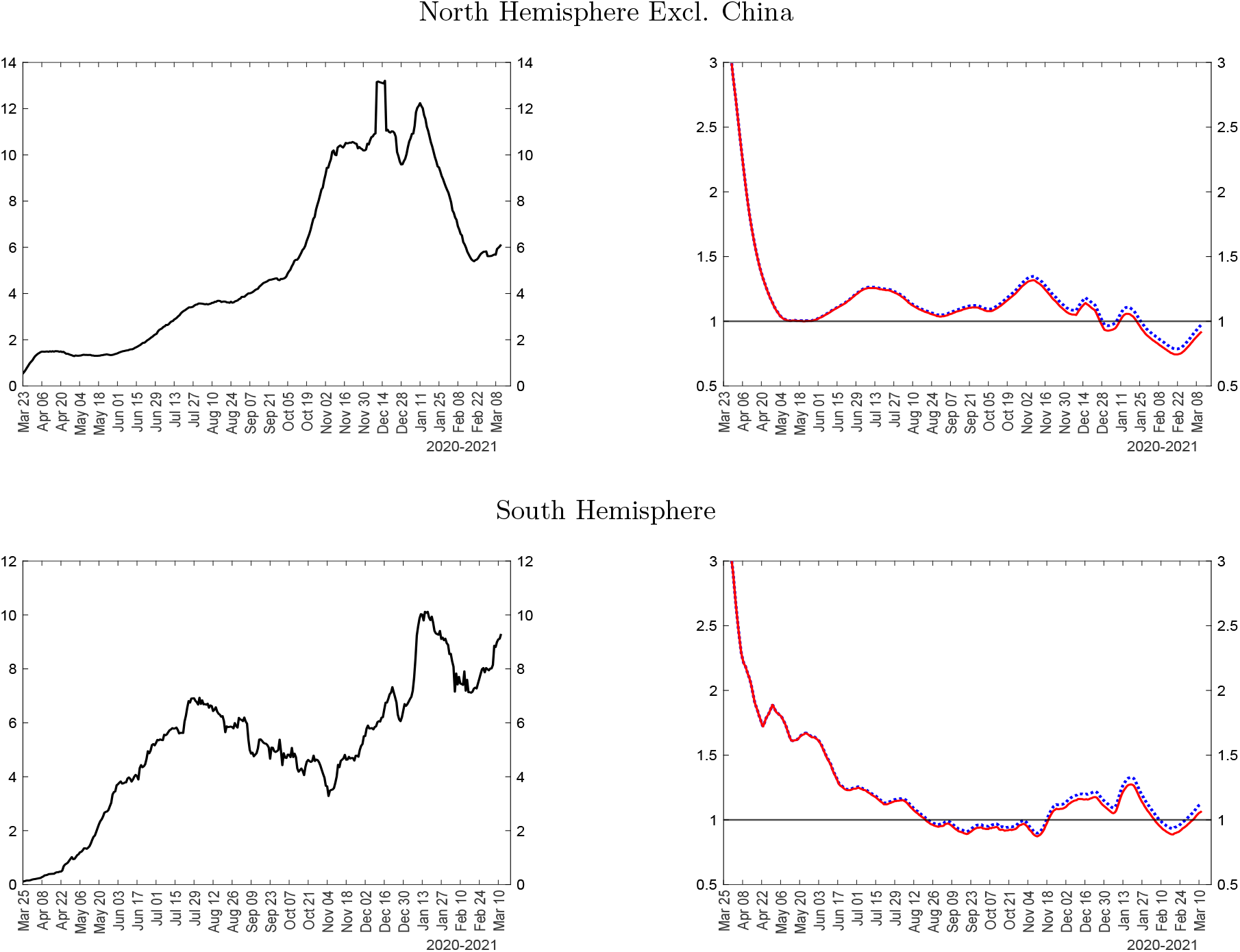
New cases (left) and *ℛ* numbers (right) for North and South Hemispheres (excl. China). Notes: The figure plots a seven-day moving average of the number of reported new cases per 100k population (left charts), the *ℛ* number, 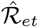 (right charts, solid red line), and the effective transmission rate, 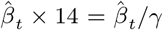 (right charts, dotted blue line). See notes to Figure 1.

**Figure 3:**
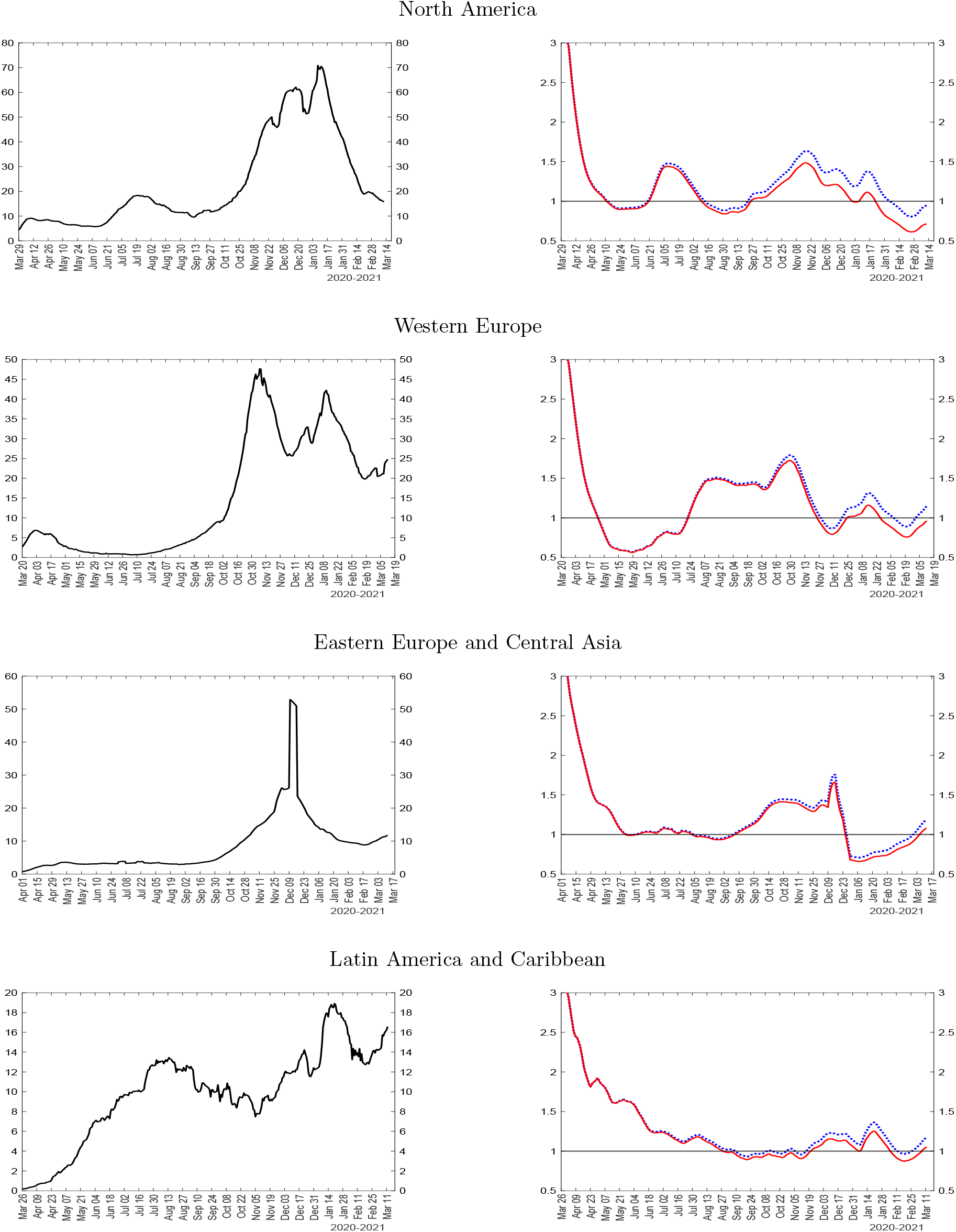
New cases (left) and *ℛ* numbers (right) for selected geographic regions Notes: The figure plots a seven-day moving average of the number of reported new cases per 100k population (left charts), the *ℛ* number, 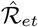 (right charts, solid red line), and the effective transmission rate, 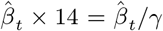 (right charts, dotted blue line). See notes to Figure 1.

**Figure 4:**
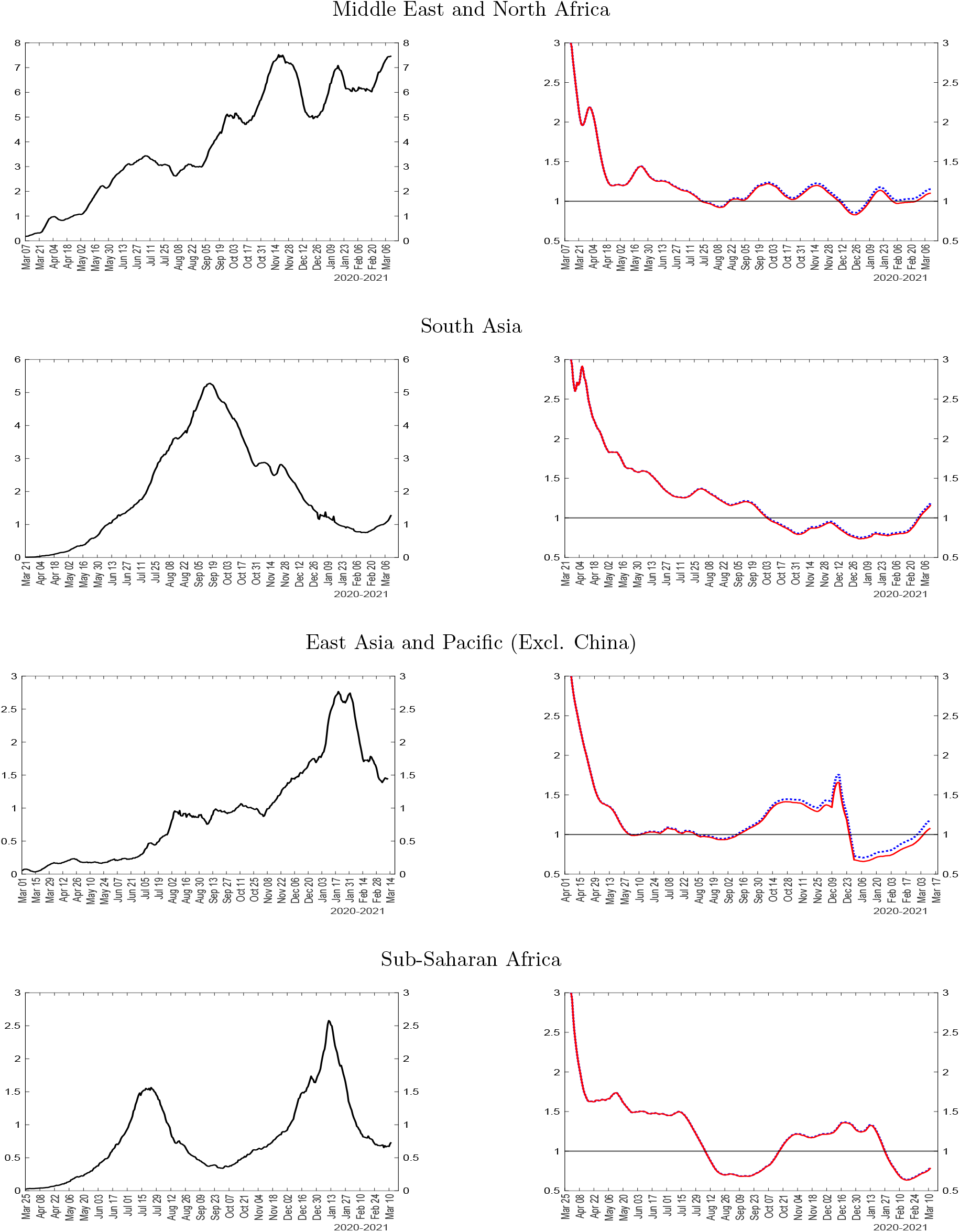
New cases (left) and *ℛ* numbers (right) for selected geographic regions Notes: The figure plots a seven-day moving average of the number of reported new cases per 100k population (left charts), the *ℛ* number, 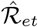 (right charts, solid red line), and the effective transmission rate, 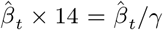 (right charts, dotted blue line). See notes to Figure 1.

**Figure 5:**
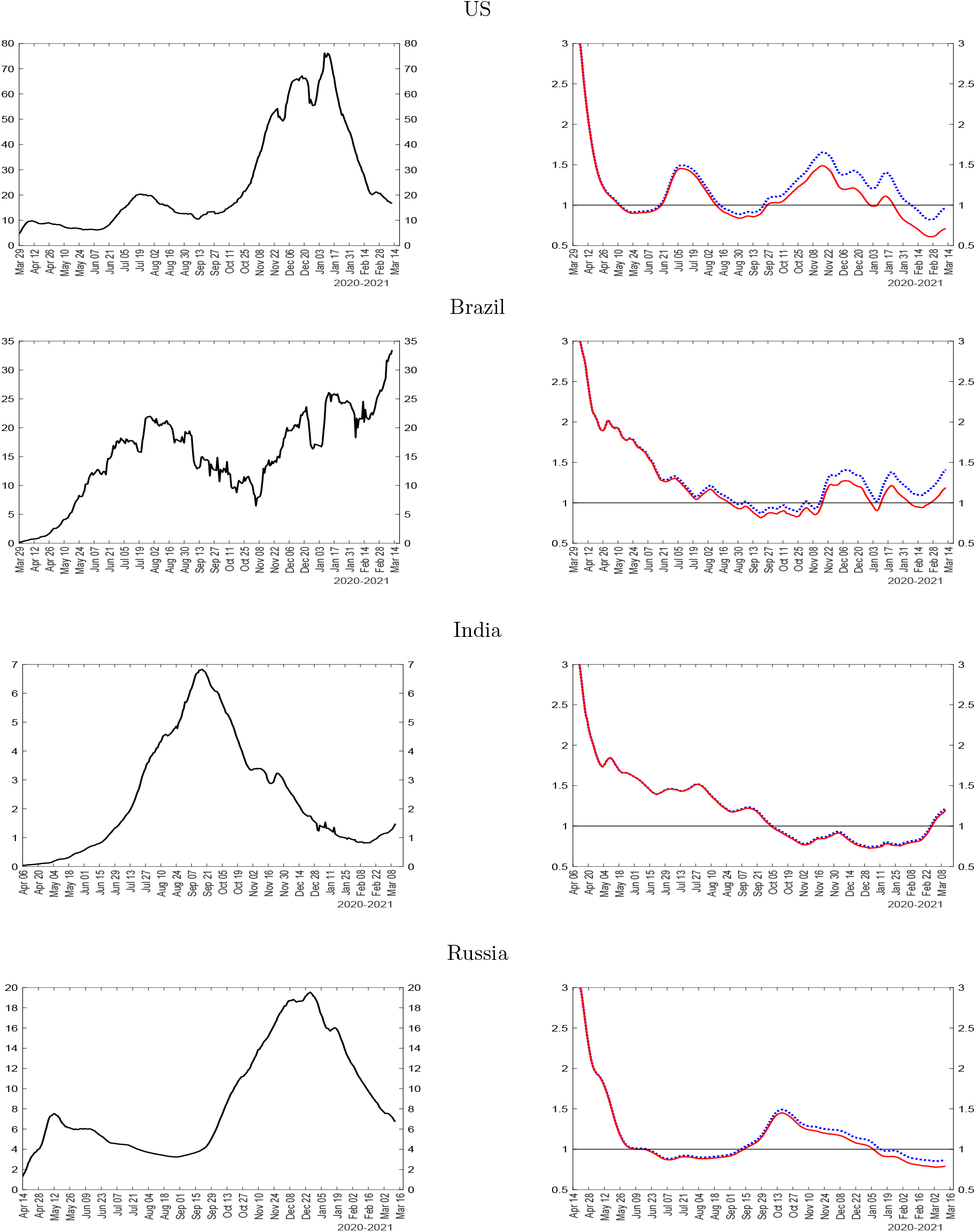
New cases (left) and *ℛ* numbers (right) for selected countries Notes: The figure plots a seven-day moving average of the number of reported new cases per 100k population (left charts), the *ℛ* number, 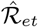 (right charts, solid red line), and the effective transmission rate, 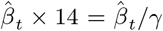 (right charts, dotted blue line). See notes to Figure 1.

**Figure 6:**
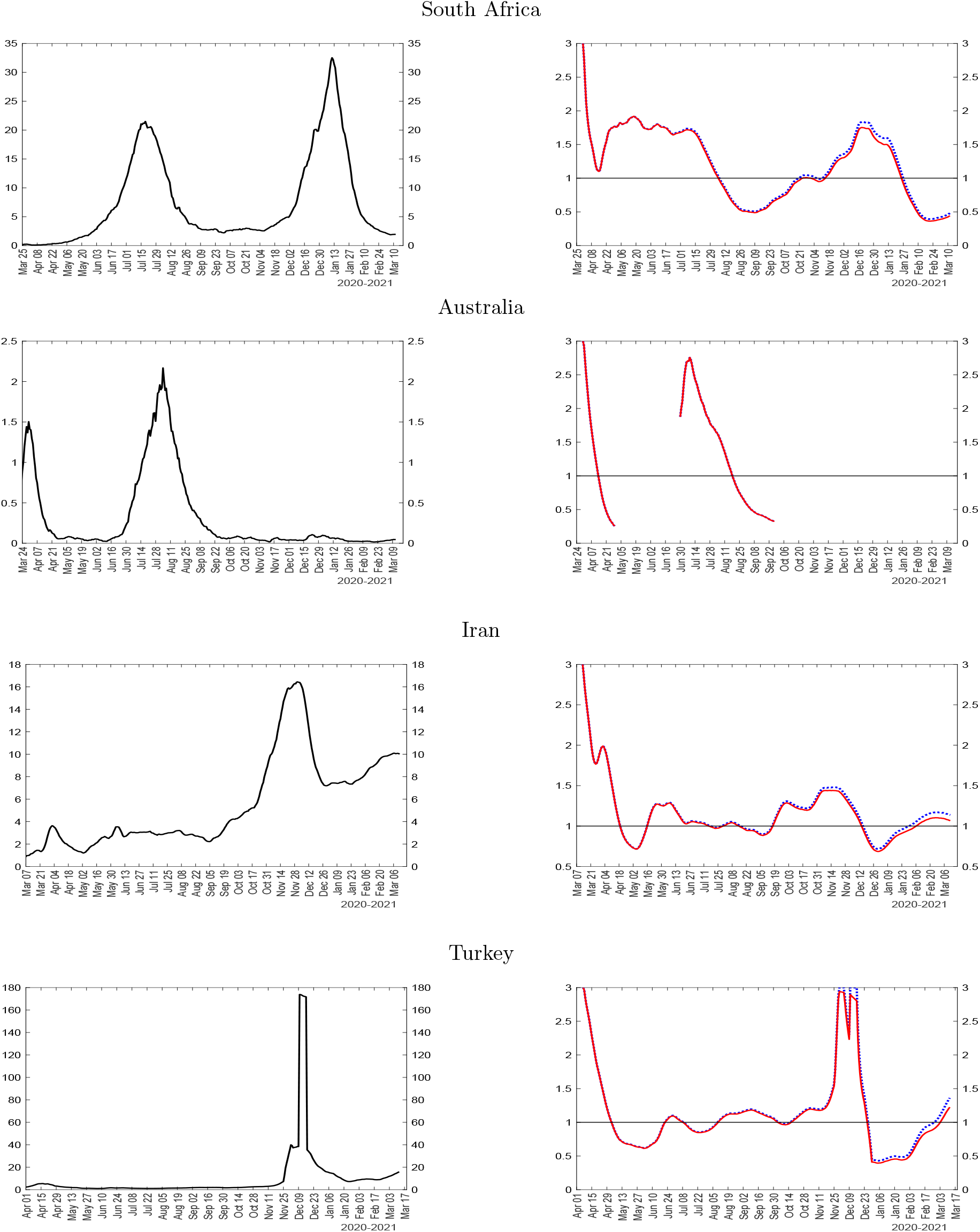
New cases (left) and *ℛ* numbers (right) for selected countries Notes: The figure plots a seven-day moving average of the number of reported new cases per 100k population (left charts), the *ℛ* number, 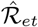 (right charts, solid red line), and the effective transmission rate, 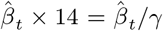 (right charts, dotted blue line). See notes to Figure 1.

**Figure 7:**
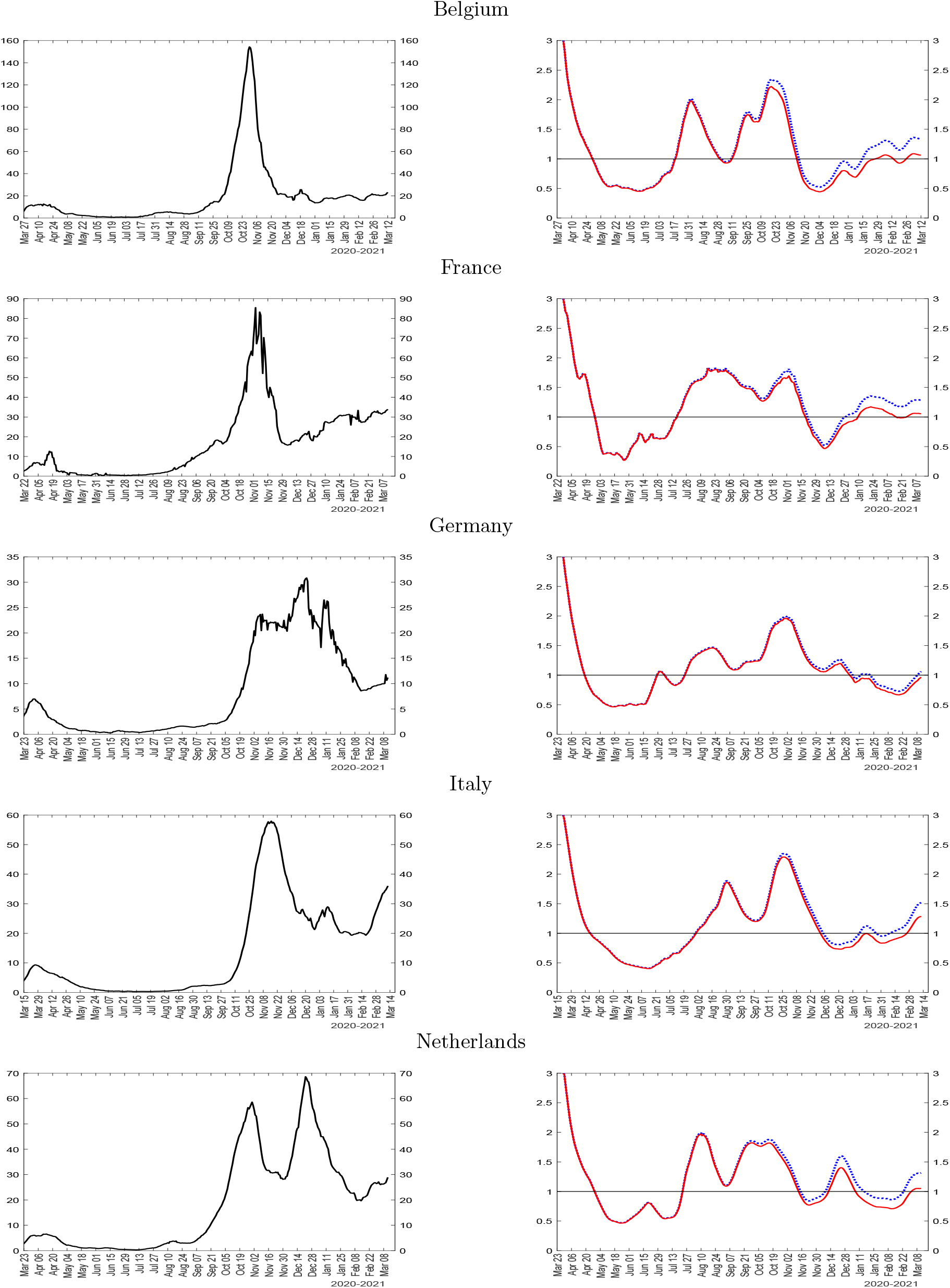
New cases (left) and *ℛ* numbers (right) for sample of European countries Notes: The figure plots a seven-day moving average of the number of reported new cases per 100k population (left charts), the *ℛ* number, 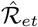 (right charts, solid red line), and the effective transmission rate, 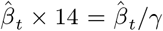 (right charts, dotted blue line). See notes to Figure 1.

**Figure 8:**
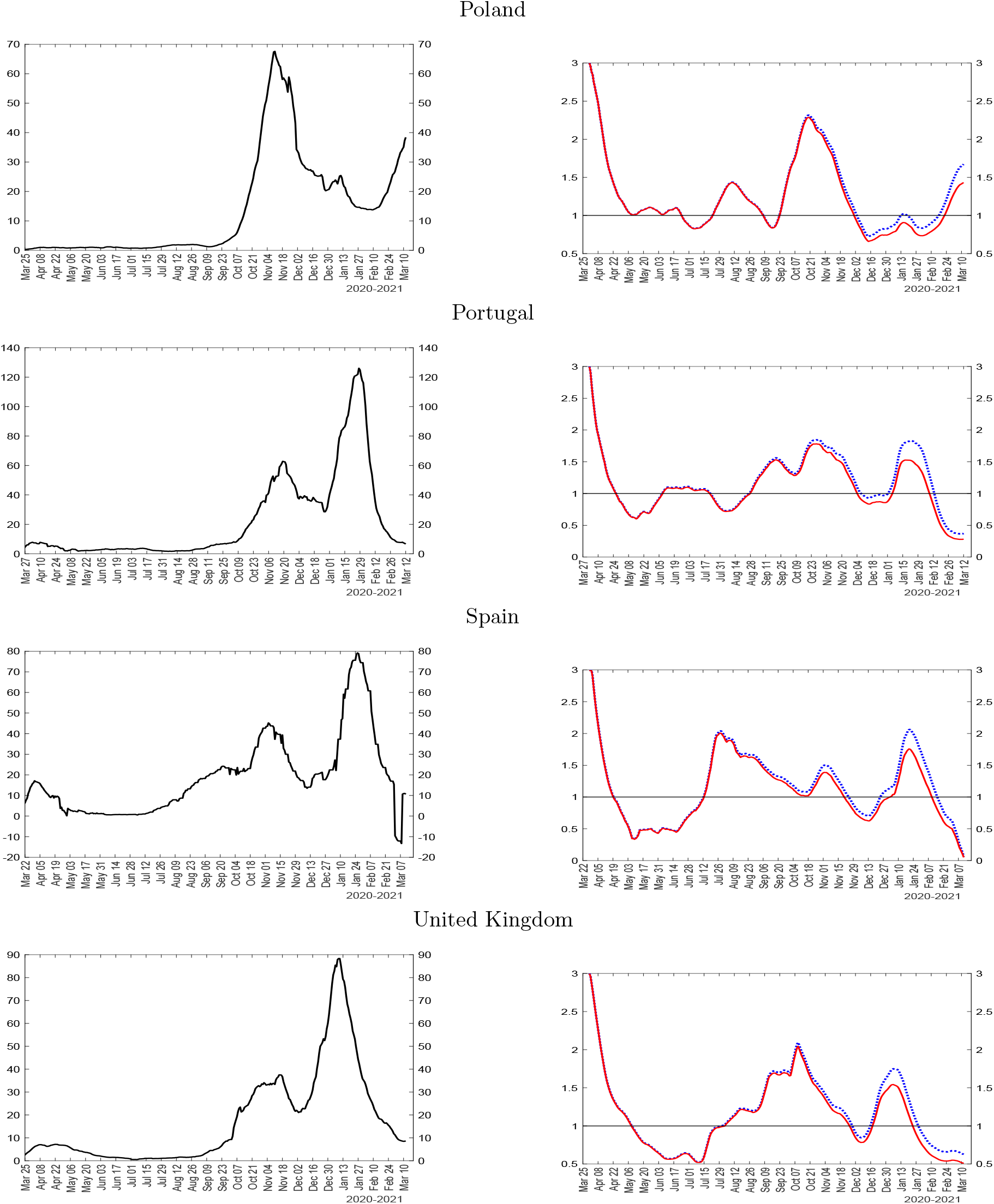
New cases (left) and *ℛ* numbers (right) for sample of European countries Notes: The figure plots a seven-day moving average of the number of reported new cases per 100k population (left charts), the *ℛ* number, 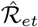 (right charts, solid red line), and the effective transmission rate, 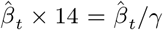 (right charts, dotted blue line). See notes to Figure 1.

Each panel reports two sets of charts. The charts on the left-hand-side of the figures report the seven-day moving average of the number of reported new infected cases per 100,000 population. The charts on the right-hand-side report two lines. The solid (red) line is the estimated R-number, 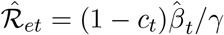. The dotted (blue) line is the effective transmission rate, 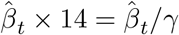. This is the variable that we model in Section 5. Recalling that the effective transmission rate, 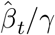, coincides with 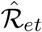 only when *c*_*t*_ *≈* 0, but as the epidemic spreads more widely we have *c*_*t*_ *>* 0, herd immunity can eventually start to play a non-negligible role and manifests itself in later stages of the epidemic with an increasing gap between 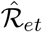 and 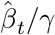, depending on the magnitude of *c*_*t*_. Also, we expect 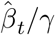 to be in the range 0 to 3 (similarly to 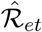), and 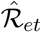 to be smaller or equal to the effective transmission rate as the epidemic progresses. Thus the gap between the red and the blue lines is a function of *s*_*t*_ = 1 − *c*_*t*_, the share of susceptible (not yet infected) population.

We start by estimating the effective transmission rate, 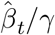, and hence the *ℛ* numbers, when the seven-day moving average of new cases exceeds a threshold of 50 cases to ensure a reasonably precise estimate of *β*_*t*_*/γ*. Note that at the early stages of the spread of the infection, when both *c*_*t*_ and *i*_*t*_ are close to zero, estimation of *β*_*t*_*/γ* becomes problematic as can be seen directly from (6). In effect it involves computing the ratio of two very small numbers, each subject to sampling errors. Note also that, since some countries (in particular China) were able to virtually eradicate the virus in some sub-periods, there will be gaps in our charts reporting the R numbers. In addition, we start to report estimated *ℛ* numbers at the beginning of the sample from the day in which 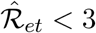 for the first time. This is to avoid showing widely varying estimated values in the initial days of the epidemic driven by unusually large growth rates of new confirmed cases, which could reflect delays in reporting the number of infected cases.

### 4.1 China and the rest of the world

#### 4.1.1 China

China experienced a large first wave followed by a few small and localized outbreaks (Figure 1). Two points are worth highlighting. First the *ℛ* number comes down very fast, in less than a month during the first wave. This is consistent with disaggregate evidence in Fang, Wang, and Yang (2020) and also clinical evidence. Second, the effective reproduction number always coincides with the effective transmission rate in the case of China, given the fact that only a very small fraction of population has been infected. The number of infected cases in China is 90, 000 *× MF* out of a population of 1.4 billion. This is a very small share even if we set *MF* to 20, which is at the upper end of the estimates reported for *MF* across many countries and reviewed in the Introduction. This confirms herd immunity had no role in the reduction of the effective reproduction number in the case of China.^12^ When the epidemic resurfaces, the estimated effective transmission rate increases sharply, but the extremely small number of cases permitted due to aggressive containment strategies prevented any new large-scale spread of the virus. Note that under mandatory social distancing the population never reaches herd immunity. So infections recur if containment is relaxed and the virus has not yet been fully eradicated. Figure 1 shows that China was successful not only in containing the epidemic at the start of its outbreak, but has thus far also been able to eradicate it quickly whenever it has re-surfaced through international travel. As we shall see, most other countries have not been able to accomplish this.

#### 4.1.2 The Rest of the World excluding China

The bottom panel of Figure 1 reports results for the rest of the world excluding China. As we noted earlier, these estimates are based on aggregate cases, as opposed to averages of country specific estimates. In the rest of the world, the COVID-19 epidemic started later than in China and the *ℛ* number comes down more slowly compared to China, never really falling below one until the end of 2020. The *ℛ* number increased from May to July 2020, and then again starting at the end of August 2020. As a result, the pandemic’s incidence was many, many times higher than in China in terms of cases. Indeed, our estimation results show that even an *ℛ* number slightly above one can be devastating once the epidemic has spread widely. Overall, the rest of the world as a whole never managed to eradicate the epidemic to an extent comparable to China. Not surprisingly, as restrictions ease during the summer of 2020, the epidemic resurfaces and worsens dramatically. Moreover, some of the decline in the *ℛ* number is due to herd immunity, which is extremely costly in terms of lives and, possibly, long term health consequences for the population.

### 4.2 Major World Regions

Comparing Northern and Southern Hemispheres reported in Figure 2, we see that climate has made a difference to both the initial spread, which was faster in the northern winter, and the shape of the epidemic curve, which was more persistent in the southern hemisphere. It does not, however, make a significant difference in terms of the epidemic peak; the number of daily new confirmed cases peaked about 10-12 per 100k population in the Northern Hemisphere, whereas the peak number of new cases (per 100k population) was about 10 the Southern Hemisphere in January of 2021. In the South, the *ℛ* number declined more slowly, but eventually dropped below one for several months in the middle of 2020. In both hemispheres, the estimates suggest that the COVID-19 transmission rate was falling in February 2021.

Figures 3-4 report the estimates at more regional levels of disaggregation.^13^ A stark difference emerges between the epidemic peaks reported in the left charts. North America reached a peak of 70 new cases per 100k population. Western Europe together with Eastern Europe and Central Asia experienced the second largest peaks at about 45-50 new cases per 100k population. Peaks in the daily new cases in Latin America and Caribbean region are also quite sizeable, but considerably smaller, staying below 20 new cases per 100k population. In contrast, the largest peak in the daily new cases is only about 7 in Middle East and North Africa, about 5 in South Asia, and even smaller peaks of less than 3 new cases per 100k population were achieved in East Asia and Pacific (excl. China) and Sub-Saharan Africa.

Large differences can be observed not only in terms of the magnitude of the peaks in new infections, but also in the trajectory of the epidemic more broadly. South Asia experienced a protracted single peak culminating in September 2021, which is reflected in the overall *ℛ* number not falling below one from the start of the epidemic until early in September 2020. By contrast, Sub-Saharan Africa experienced two definite peaks (July 2020 and January 2021). North America and Western Europe experienced three major waves. The first wave occurred in March/April in both regions. After some significant community spread of the virus, containment policies were enacted which helped to bring the *ℛ* number below one in a very short period of time. In North America, containment measures were relaxed quicker, and therefore the *ℛ* number did not stay below one for long, resulting in the second wave in the summer of 2020. By contrast, the *ℛ* number stayed below one for longer in Western Europe, until about mid-summer, when the virus began to spread exponentially again, resulting in the second (and largest) European wave in the Fall. After the new containment measures, *ℛ* number declined again, but it did not stay below one for long, resulting in the third wave of infections in January 2021 in both regions.

Experience from the remaining regions is more atypical than one might expect from the epidemic models. New cases in the Middle East and North Africa and, to some extent East Asia and Pacific (excl China), exhibit a broad upward trend throughout 2020 with a number of local peaks; new cases data for Latin America and Caribbean appear to be subject to much more noise compared with any other regions, and there is an unusual jump in the daily new cases in Eastern Europe and Central Asia, driven by the data for Turkey. *ℛ* numbers closely reflect the first derivative of the smoothed version of the new cases data in all regions; new cases subside when R falls below one and increase when R is above one.

The difference between the solid red lines (*ℛ* numbers) and the dotted blue lines (effective transmission rate) is virtually zero in the most successful regions in terms of the total number of cases, such as Sub-Saharan Africa and South Asia, suggesting that herd immunity played no role in these regions due to the relatively small number of overall infections. On the other hand, the gap between the two lines is largest in North America, followed by Western Europe, showing that herd immunity has started to contribute more meaningfully to mitigation of the epidemic in these regions starting in December 2020.

### 4.3 Selected Large Countries

Clearly the trajectory of the epidemics has been quite heterogeneous across regions. In addition, there are considerable differences across countries within each region, to which we now turn for selected large countries. We report estimates for the United States, Brazil, India and Russia in Figure 5, for South Africa, Australia, Iran and Turkey in Figure 6, and nine European countries in Figures 7-8—Belgium, France, Germany, Italy, Netherlands, Poland, Portugal, Spain, and UK. The selected countries include most of the G20 economies with the widest regional coverage globally.

In contrast to China’s and the rest of the world, the United States (reported in the top panel of Figure 5) stands out for the largest gap between the effective reproduction number and the effective transmission rate since the reopening of the economy in May 2020. The gap continues to widen throughout the subsequent period, peaking at the end of the sample in February 2021. Only a few other countries in the world, including the United Kingdom, Israel and some Latin American countries, display a comparable contribution of herd immunity to the decline in the *ℛ* number. The US case also stands out because of the three very distinct waves, with the second and the third re-emerging after a brief fall of the *ℛ* number below one. This led to a much higher number of infections per 100,000 people compared to the rest of the world.

Like the United States, Brazil’s estimates also show visible gaps between the *ℛ* number and the effective transmission rate starting in mid-2020. The case count in Brazil is more volatile compared to the United States and the remaining countries, possibly due to differences in the data quality other than under-reporting controlled for with the multiplication factor. Unlike the US case, Brazil brought down the *ℛ* number more gradually, falling below one for the first time only during the summer of 2020. This resulted in a protracted first wave that peaked in August. The *ℛ* number however did not remain below one for long, and in November a second large wave took off.

India also experienced a protracted first wave. Estimates of the *ℛ* number in India stayed above one until late September. Nevertheless, India did not experience a large number of cases per 100k population, compared with the remaining countries. As a result, herd immunity has not played a role in India. Russia, by contrast, experienced two large waves. Similarly to the western countries, Russia managed to bring the *ℛ* number down relatively fast, but not permanently, resulting in a larger second wave at the end of 2020.

A two-wave epidemic trajectory is also observed in the case of South Africa and Australia (in Figure 6), but with a different time profile. The first wave of the epidemic peaked in July 2020 in South Africa as authorities were unable to bring the *ℛ* number below one quickly enough. South Africa, as the richest country in the region, stands out with much higher infection rates compared to the rest of Africa. Australia, on the other hand, managed the virus very well. We can see two small peaks, one in March and the second in July-August 2020, each followed by a rapid decline in the *ℛ* number well below one, each time almost eradicating the virus without any discernible contribution from herd immunity.

For the two major neighboring countries, Iran and Turkey in the Middle East (the bottom two panels of Figure 6), the trajectories of the number of new cases differ markedly, with the outbreak of the virus starting much earlier in Iran due to the close trading relations with China. The initial spread in Iran began in late February 2020 and peaked in late March after the Iranian New Year (20^*th*^ March) and then declined slightly before starting to move up to its second peak in November 2020. By contrast, new cases in Turkey were detected in March and remained low for quite a few months before rising dramatically to a peak of 165 per 100,000 in December 2020. The associated *ℛ* numbers for Iran and Turkey also show very different trajectories, with Turkey’s *ℛ* number hitting the maximum value of 3 during the December 2020 peak.

The estimation results for selected European countries are reported in Figures 7-8. We report the same sample of countries as the one used in the next section for panel estimation of the transmission rate determinants. The virus outbreak in continental Europe begins with Italy in early 2020, with the recorded number of infections accelerating rapidly from February 21, 2020 onward. A rapid rise in infections takes place about one week later in Spain, Germany and France, followed by Austria (not reported) at the end of February. As the rolling estimates show, the *ℛ* number fell below one in mid- to late-April in all these countries. As lock-downs were eased during the summer, however, the transmission rates started to rise again. By the end of the 2020, the R numbers were much more dispersed, with some countries doing better than others. However, all large European countries reported in Figures 7-8 show a second wave much larger than the first one. The United Kingdom, Spain, Portugal and Netherlands exhibit distinct third waves, with larger case counts compared with their second-waves.

In summary, only China and a few other countries have been successful in containing the COVID-19 epidemic well. Contrary to common perception, however, not all countries accomplished this with the same draconian mandatory social distancing as in China. So we now turn to explaining the effective transmission rates to better understand the heterogeneity that we described, focusing on selected European countries reported in Figures 7-8, all experiencing quite similar starting dates and the initial wave of the epidemic, but quite differing subsequent trajectories.

## 5 Modelling the effective transmission rates in Europe

We saw earlier that the spread of the epidemic in Europe followed very similar patterns during the first wave, but diverged significantly towards the end of 2020 both in terms of epidemic peaks, level of effective reproduction numbers, and the importance of herd immunity in slowing down the spread of the virus. It is well known that many factors can contribute to the realized evolution of the epidemics, and particularly the effective reproduction numbers, *ℛ*_*et*_. Noting that *ℛ*_*et*_ = (1 − *c*_*t*_) *β*_*t*_*/γ*, in this section we develop a panel data model of the time evolution of *β*_*t*_ across a number of European countries jointly with *c*_*t*_ in terms of three fundamental drivers: mandatory and voluntary social distancing, as well as policy support that minimizes lack of compliance with mandatory social distancing. For policy analysis, it is important that the herding component of *ℛ*_*et*_ be distinguished from the virus transmission component *β*_*t*_*/γ*, as it is the latter that can be causally altered by mandatory and voluntary changes in social contacts. Recall here that based on clinical evidence we set *γ* = 1*/*14.

Also as noted already, over our sample period, that covers the first ten months of the pandemic, the evolution of transmission rates across countries are primarily determined by changes in behaviour (average contact numbers and exposure intensities per contact), with mutation playing a secondary role, and vaccination only having a very small role in the case of a few countries towards the end of our sample. Therefore, in explaining the cross country variations in *β*_*t*_ in what follows we shall focus on policy interventions and behavioural factors. Consistent with the simple decision theoretic model presented below, as well as a large literature on behavioral epidemic modeling, we consider three main factors: mandatory social distancing, economic incentives to comply with them in the form of economic support, and the awareness and information about COVID-19 and its rate of spread that can affect voluntary social distancing.

Mandatory social distancing directly reduces the number contacts as well as the exposure intensity. A strong theoretical rationale for imposition of mandated social distancing is the presence of externalities, i.e. the fact that agents do not internalize in their cost-benefit analysis that their individual behavior contributes to the aggregate diffusion of the epidemic.^14^ However, mandated social distancing imposes economic costs and infringes on individual liberty leading to personal inconveniences (Hamermesh, 2020).

Economic support to workers, households and small businesses during the pandemic can shape incentives of individuals to comply with mandatory social distancing, as it weakens the economic need to interact in work activities. Consider an individual who has a non-teleworkable job and is fired or furloughed. While this leads to an immediate loss of income, if economic support is adequate, individuals can weather the pandemic without needing to seek paid employment in exposed occupations continuing to interact in production activities. Lack of compliance with social distancing has been documented empirically by Wright et al. (2020). Based on survey evidence, Papageorge et al. (2021) find that higher income is associated with larger changes in self-protective behavior, particularly for individuals who cannot telework. They conclude that, both in the United States and elsewhere, policies which assume universal compliance with self-protective measures or that otherwise do not account for socio-economic differences in the costs of doing so are unlikely to be effective or sustainable.

It is also well understood that risk induces precautionary behavior. Behavioral models of COVID-19 diffusion show that, as the probability of getting infected rises, individuals lower consumption and leisure activities to avoid infection (see Eichenbaum, Rebelo, and Trabandt (2020), Toxvaerd (2020), Atkeson (2021), and Gupta, Simon, and Wing (2020)). In particular, Battiston and Gamba (2020) provide cross section evidence that the *ℛ* number during a COVID-19 outbreak is lower the larger the size of the initial wave.

### 5.1 A simple decision-theoretic model of voluntary social distancing

To further clarify and motivate our empirical approach and to provide some theoretical rationale behind our modelling strategy, here we introduce a simple decision-theoretic model of social distancing. Consider an individual *j* from a fixed population of size *n* in the epidemic day *t*, and suppose the individual in question is faced with the voluntary decision of whether to isolate or not. Under self-isolation, an individual that does not telework incurs the loss of wages net of any COVID-19 economic support amounting to (1 − *τ*_*jt*_)*w*_*jt*_, plus the inconvenience cost, *a*_*jt*_, of being isolated, where *w*_*jt*_ is the wage and *τ*_*jt*_ is the percentage of income lost which is compensated by the government support. For those individuals who can work from home *τ*_*jt*_ is likely to be 1 or very close to it. But for many workers who are furloughed or become unemployed, *τ*_*jt*_ is likely to be close to zero, unless they are compensated by transfers from the government.

On the other hand, if the individual decides not to self-isolate then he/she receives the uncertain pay-off of (1−*d*_*jt*_)*w*_*jt*_−*d*_*jt*_*ϕ*_*jt*_, where *d*_*jt*_ is an indicator which takes the value of unity if the individual contracts the disease and zero otherwise. The parameter *ϕ*_*jt*_ represents the cost of contracting the disease and is expected to be quite high. We are ruling out the possibility of death as an outcome and also assume that if the individual does not isolate and get sick does not earn the wage.

In this setting the individual decides to self-isolate if the sure loss of self-isolating is less than the expected loss of not self-isolating, namely if

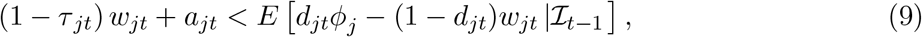

where *ℐ*_*t*−1_ is the publicly available information that includes *c*_*t*−1_, the total number of infections. We assume that the probability of anyone contracting the disease is uniform across the population and this is correctly perceived to be given by *π*_*t*−1_. Hence *E* (*d*_*jt*_ |*ℐ*_*t*−1_) = *π*_*t*−1_, and the condition for self-isolating in any day *t* can be written as

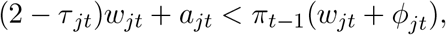

or as

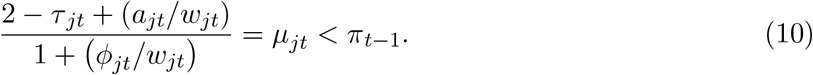

Since *π*_*t*−1_ ≤ 1, then for individual *j* to self-isolate we must have *µ*_*jt*_ *<* 1 (note that *µ*_*jt*_ ≥ 0, with *µ*_*jt*_ = 0 when *ϕ*_*jt*_ → ∞) or if

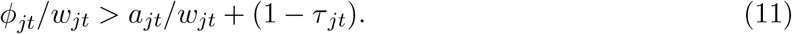

This condition clearly illustrates that an individual is more likely to self-isolate if the relative cost of contracting the disease, *ϕ*_*jt*_*/w*_*jt*_, is higher than the inconvenience cost of self-isolating plus the proportion of wages being lost due to self-isolation. Also, an individual is more likely to self-isolate voluntarily if the wage loss, measured by *τ*_*jt*_, is low thus providing an additional theoretical argument in favor of compensating some workers for the loss of their wages, not only to maintain aggregate demand but also to encourage a larger fraction of the population to comply with mandatory social distancing. The above formulation could also captures the differential incentive to self-isolate across different age groups and sectors of economic activity. Given that the epidemic affects the young and the old differently, with the old being more at risk as compared to the young, then *ϕ*_*old*_ *> ϕ*_*young*_, and the old are more likely to self-isolate. Similarly, low-wage earners are more likely to self-isolate as compared to high-wage earners with the same preferences (*ϕ*_*jt*_ and *a*_*jt*_), and facing the same transfer rates, *τ*_*jt*_. But the reverse outcome could occur if low-wage earner face a higher rate of transfer as compared to the high-wage earners. These and many other micro predictions of the theory are embedded in this specification of voluntary social distancing decision.

According to this simple model the fraction of population that are willing to socially isolate voluntarily is given by

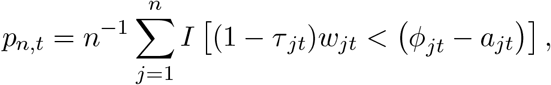

where *I*(*A*) is an indicator function that takes the value of 1 if *A* holds and zero otherwise. It is clear that the extent of voluntary social distancing, *p*_*n,t*_, is positively related to the size of the economic support, *τ*_*jt*_, and the perceived net cost of contracting the virus, *ϕ*_*jt*_ − *a*_*jt*_, which could rise sharply when epidemic surges and/or if better messaging by health authorities about the true costs of contracting the disease is provided. To capture voluntary as well as mandatory social distancing policies, in our statistical analyses we make use of data compiled by the Oxford COVID-19 Government Response Tracker (OxCGRT) project, which is a standard source of comparable indices measuring social distancing and other COVID-19 policies across countries.^15^ In particular, we use two aggregate indices: the ‘policy stringency index’ (capturing the containment and closure policies) and the ‘economic support index’ (as a proxy variable for support to comply with the containment policies). We model precautionary behavior leading to voluntary social distancing with a threshold effect explained in more detail below.

### 5.2 Statistical model

The econometric specification that we propose is based on the same framework used in Section 4 for the estimation of the evolution of the *ℛ* number. Namely, by taking logs on both sides of equation (6) above and recalling that *n* is large, we obtain

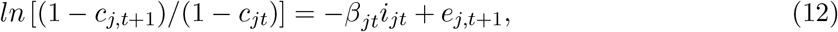

where the subscript *j* denotes individual countries, *j* = 1, 2,*…, N*, and *e*_*j,t*+1_ is an error term, assumed to be orthogonal to *i*_*jt*_. We complement the structural equation (12) with the following specification for the transmission rate process

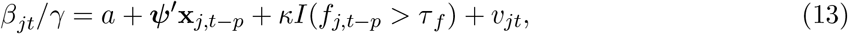

where **x**_*j,t*−*p*_ is a vector of regressors, lagged *p* periods, and the indicator variable, *I*(*f*_*j,t*−*p*_ *> τ*_*f*_), is an indicator variable which takes the value of unity if the threshold variable, *f*_*jt*_, also lagged *p* periods, goes above the threshold parameter *τ*_*f*_, which as a first-order approximation is assumed to be the same across countries. The choice of the threshold variable, *f*_*jt*_, is discussed below. In addition to estimating the panel regressions with a common constant term, *a*, we also check the robustness of our results by estimating the panel with fixed-effects where we replace *a* in (13) with country-specific intercept terms, *a*_*j*_, for *j* = 1, 2,*…, N*.

As a threshold variable, we use the 7-day moving average of the reported number of new cases (per 100,000 people), denoted by *f*_*jt*_, as this is the most commonly watched variable used in the media when reporting on the spread of COVID-19 worldwide. The idea for a threshold effect, defined in terms of the number of new cases, is to capture possible non-linearities and changes in people’s willingness to isolate consistent with surveys on the role of information diffusion under COVID-19 (Bursztyn et al., 2020).

Substituting (13) in (12), we obtain the following estimating equation:

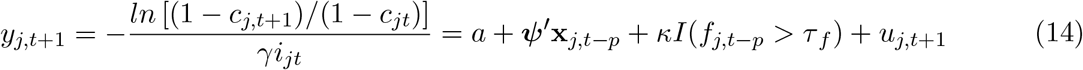

where *u*_*j,t*+1_ is the following composite error term:

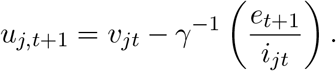

Depending on the assumptions regarding the error terms *u*_*j,t*+1_, we will report three types of standard errors for the estimates. First is the standard pooled (or fixed effects) standard errors, which assumes *u*_*j,t*+1_ is cross-sectionally as well as serially uncorrelated. Our second reported standard errors, labelled as “robust1”, allows for serial correlation and heteroskedasticity, while our third choice for standard errors, denoted as “robust2”, allows *u*_*j,t*+1_ to be correlated both over time as well as over countries (namely over both *j* and *t* dimensions). Further details are provided in the online Appendix.

The parameters of interest are *a* (or country-specific *a*_*j*_), ***ψ***, *κ*, and *τ* _*f*_. Our regressors are weakly exogenous and therefore the specification in equation (14) can be estimated by least squares, since the time series dimension of the panel is very large (*T* = 321 to 343) and the cross-section dimension is rather small (*N* = 9). This is in contrast to short panels (*T* small and *N* large), where strict exogeneity is required for consistency of least squares method. Additional concern with the specification could be omitted variables and the presence of other confounding factors. The proposed specification (14) is parsimonious and encompasses the main factors considered in the literature. As contacts and susceptibility are not separately identified in our model, we use aggregate indices of mandatory, support, and voluntary distancing, and we do not attempt to disentangle measures targeting the frequency of contacts or individual vulnerability.

We estimate the parameters of interest jointly using (14), as opposed to using a two-step procedure in which *β*_*jt*_ is estimated from (12) first and its determinants are estimated as a second stage regression from (13), using the estimated values of *β*_*jt*_ as the dependent variable from the first step. The advantage of estimating determinants of the transmission rate directly based on (14) is that the joint approach allows us to obtain more precise estimates and hence make more accurate inferences.

We focus on the nine European countries discussed in Section 4: Belgium, France, Germany, Italy, Netherlands, Poland, Portugal, Spain, and the United Kingdom– so *N* = 9 is relatively small. As we shall see, however, the time dimension of our panel is reasonably large (between 321 to 343 days), and we do not expect the relatively small *N* to be a problem for estimation and inference. The reason for focusing on these 9 countries is the fact that they experienced a similar start to the outbreak of the virus, but had differing outcomes subsequently. In this way we are able to exploit the cross country, as well as time series variations in the case data to identify the effects of mandatory and voluntary social distancing policies on the effective transmission rates, *β*_*jt*_*/γ*. Recall here that *β*_*jt*_*/γ* differs from the effective reproduction number, *ℛ*_*j,et*_, given by *ℛ*_*j,et*_ = (1−*c*_*jt*_) *β*_*jt*_*/γ*. As we noted earlier, *ℛ*_*j,et*_ can fall below unity not because of the effectiveness of the mitigating policies, but simply because an increasingly larger fraction of the population is getting infected, the so called herd-immunity effect. To avoid the confounding effect of herding on the outcome variable that we want to explain, we focus on modeling of *β*_*jt*_*/γ* and not *ℛ*_*j,et*_.

### 5.2 Empirical Results

Consider first the panel regressions without threshold effect:

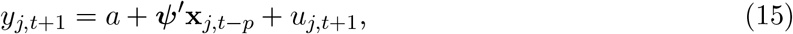

where *y*_*j,t*+1_ is defined in (14) and *j* = 1, 2,*…*, 9. The regressions are estimated with an unbalanced panel over the period February 23, 2020 to January 30, 2021, allowing for differences in the start dates of the outbreaks across the countries. The pooled estimates for this specification are reported in Table 1. We report results for *MF* = 3 and *MF* = 5 (the multiplication factor used to correct for under-reporting) and the lag orders *p* = 14 or *p* = 21 days. As can be seen, both the social distancing policy index and the index of policy support have the expected signs, and are both statistically highly significant. The results are robust to alternative MF corrections and lag lengths. Adding fixed effects slightly lowers the coefficient on the stringency index and increases the coefficient of the economic support indicator, but does not impact the overall conclusion. Using robust standard errors increases the estimated standard errors, as to be expected, but these increases are not sufficiently large to change the inference that we make. Both stringency and economic support indices remain highly significant.

**Table 1:** Pooled panel estimation results in the baseline specification without threshold effects

| Lag order:<br>Multiplication Factor: | Specifications with common constant |  |  |  | Specifications with fixed effects (FE) |  |  |  |
| --- | --- | --- | --- | --- | --- | --- | --- | --- |
| | $p = 14$ | | $p = 21$ | | $p = 14$ | | $p = 21$ | |
| | $MF = 3$ | $MF = 5$ | $MF = 3$ | $MF = 5$ | $MF = 3$ | $MF = 5$ | $MF = 3$ | $MF = 5$ |
| **Stringency Index** | **-3.37** | **-3.34** | **-3.40** | **-3.37** | **-3.12** | **-3.07** | **-3.23** | **-3.20** |
| standard s.e. (t-ratio) | 0.12 (-27.9) | 0.12 (-27.2) | 0.12 (-28.9) | 0.12 (-28.2) | 0.12 (-25.1) | 0.13 (-24.4) | 0.12 (-26.2) | 0.13 (-25.5) |
| robust1 s.e. (t-ratio) | 0.22 (-15.0) | 0.23 (-14.8) | 0.21 (-16.1) | 0.21 (-15.9) | 0.20 (-15.2) | 0.21 (-14.9) | 0.20 (-16.2) | 0.20 (-15.9) |
| robust2 s.e. (t-ratio) | 0.36 (-9.3) | 0.36 (-9.2) | 0.39 (-8.6) | 0.39 (-8.6) | 0.77 (-4.0) | 0.77 (-4.0) | 0.76 (-4.3) | 0.76 (-4.2) |
| **Economic Support** | **-0.97** | **-0.94** | **-0.64** | **-0.61** | **-1.61** | **-1.60** | **-1.05** | **-1.03** |
| standard s.e. (t-ratio) | 0.08 (-12.3) | 0.08 (-11.8) | 0.08 (-8.1) | 0.08 (-7.6) | 0.09 (-17.0) | 0.10 (-16.6) | 0.09 (-11.2) | 0.10 (-10.8) |
| robust1 s.e. (t-ratio) | 0.13 (-7.4) | 0.13 (-7.2) | 0.12 (-5.5) | 0.12 (-5.2) | 0.17 (-9.7) | 0.17 (-9.6) | 0.17 (-6.2) | 0.17 (-6.1) |
| robust2 s.e. (t-ratio) | 0.22 (-4.3) | 0.22 (-4.2) | 0.19 (-3.3) | 0.19 (-3.2) | 0.40 (-4.0) | 0.40 (-4.0) | 0.38 (-2.8) | 0.38 (-2.7) |
| **Constant terms** |  |  |  |  |  |  |  |  |
| **Common** (robust t-ratio) | 4.13 (10.6) | 4.13 (10.6) | 3.87 (10.0) | 3.87 (10.0) | - | - | - | - |
| FE (robust t-ratio): |  |  |  |  |  |  |  |  |
| **Belgium** | - | - | - | - | 4.41 (5.7) | 4.44 (5.7) | 4.00 (5.3) | 4.02 (5.4) |
| **France** | - | - | - | - | 4.46 (5.7) | 4.46 (5.7) | 4.11 (5.4) | 4.11 (5.4) |
| **Germany** | - | - | - | - | 3.92 (5.7) | 3.91 (5.7) | 3.70 (5.5) | 3.69 (5.5) |
| **Italy** | - | - | - | - | 4.35 (5.6) | 4.35 (5.6) | 4.05 (5.5) | 4.04 (5.5) |
| **Netherlands** | - | - | - | - | 4.39 (5.7) | 4.39 (5.7) | 3.96 (5.3) | 3.96 (5.3) |
| **Poland** | - | - | - | - | 3.91 (5.8) | 3.90 (5.8) | 3.63 (5.7) | 3.63 (5.7) |
| **Portugal** | - | - | - | - | 4.39 (5.7) | 4.40 (5.7) | 4.08 (5.5) | 4.10 (5.5) |
| **Spain** | - | - | - | - | 4.78 (5.6) | 4.80 (5.6) | 4.36 (5.2) | 4.38 (5.3) |
| **United Kingdom** | - | - | - | - | 4.93 (5.6) | 4.92 (5.6) | 4.42 (5.2) | 4.42 (5.2) |
| R-squared | 0.35 | 0.33 | 0.35 | 0.33 | 0.39 | 0.38 | 0.38 | 0.36 |

Notes: Total number of observations is 2991 with $N = 9$ countries, $T_{\min} = 321$ and $T_{\max} = 343$ days. The estimation sample is unbalanced at the beginning. Starting dates of individual country samples are: 6-March-2020 (Belgium), 1-March-2020 (France), 1-March-2020 (Germany), 23-February-2020 (Italy), 6-March-2020 (Netherlands), 16-March-2020 (Poland), 14-March-2020 (Portugal), 2-March-2020 (Spain), and 2-March-2020 (United Kingdom). The last period is 30-January-2021 for all countries. “Robust1” standard errors are robust to serial correlation only (Newey-West type correction), whereas “robust2” standard errors are robust to serial correlation as well as cross-sectional correlation. See online Appendix for a description of the estimation of standard errors. Oxford stringency and economic support indices are divided by 100 so that they take values between zero and one.

Another issue that could impact the inference that we draw is the heterogeneity in the slope coefficients across countries, namely the consequence of replacing ***ψ*** in (15) by ***ψ***_*j*_. Table 2 reports country-specific estimates of the coefficients of interest, as well as the associated Mean Group (MG) estimates together with their standard errors as in Pesaran and Smith (1995). The MG estimates are computed by taking simple averages of country-specific estimates, with their robust standard errors proposed by Pesaran and Smith (1995). An important advantage of the MG estimates is their robustness to slope heterogeneity, in which case MG is a consistent estimator of the average slopes. An additional advantage of the MG estimates is the robustness to weak cross-sectional correlations of the country-specific estimates (Chudik and Pesaran, 2019). The country-specific results continue to provide strong support for both explanatory factors. The reported MG estimates in Table 2 are all highly statistically significant and close to the fixed effect estimates in Table 1, which provide further evidence of the robustness of our main results to slope heterogeneity.

**Table 2:**
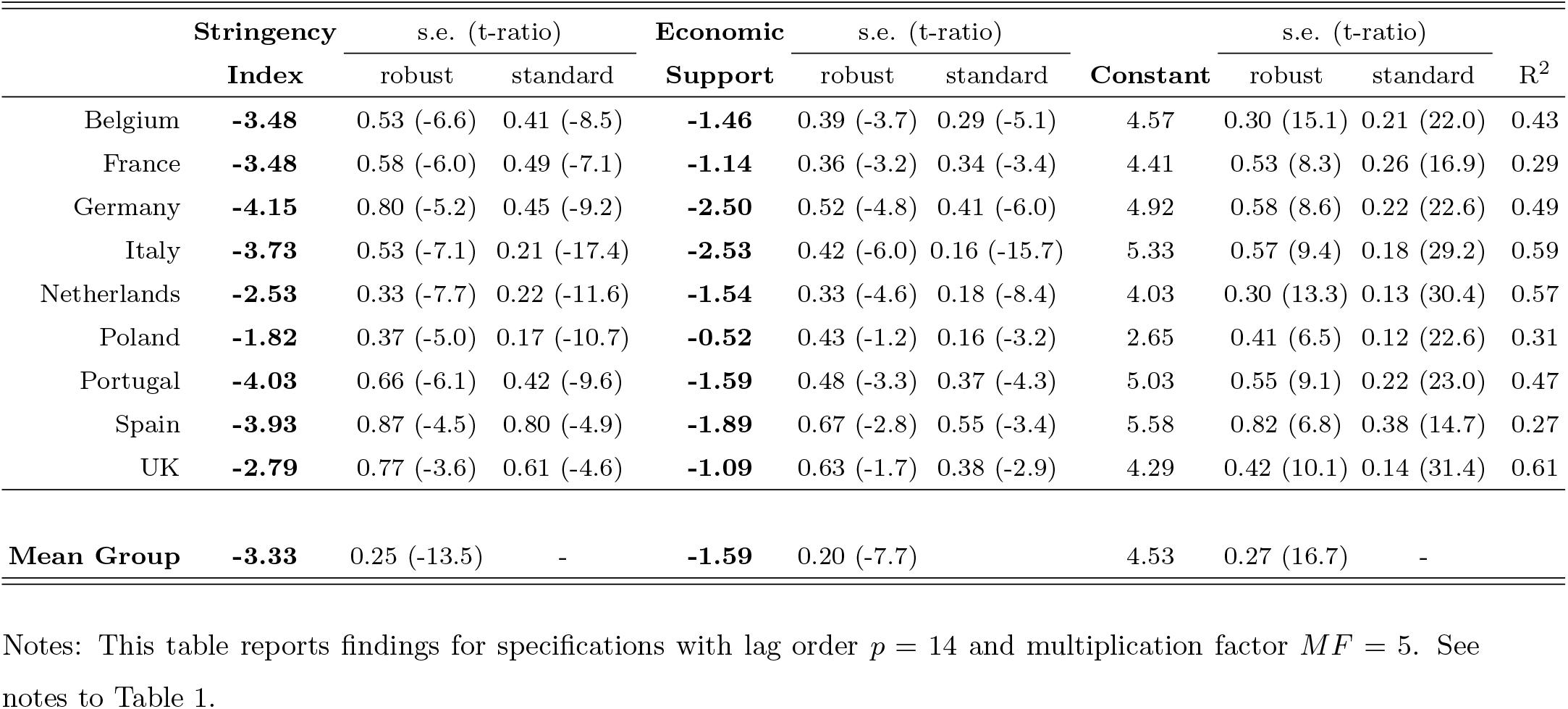
Individual country and mean group estimation results

|  | **Stringency** | s.e. (t-ratio) |  | **Economic** | s.e. (t-ratio) |  |  | s.e. (t-ratio) |  |  |
| --- | --- | --- | --- | --- | --- | --- | --- | --- | --- | --- |
|  | **Index** | robust | standard | **Support** | robust | standard | **Constant** | robust | standard | **R<sup>2</sup>** |
| Belgium | **-3.48** | 0.53 (-6.6) | 0.41 (-8.5) | **-1.46** | 0.39 (-3.7) | 0.29 (-5.1) | 4.57 | 0.30 (15.1) | 0.21 (22.0) | 0.43 |
| France | **-3.48** | 0.58 (-6.0) | 0.49 (-7.1) | **-1.14** | 0.36 (-3.2) | 0.34 (-3.4) | 4.41 | 0.53 (8.3) | 0.26 (16.9) | 0.29 |
| Germany | **-4.15** | 0.80 (-5.2) | 0.45 (-9.2) | **-2.50** | 0.52 (-4.8) | 0.41 (-6.0) | 4.92 | 0.58 (8.6) | 0.22 (22.6) | 0.49 |
| Italy | **-3.73** | 0.53 (-7.1) | 0.21 (-17.4) | **-2.53** | 0.42 (-6.0) | 0.16 (-15.7) | 5.33 | 0.57 (9.4) | 0.18 (29.2) | 0.59 |
| Netherlands | **-2.53** | 0.33 (-7.7) | 0.22 (-11.6) | **-1.54** | 0.33 (-4.6) | 0.18 (-8.4) | 4.03 | 0.30 (13.3) | 0.13 (30.4) | 0.57 |
| Poland | **-1.82** | 0.37 (-5.0) | 0.17 (-10.7) | **-0.52** | 0.43 (-1.2) | 0.16 (-3.2) | 2.65 | 0.41 (6.5) | 0.12 (22.6) | 0.31 |
| Portugal | **-4.03** | 0.66 (-6.1) | 0.42 (-9.6) | **-1.59** | 0.48 (-3.3) | 0.37 (-4.3) | 5.03 | 0.55 (9.1) | 0.22 (23.0) | 0.47 |
| Spain | **-3.93** | 0.87 (-4.5) | 0.80 (-4.9) | **-1.89** | 0.67 (-2.8) | 0.55 (-3.4) | 5.58 | 0.82 (6.8) | 0.38 (14.7) | 0.27 |
| UK | **-2.79** | 0.77 (-3.6) | 0.61 (-4.6) | **-1.09** | 0.63 (-1.7) | 0.38 (-2.9) | 4.29 | 0.42 (10.1) | 0.14 (31.4) | 0.61 |
| **Mean Group** | **-3.33** | 0.25 (-13.5) | - | **-1.59** | 0.20 (-7.7) |  | 4.53 | 0.27 (16.7) | - |  |

Notes: This table reports findings for specifications with lag order $p = 14$ and multiplication factor $MF = 5$ . See notes to Table 1.

Table 3 adds the threshold variable to (15). The estimated threshold effects are also highly statistically significant. Interestingly, the stringency and support indices now have smaller coefficients, pointing to a possible upward bias in estimating the effectiveness of mandatory social distancing policies if the voluntary mitigating effects, driven by better information dissemination or the fear factor, are ignored. These results suggest that the role of mandatory policies might be overestimated in studies that do not explicitly allow for voluntary changes in behavior.

**Table 3:** Pooled panel estimation results in the specification with threshold effects

| Lag order:<br>Multiplication Factor: | Specifications with common constant |  |  |  | Specifications with fixed effects (FE) |  |  |  |
| --- | --- | --- | --- | --- | --- | --- | --- | --- |
| | $p = 14$ | | $p = 21$ | | $p = 14$ | | $p = 21$ | |
| | $MF = 3$ | $MF = 5$ | $MF = 3$ | $MF = 5$ | $MF = 3$ | $MF = 5$ | $MF = 3$ | $MF = 5$ |
| **Stringency Index** | **-2.17** | **-2.14** | **-2.25** | **-2.23** | **-2.39** | **-2.35** | **-2.34** | **-2.31** |
| standard s.e. (t-ratio) | 0.13 (-17.2) | 0.13 (-16.6) | 0.12 (-18.7) | 0.12 (-18.1) | 0.12 (-19.1) | 0.13 (-18.4) | 0.12 (-18.9) | 0.13 (-18.3) |
| robust1 s.e. (t-ratio) | 0.17 (-12.8) | 0.17 (-12.6) | 0.15 (-15.4) | 0.15 (-15.1) | 0.17 (-14.2) | 0.17 (-13.9) | 0.14 (-16.2) | 0.15 (-15.9) |
| robust2 s.e. (t-ratio) | 0.22 (-10.0) | 0.22 (-9.6) | 0.18 (-12.3) | 0.19 (-11.8) | 0.35 (-6.8) | 0.36 (-6.6) | 0.41 (-5.7) | 0.41 (-5.6) |
| **Economic Support** | **-0.29** | **-0.27** | **-0.16** | **-0.13** | **-1.05** | **-1.04** | **-0.41** | **-0.40** |
| standard s.e. (t-ratio) | 0.08 (-3.6) | 0.08 (-3.3) | 0.08 (-2.1) | 0.08 (-1.7) | 0.10 (-11.0) | 0.10 (-10.7) | 0.09 (-4.4) | 0.10 (-4.2) |
| robust1 s.e. (t-ratio) | 0.11 (-2.7) | 0.11 (-2.5) | 0.09 (-1.8) | 0.09 (-1.5) | 0.14 (-7.4) | 0.14 (-7.4) | 0.12 (-3.5) | 0.12 (-3.4) |
| robust2 s.e. (t-ratio) | 0.11 (-2.7) | 0.11 (-2.5) | 0.09 (-1.8) | 0.09 (-1.5) | 0.20 (-5.2) | 0.20 (-5.2) | 0.17 (-2.4) | 0.18 (-2.3) |
| **Threshold variable** | **-2.29** | **-2.29** | **-2.35** | **-2.35** | **-2.24** | **-2.23** | **-2.20** | **-2.19** |
| standard s.e. (t-ratio) | 0.11 (-21.0) | 0.11 (-20.5) | 0.11 (-22.1) | 0.11 (-21.6) | 0.13 (-17.7) | 0.13 (-17.3) | 0.11 (-20.1) | 0.11 (-19.6) |
| robust1 s.e. (t-ratio) | 0.14 (-16.5) | 0.14 (-16.4) | 0.12 (-20.0) | 0.12 (-19.9) | 0.18 (-12.5) | 0.18 (-12.4) | 0.15 (-15.1) | 0.15 (-14.9) |
| robust2 s.e. (t-ratio) | 0.16 (-13.9) | 0.17 (-13.7) | 0.12 (-19.2) | 0.12 (-18.8) | 0.37 (-6.0) | 0.37 (-6.0) | 0.41 (-5.3) | 0.42 (-5.3) |
| threshold | 0.12 | 0.12 | 0.12 | 0.12 | 0.007 | 0.007 | 0.002 | 0.002 |
| Constant terms |  |  |  |  |  |  |  |  |
| common (robust t-ratio) | 5.11 (31.0) | 5.11 (30.7) | 5.11 (41.6) | 5.11 (41.0) | - | - | - | - |
| FE (robust t-ratio): |  |  |  |  |  |  |  |  |
| Belgium | - | - | - | - | 5.73 (16.6) | 5.74 (16.7) | 5.10 (13.7) | 5.12 (13.7) |
| France | - | - | - | - | 5.79 (17.1) | 5.79 (17.1) | 5.24 (14.0) | 5.24 (13.9) |
| Germany | - | - | - | - | 5.41 (17.9) | 5.38 (17.7) | 5.02 (15.0) | 5.00 (14.9) |
| Italy | - | - | - | - | 5.70 (17.0) | 5.69 (16.9) | 5.20 (14.3) | 5.18 (14.2) |
| Netherlands | - | - | - | - | 5.72 (17.3) | 5.71 (17.3) | 5.09 (13.9) | 5.09 (13.9) |
| Poland | - | - | - | - | 5.40 (18.5) | 5.38 (18.3) | 4.95 (15.1) | 4.94 (14.9) |
| Portugal | - | - | - | - | 5.78 (17.2) | 5.79 (17.2) | 5.29 (14.3) | 5.30 (14.2) |
| Spain | - | - | - | - | 6.04 (16.4) | 6.05 (16.4) | 5.41 (13.4) | 5.42 (13.3) |
| United Kingdom | - | - | - | - | 6.10 (16.4) | 6.09 (16.3) | 5.42 (13.1) | 5.41 (13.0) |
| R-squared | 0.43 | 0.41 | 0.44 | 0.42 | 0.45 | 0.43 | 0.45 | 0.44 |

Notes: Number of observations is 2991 with $N = 9$ countries, $T_{\min} = 321$ and $T_{\max} = 343$ days. “Robust1” standard errors are robust to serial correlation only (Newey-West type correction), whereas “robust2” standard errors are robust to serial correlation as well as cross-sectional correlation. See notes to Table 1.

The estimated coefficients in Table 3 of our preferred specification that includes the threshold effect predicts an *ℛ* number of around 5.11 (0.16) without any changes in behavior, be it voluntary and/or mandatory, with its standard error given in brackets. This is the reported estimated value of the common intercept, *α*, in Table 3, which is higher than the basic reproduction number of 3 − 4 assumed in the literature. In our earlier specifications that ignore the threshold effect in Table 1, the estimated reproduction number lies in a range between 3.87 to 4.13 (see the common intercept in Table 1). The contribution of the threshold effect to reducing the *ℛ* number is quite large, estimated in a tight range between −2.19 and −2.35, bringing the *ℛ* number down from 5.11 to just below 3.^16^

The contribution from the economic support index can also be interpreted as *voluntary* to the extent to which it captures the working of *incentives* to comply with mandatory restrictions as we discussed earlier. Our estimates of the contribution of the economic support to the change in the *ℛ* number range between −0.13 and −1.05, suggesting a high degree of heterogeneity across the specifications in Table 3, in contrast to the coefficients estimated for the other variables, which lie in a rather narrow range.^17^

Together with the contribution from the threshold effect, the upper bound on the impact of voluntary changes in behavior, therefore, is estimated to be reduced *ℛ* numbers ranging from 1.7 to 2.8, which is still well above unity. Recall that we need to bring the *ℛ* number below one before we are confident in falling case numbers. Hence, our estimates suggest that voluntary behavior alone is not sufficient to bring the *ℛ* number below one without substantial contributions from herd immunity.

In contrast, the estimates of the coefficients of the stringency index lie in the narrow range of −2.14 to −2.39 (see Table 3). We conclude from this exercise that mandatory social distancing *in conjunction* with the effects from the voluntary social distancing can bring the R below one without any additional help from the herding component. The estimates of the stringency index coefficients in Tables 1 and 3 also show that omitting to control for voluntary behavior can lead to overestimation of the impact of mandatory social distancing.

Finally, given the highly infectious nature of COVID-19, with its basic reproduction number estimated to be between 3 to 4, mandatory policies are likely to be effective if combined with mass vaccination programs, and the present research should be viewed as an interim report on a complex continuously evolving process.

## 6 Conclusions

This paper makes two related contributions. It first estimates effective reproduction numbers for all jurisdictions for which JHU reports COVID-19 statistics based on a moment condition that can be derived from an agent-based stochastic network epidemic model. It then explains their evolution distinguishing between herding, voluntary, mandatory social distancing and incentive to comply in a group of European countries with similar experiences at the outset of the pandemic but different outcomes subsequently.

From a methodological perspective, the econometric framework that we propose permits distinguishing, at any jurisdictional level, between changes in the effective reproduction number due to herd immunity and changes due to variation in the average contact or the susceptibility to infection, which are the structural determinant of the epidemic diffusion. At the empirical level, using only JHU daily COVID-19 case statistics, the paper provides estimates of transmission rates, allowing for the under-reporting of infected cases in available COVID case statistics.

Importantly, it is shown that while targeted mandated policies can be very useful in flattening the epidemic curve, especially at the onset of the epidemic, they are not necessarily sufficient. In some cases, countries with seemingly different social distancing policies achieved quite similar outcomes in terms of number of infections and reproduction numbers. There are also a small number of countries, for example Taiwan, Tanzania and Vietnam, that did not implement China’s draconian measures, yet have managed so far to accomplish similar low levels of infections and transmission rates.

Consistent with this result, when we turn to explain effective transmission rates in Europe we find that all considered factors played a significant role: the stringency of the mandatory policies implemented, the economic support and the threshold which captures the voluntary component of social distancing. However, our estimates suggest that voluntary social distancing alone is not sufficient to bring the *ℛ* number below one and to keep it there without substantial contributions from herd immunity. We also find that the role of the mandatory component, which is shown in the literature (including this paper) to be critical in containing and controlling the rapid spread of the virus, could be overestimated when voluntary mitigating drivers are neglected. Our main conclusions are robust to lag orders, error heteroskedasticity, error serial correlation, fixed effects and slope heterogeneity.

## Supporting information

Replication package

## Data Availability

All data needed, a full set of estimation results and the program codes to replicate the paper's results are available at the following website: sites.google.com/site/alexanderchudik/, pesaran.com.
sites.google.com/site/alessandrorebucciphd/)

https://www.google.com/url?q=https%3A%2F%2F www.dallasfed.org%2F-%2Fmedia%2Fdocuments%2Finstitute%2Fwpapers%2F2021%2F04072.zip&sa=D&sntz=1&usg=AFQjCNG0-K9v0xTf3EE_MC9anLuIgGL8uQ

## A Online Appendix

This online appendix is organized as follows. Section A.1 provides details regarding the estimation of standard errors in pooled regressions. Section A.2 provides regions definitions. Section A.3 provides comparisons of the estimated *ℛ* numbers for *MF* = 3 and 5, and Section A.4 provides comparisons of the estimated effective transmission rates for *MF* = 3 and 5.

### A.1 Conducting inference about the pooled panel results

Consider the panel data model (15), which, for convenience, can be equivalently written as

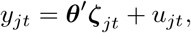

for *j* = 1, 2, *…, N*, where 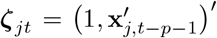. We allow for unbalanced panel by assuming *t* = 1, 2, *…, T*_*j*_. Let 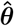 be the pooled estimator. We have

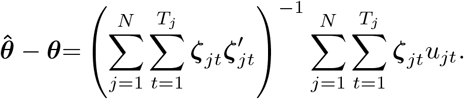

Variance of 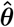 is given by

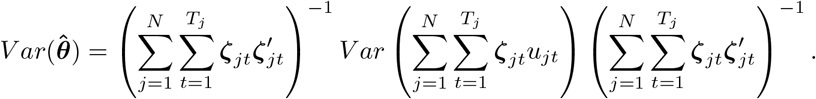

Assuming *E*(***ζ***_*jt*_*u*_*jt*_)= 0, we obtain

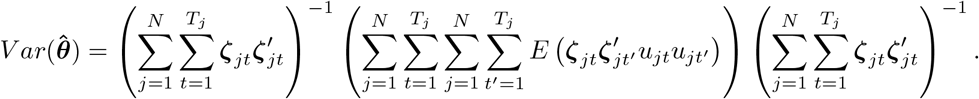

#### A.1.1 Inference robust to serial correlation of errors

Let

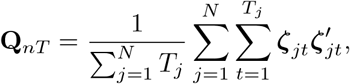

and

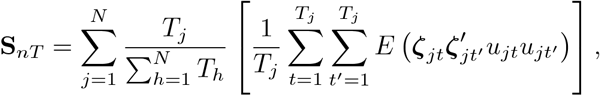

then

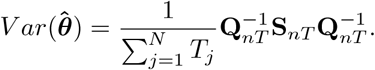

We estimate **S**_*nT*_ by the Newey-West method, extended to our panel setup:

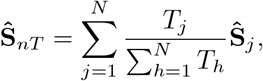

where

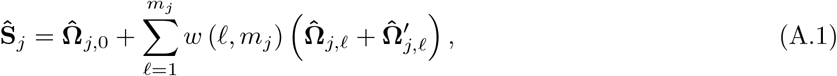

and

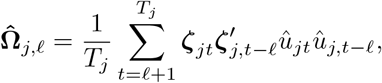

in which 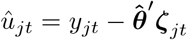. We set

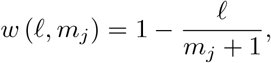

and *m*_*j*_ = *m*_*j,nT*_ is chosen to be a suitable increasing function of the sample size. We set *m*_*j,nT*_ to be the integer part of (*T*_*j*_)^1*/*3^.

#### A.1.2 Inference robust to serial and cross-sectional correlation of errors

Allowing for correlation of errors over time, as well as across units (countries) requires a different estimator of **S**_*nT*_. It is useful to re-write **S**_*nT*_ as

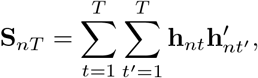

where

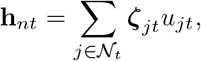

in which we use *𝒩*_*t*_ as the index set of cross-section units with available observations for a period *t*. **S**_*nT*_ is estimated as

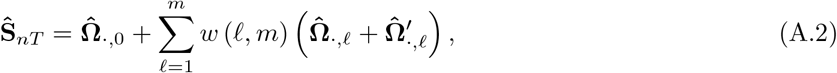

where

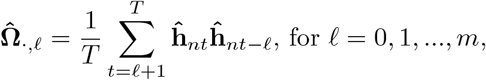

and

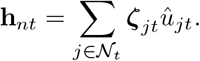

*m* = *m*_*nT*_ is chosen to be a suitable increasing function of the sample size. We set *m*_*nT*_ to be the integer part of *T* ^1*/*3^.

### A.2 Regions definitions

**Table A1:** Regions definitions

|  |
| --- |
| **East Asia and Pacific** |
| Australia, Brunei, Cambodia, China, South Korea, Fiji, Indonesia, Japan, Laos, Malaysia, Marshall Islands, Mongolia, Burma, New Zealand, Papua New Guinea, Philippines, Samoa, Singapore, Solomon Islands, Taiwan, Thailand, Timor-Leste, Vanuatu, Vietnam |
| **Eastern Europe and Central Asia** |
| Albania, Armenia, Azerbaijan, Belarus, Bosnia and Herzegovina, Bulgaria, Croatia, Georgia, Kazakhstan, Kyrgyzstan, Montenegro, Moldova, Romania, Russia, Serbia, Tajikistan, North Macedonia, Turkey, Ukraine, Uzbekistan |
| **Western Europe** |
| Andorra, Austria, Belgium, Cyprus, Czechia, Denmark, Estonia, Finland, France, Germany, Greece, Holy See, Hungary, Iceland, Ireland, Italy, Latvia, Liechtenstein, Lithuania, Luxembourg, Malta, Monaco, Netherlands, Norway, Poland, Portugal, San Marino, Slovakia, Slovenia, Spain, Sweden, Switzerland, United Kingdom |
| **North America** |
| United States, Canada |
| **Latin America and Caribbean** |
| Antigua and Barbuda, Argentina, Bahamas, Barbados, Belize, Bolivia, Brazil, Chile, Colombia, Costa Rica, Cuba, Dominica, Dominican Republic, Ecuador, El Salvador, Grenada, Guatemala, Guyana, Haiti, Honduras, Jamaica, Mexico, Nicaragua, Panama, Paraguay, Peru, Saint Kitts and Nevis, Saint Lucia, Saint Vincent and the Grenadines, Suriname, Trinidad and Tobago, Uruguay, Venezuela |
| **Middle East and North Africa** |
| Algeria, Bahrain, Egypt, Iran, Iraq, Israel, Jordan, Kuwait, Lebanon, Libya, Morocco, Oman, Qatar, Saudi Arabia, Syria, Tunisia, United Arab Emirates, Yemen |
| **South Asia** |
| Afghanistan, Bangladesh, Bhutan, India, Maldives, Nepal, Pakistan, Sri Lanka |
| **SubSaharan Africa** |
| Eastern and Southern Africa and West and Central Africa |
| **Eastern and Southern Africa** |
| Angola, Botswana, Burundi, Comoros, Djibouti, Eritrea, Ethiopia, Kenya, Lesotho, Madagascar, Malawi, Mozambique, Namibia, Rwanda, Seychelles, Somalia, South Africa, South Sudan, Sudan, Uganda, Tanzania, Zambia, Zimbabwe |
| **West and Central Africa** |
| Benin, Burkina Faso, Cabo Verde, Cameroon, Central African Republic, Chad, Congo (Brazzaville), Congo (Kinshasa), Cote d'Ivoire, Croatia, Equatorial Guinea, Gabon, Gambia, Ghana, Guinea, Guinea-Bissau, Liberia, Mali, Mauritania, Niger, Nigeria, Sao Tome and Principe, Senegal, Sierra Leone, Togo |

### A.3 Comparison of estimated *ℛ* numbers for selected countries and regions for two choices of multiplication factors, MF=5 and MF=3

**Figure A.1:**
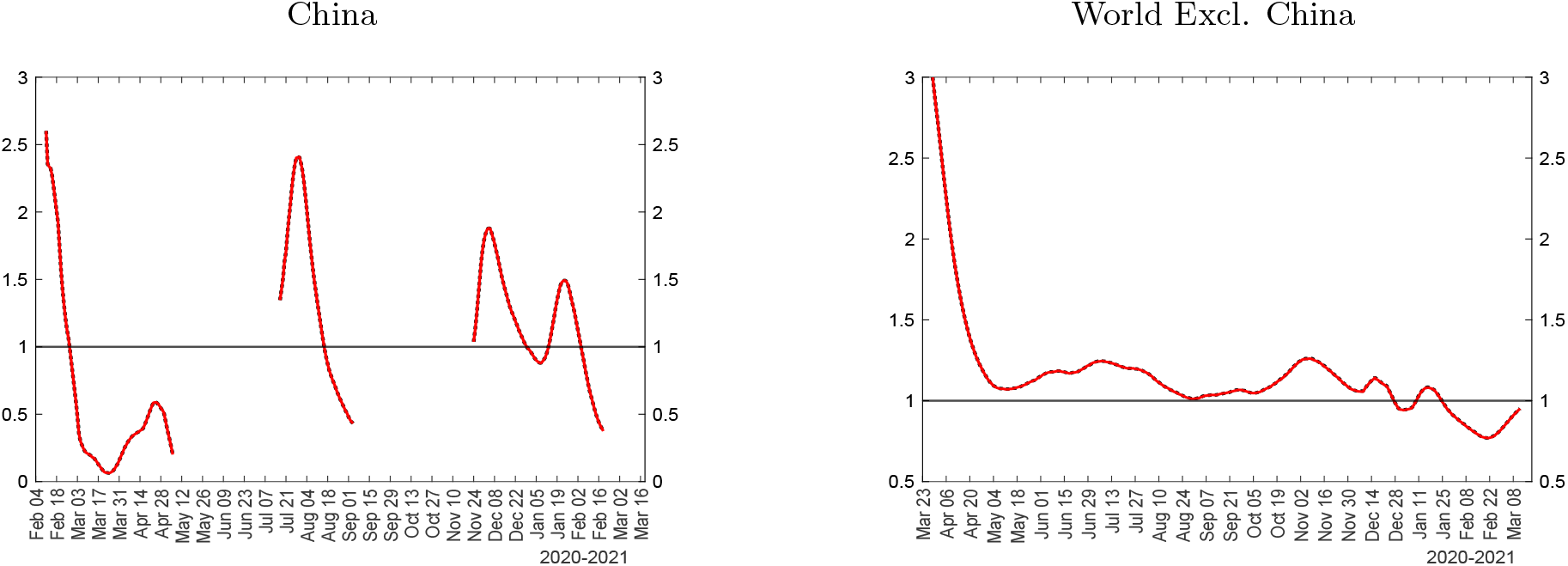
Comparisons of estimated *ℛ* numbers for China and the rest of the world for two choices of multiplication factors, MF=5 (solid red line) and MF=3 (dotted black line) Notes: The figure plots the *ℛ* number, 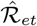, using MF=5 (solid red line) and MF=3 (dotted black line).

**Figure A.2:**
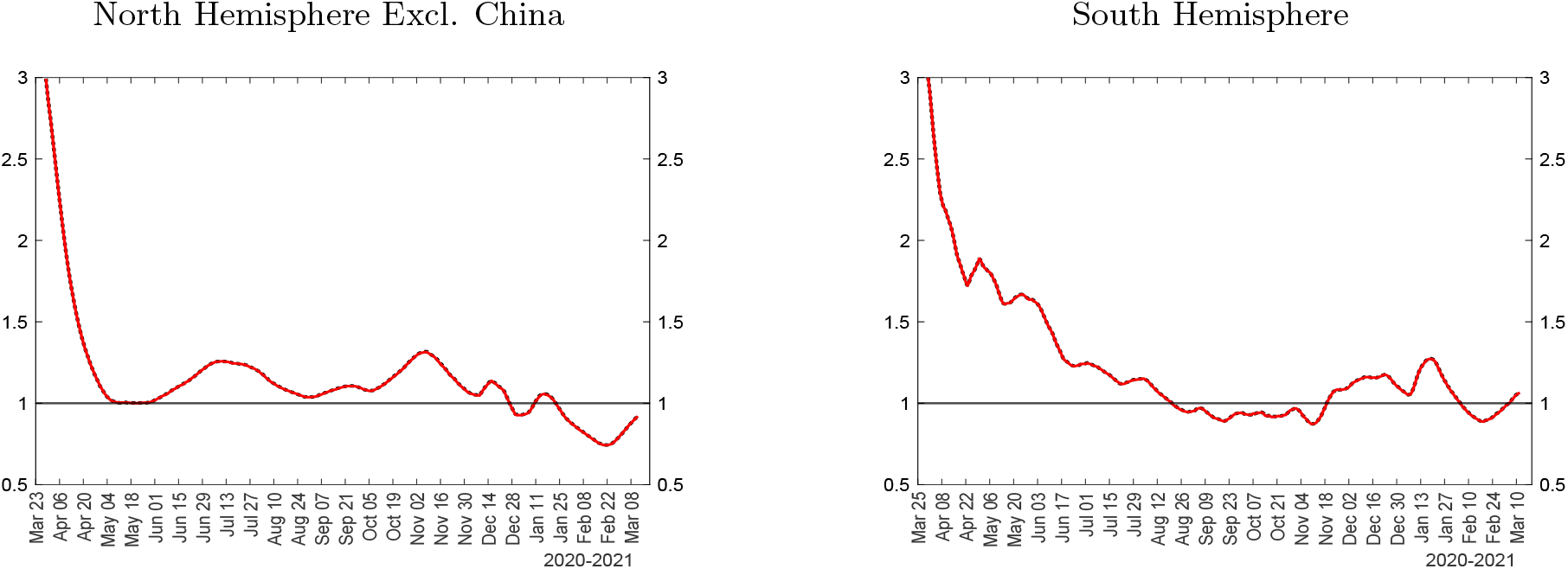
Comparisons of estimated *ℛ* numbers for North and South Hemispheres (excl. China) for two choices of multiplication factors, MF=5 (solid red line) and MF=3 (dotted black line) Notes: The figure plots the *ℛ* number, 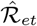, using MF=5 (solid red line) and MF=3 (dotted black line).

**Figure A.3:**
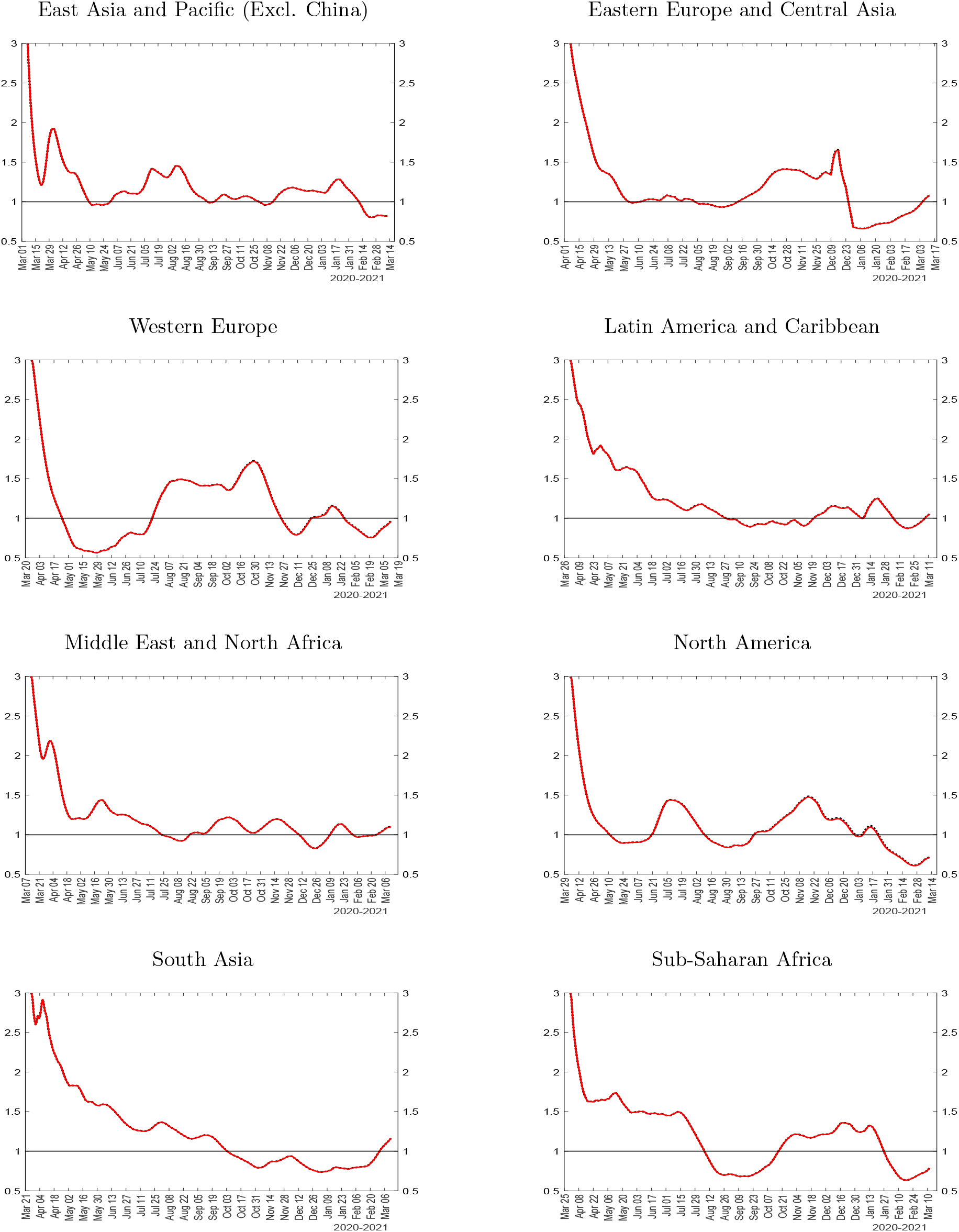
Comparisons of estimated *ℛ* numbers for main geographic regions (excl. China) for two choices of multiplication factors, MF=5 (solid red line) and MF=3 (dotted black line) Notes: The figure plots the *ℛ* number, 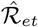, using MF=5 (solid red line) and MF=3 (dotted black line).

**Figure A.4:**
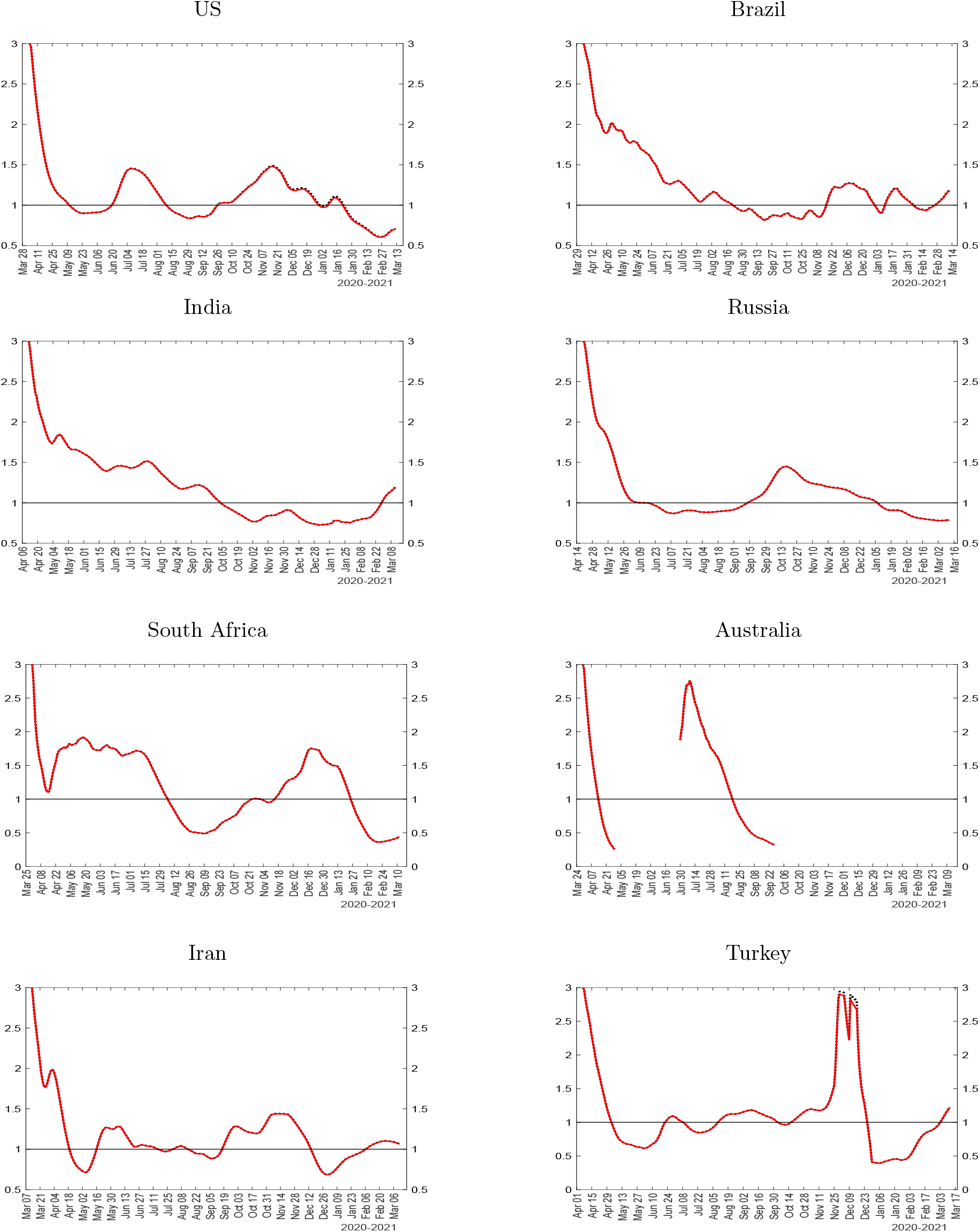
Comparisons of estimated *ℛ* numbers for selected countries for two choices of multi-plication factors, MF=5 (solid red line) and MF=3 (dotted black line) Notes: The figure plots the *ℛ* number, 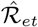, using MF=5 (solid red line) and MF=3 (dotted black line).

**Figure A.5:**
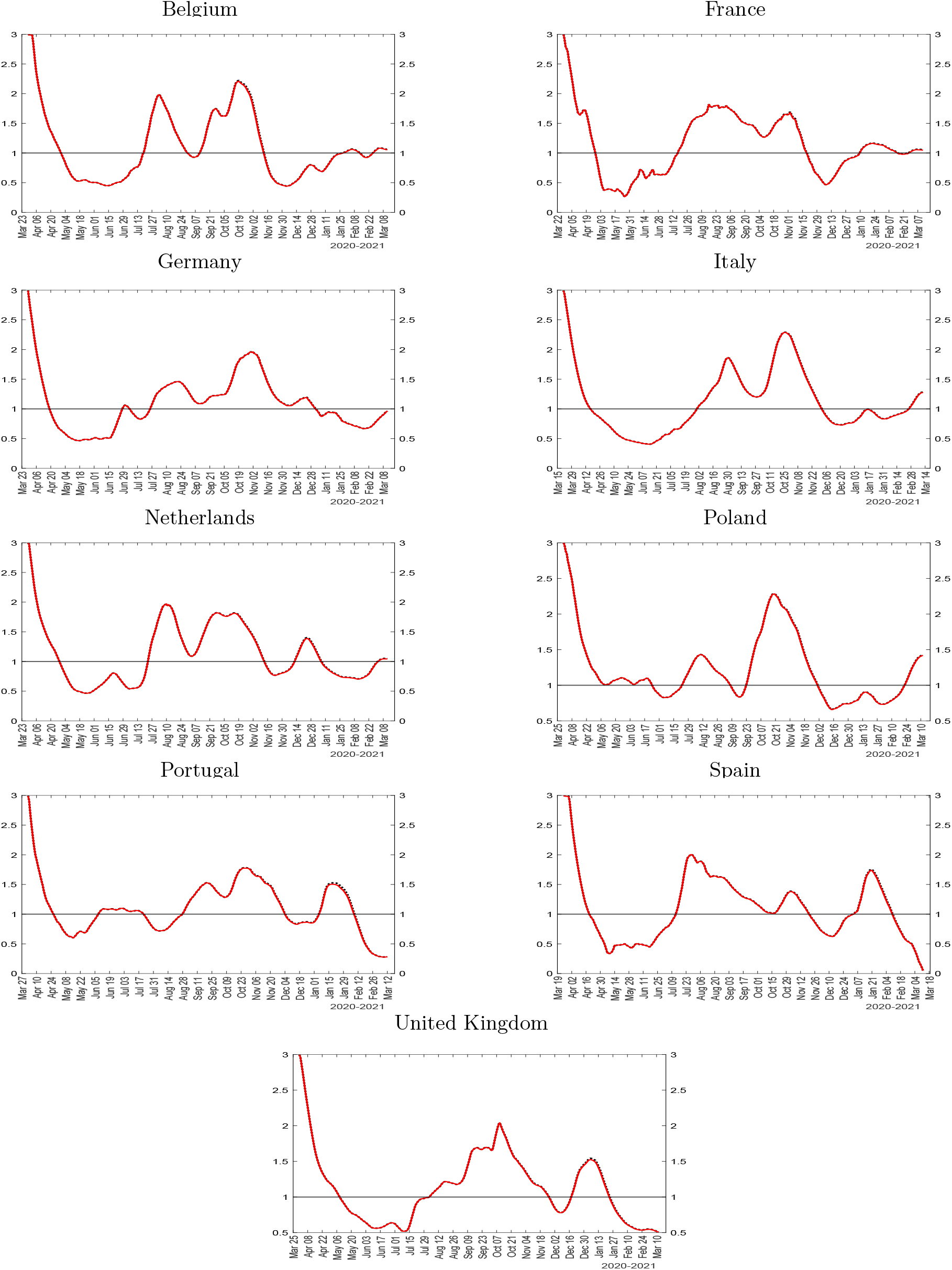
Comparisons of estimated *ℛ* numbers for sample of European countries for two choices of multiplication factors, MF=5 (solid red line) and MF=3 (dotted black line) Notes: The figure plots the *ℛ* number, 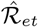, using MF=5 (solid red line) and MF=3 (dotted black line).

### A.4 Comparison of estimated transmission rates for selected countries for two choices of multiplication factors, MF=5 and MF=3

**Figure A.6:**
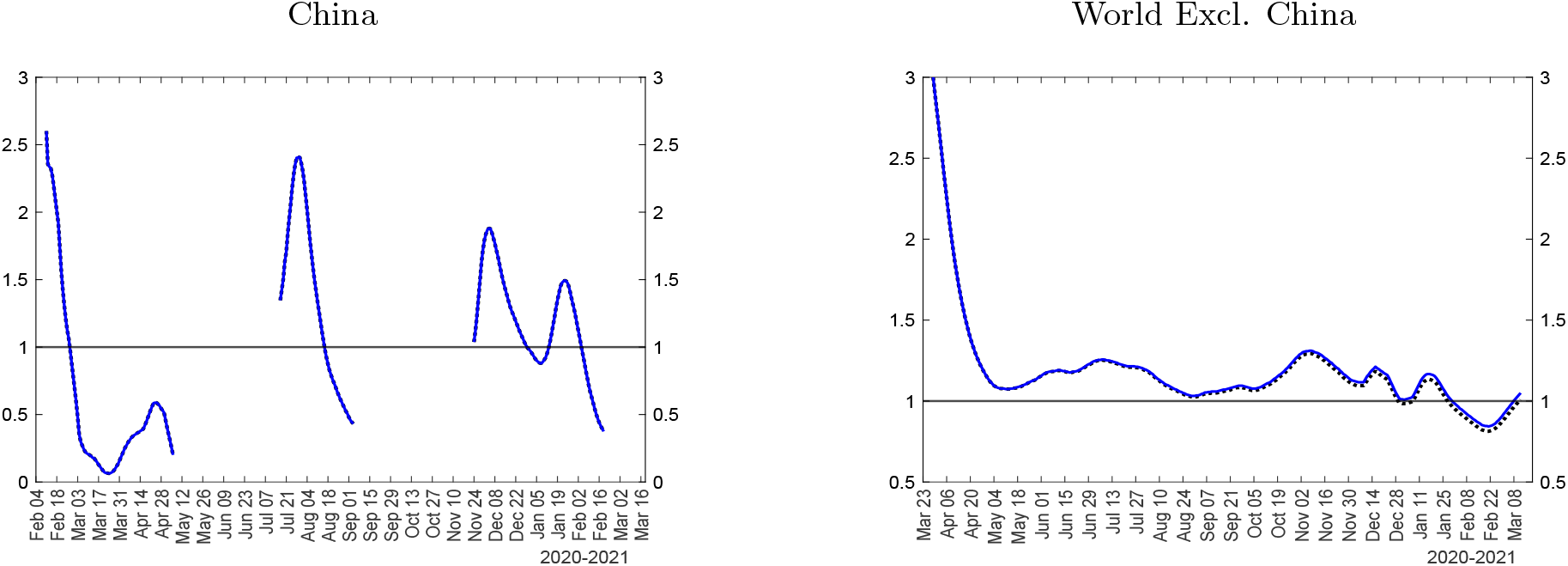
Comparison of estimated transmission rates for China and the rest of the world for two choices of multiplication factors, MF=5 (solid blue line) and MF=3 (dotted black line) Notes: The figure plots the effective transmission rate 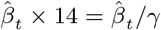 using multiplication factor MF=5 (solid blue line) and MF=3 (dotted black line).

**Figure A.7:**
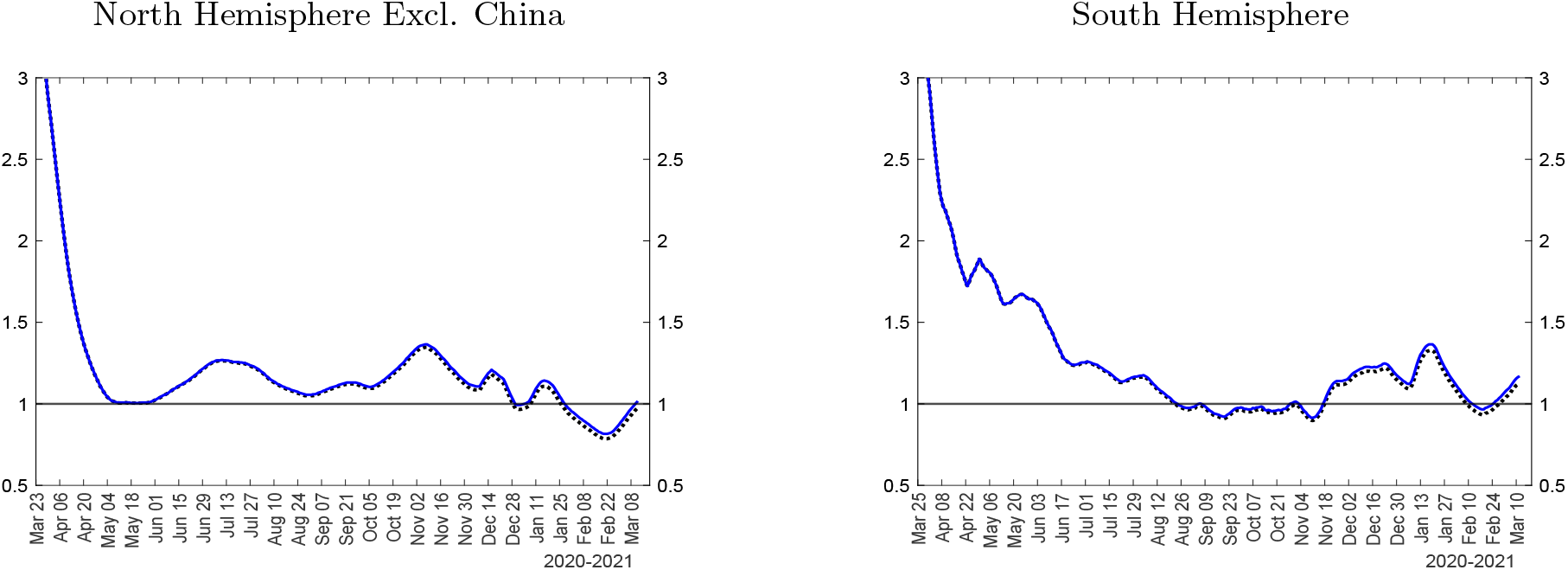
Comparison of estimated transmission rates for North and South Hemispheres (excl. China) for two choices of multiplication factors, MF=5 (solid blue line) and MF=3 (dotted black line) Notes: The figure plots the effective transmission rate 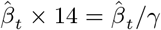 using multiplication factor MF=5 (solid blue line) and MF=3 (dotted black line).

**Figure A.8:**
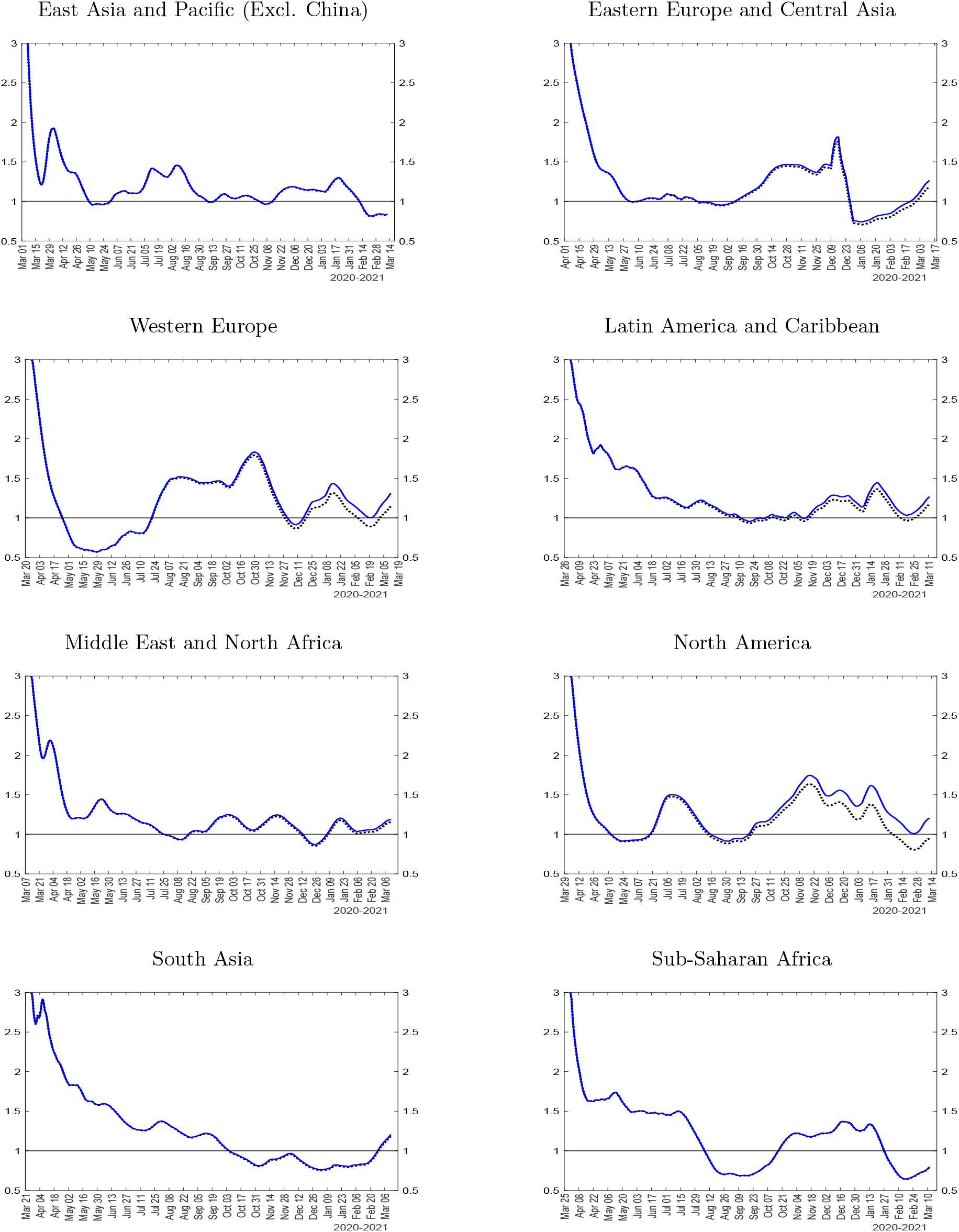
Comparison of estimated transmission rates for main geographic regions (excl. China) for two choices of multiplication factors, MF=5 (solid blue line) and MF=3 (dotted black line) Notes: The figure plots the effective transmission rate 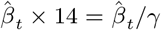 using multiplication factor MF=5 (solid blue line) and MF=3 (dotted black line).

**Figure A.9:**
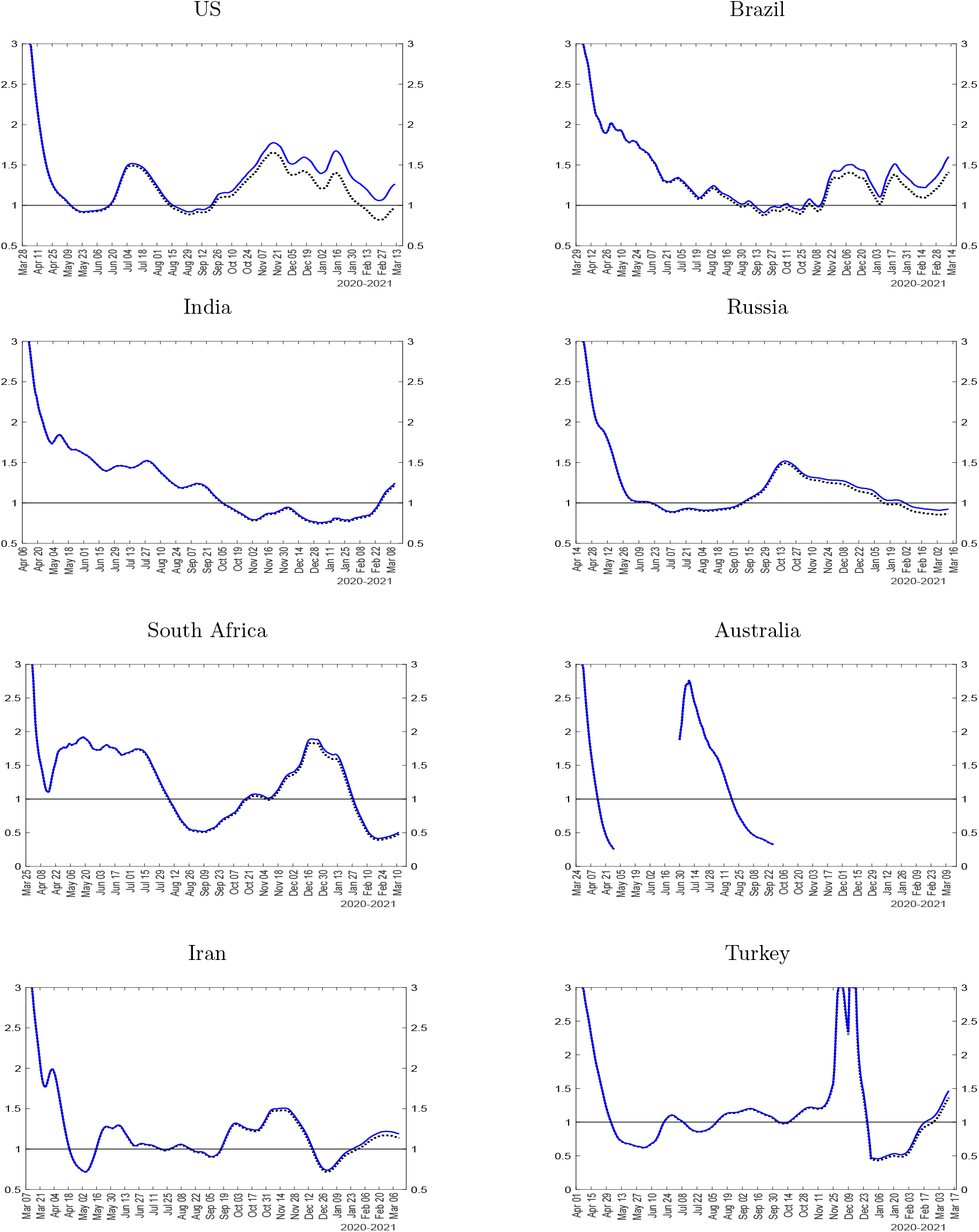
Comparison of estimated transmission rates for selected countries for two choices of multiplication factors, MF=5 (solid blue line) and MF=3 (dotted black line) Notes: The figure plots the effective transmission rate 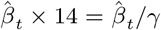 using multiplication factor MF=5 (solid blue line) and MF=3 (dotted black line).

**Figure A.10:**
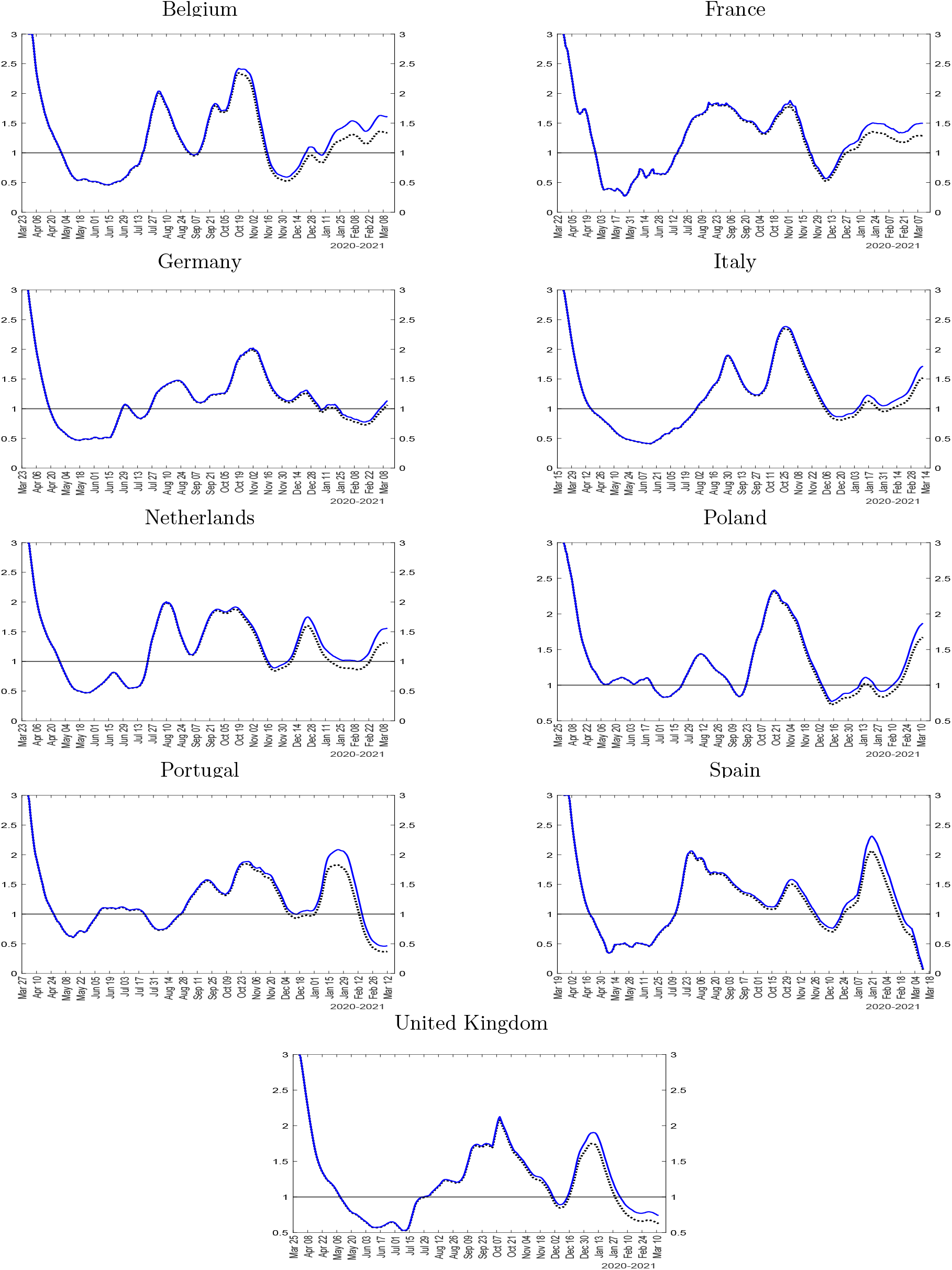
Comparison of estimated transmission rates for sample of European countries for two choices of multiplication factors, MF=5 (solid blue line) and MF=3 (dotted black line) Notes: The figure plots the effective transmission rate 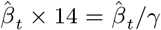 using multiplication factor MF=5 (solid blue line) and MF=3 (dotted black line).

## Footnotes

1 The full set of estimation results are available on the authors’ websites (sites.google.com/site/alexanderchudik/,pesaran.com, sites.google.com/site/alessandrorebucciphd/).

2 See Table 2 of Jagodnik et al. (2020). The seven countries considered are China, France, Italy, Spain, US,Germany and UK.

3 Available at https://www.bsg.ox.ac.uk/research/research-projects/coronavirus-government-response-tracker.

4 See Brodeur et al. (2020), Gupta, Simon, and Wing (2020), and Avery et al. (2020) for surveys of the early literature through the end 2020.

5 See for example Acemoglu et al. (2020), Akbarpour et al. (2020), Atkeson, Kopecky, and Zha (2020b),Cakmakli, Demiralp, Kalemli-Ozcan, Yesiltas, and Yildirim (2021), Cakmakli, Demiralp, Kalemli-Ozcan, Yesiltas, and Yildirim (2020), Matrajt and Leung (2020), Toda (2020), and Chudik, Pesaran, and Rebucci (2020) among many others.

6 For example, using data from Wuhan, Wang et al. (2020) report a pre-intervention reproductive rate of 3.86; Kucharski et al. (2020) estimate that, in China, the reproductive rate was 2.35 one week before travel restrictions were imposed on Jan 23, 2020. Ferguson et al. (2020) made the baseline assumption of *R*_0_ = 2.4 and also examined values of 2.0 and 2.6 based on fitting the early growth-rate of the epidemic in Wuhan by Li et al. (2020) and Riou and Althaus (2020).

7 An alternative, pursued for example by Fernández-Villaverde and Jones (2020), is to rely only on death data. While some countries might have good death statistics, using COVID-19 death data pose challenges similar to those raised by cases. The use of death data also has the added disadvantage of being a lagging indicator and could differ across countries due to factors such as age composition, obesity, and the quality of care system.

8 See, for example, the medical evidence documented in Ferguson et al. (2020) which implies a value for *γ* in the range 0.048 to 0.071. Our results are robust to assuming *γ* = 1*/*21.

9 The data from the Diamond Princess cruise ship reported by Moriarty et al. (2020) suggest about half of the COVID-19 cases are asymptomatic, and therefore *MF* =2 seems to be a good lower bound.

10 The COVID-19 data are sourced from the repository of the Center for Systems Science and Engineering (CSSE) at Johns Hopkins University–available at https://github.com/CSSEGISandData/COVID-19. The population data (for year 2019) are obtained from the World Bank database, available at https://data.worldbank.org/indicator/SP.POP.TOTL.

11 The full set of estimation results is available on the authors’ websites (sites.google.com/site/alexanderchudik/, pesaran.com, sites.google.com/site/alessandrorebucciphd/).

12 The effective reproduction number coincides with the effective transmission rate in most other Asian countries. Nonetheless, even in Asia, we observe a great deal of heterogeneity in terms of the shape of the epidemic curve. Japan and Indonesia fared better at the start of the pandemic, but did not avoid a large second wave. South Korea, in contrast, had two waves, one in March 2020 and a second toward the end of 2020, possibly reflecting its decision to avoid China-style mandatory social distancing, embracing a strategy revolving around testing and tracing with less restrictive limits on mobility and interactions (results not reported but available from the authors).

13 Table A1 in the online Appendix lists countries included in each region.

14 See, for example, Bethune and Korinek (2020), Eichenbaum, Rebelo, and Trabandt (2020), and Beck and Wagner (2020) in the international context. Eichenbaum, Rebelo, and Trabandt (2020), in particular, propose a behavioral SIR-macro model in which susceptible workers and consumers react to the epidemic risk by reducing their labor supply and consumption. Infected individuals take the aggregate infection process as given. As a result mandatory social distancing is optimal even though it is extremely costly in economic terms.

15 Data available at https://www.bsg.ox.ac.uk/research/research-projects/coronavirus-government-response-tracker.

16 Interestingly, the threshold effect kicks in at relatively very low levels of infections. In particular, the estimated threshold value is only 0.12 cases per 100,000 population.

17 Regarding the interpretation of the estimated coefficients of the stringency and economic support variables, we note here that we have divided the 0-100 Oxford stringency and economic support indices by 100 so that our scaled variables take values between 0 (no restrictions/economic support) and 1 (maximum restrictions/economic support as defined by Hale et al., 2020).

