## Supplementary material for "COVID-19 Time-varying Reproduction Numbers Worldwide: An Empirical Analysis of Mandatory and Voluntary Social Distancing": Replication package: add_aggregates.m

```
function A=add_aggregates(A)
cl=country_list(A);
% add Canada
toadd={'Canada: Alberta';'Canada: British Columbia';'Canada: Manitoba';'Canada: New Brunswick';'Canada: Newfoundland and Labrador';'Canada: Northwest Territories';'Canada: Nova Scotia';'Canada: Ontario';'Canada: Prince Edward Island';'Canada: Quebec';'Canada: Saskatchewan';'Canada: Yukon'};
A=addagg(A,cl,toadd,'Canada',55);

% Add China
toadd={'China: Anhui';'China: Beijing';'China: Chongqing';'China: Fujian';'China: Gansu';'China: Guangdong';'China: Guangxi';'China: Guizhou';'China: Hainan';'China: Hebei';'China: Heilongjiang';'China: Henan';'China: Hong Kong';'China: Hubei';'China: Hunan';'China: Inner Mongolia';'China: Jiangsu';'China: Jiangxi';'China: Jilin';'China: Liaoning';'China: Macau';'China: Ningxia';'China: Qinghai';'China: Shaanxi';'China: Shandong';'China: Shanghai';'China: Shanxi';'China: Sichuan';'China: Tianjin';'China: Tibet';'China: Xinjiang';'China: Yunnan';'China: Zhejiang'};
A=addagg(A,cl,toadd,'China',33);

%Add Australia
toadd={'Australia: Australian Capital Territory';'Australia: New South Wales';'Australia: Northern Territory';'Australia: Queensland';'Australia: South Australia';'Australia: Tasmania';'Australia: Victoria';'Australia: Western Australia'};
A=addagg(A,cl,toadd,'Australia',-32);


return
```
