## Supplementary material for "COVID-19 Time-varying Reproduction Numbers Worldwide: An Empirical Analysis of Mandatory and Voluntary Social Distancing": Replication package: add_aggregatesW.m

```
function A=add_aggregatesW(A, Ac)
cl=country_list(A);
[worldlist,worldlist_exChina] =world_list();
[SHlist,NHlist]=hemispheres(worldlist, Ac);
[~,NHlist_exChina]=hemispheres(worldlist_exChina, Ac);

% add World
A=addagg(A,cl,worldlist,'World',0);
A=addagg(A,cl,worldlist_exChina,'World excl. China',0);

% Add Southern Hemisphere
A=addagg(A,cl,SHlist,'Southern Hemisphere',0);

% Add Northern Hemisphere
A=addagg(A,cl,NHlist,'Northern Hemisphere',0);
A=addagg(A,cl,NHlist_exChina,'Northern Hemisphere excl. China',0);

% regional aggregates
    % East Asia and Pacific
[EA_P,EA_P_exChina] =east_asia_and_pacific();
A=addagg(A,cl,EA_P,'East Asia and Pacific',0);
A=addagg(A,cl,EA_P_exChina,'East Asia and Pacific excl. China',0);

    % Eastern Europe and Central Asia
[EE_CA] =Eastern_Europe_and_Central_Asia();
A=addagg(A,cl,EE_CA,'Eastern Europe and Central Asia',0);
    % Western Europe
[WE] =Western_Europe();
A=addagg(A,cl,WE,'Western Europe',0);
    % Latin America and Caribbean
[LA_C] =Latin_America_and_Caribbean();
A=addagg(A,cl,LA_C,'Latin America and Caribbean',0);
    % Middle East and North Africa
[list] =Middle_East_and_North_Africa();
A=addagg(A,cl,list,'Middle East and North Africa',0);

    % South Asia
[list] =South_Asia();
A=addagg(A,cl,list,'South Asia',0);
    % Eastern and Southern Africa
[list] =Eastern_and_Southern_Africa();
A=addagg(A,cl,list,'Eastern and Southern Africa',0);
    % West and Central Africa
[list] =West_and_Central_Africa();
A=addagg(A,cl,list,'West and Central Africa',0);

        % SubSaharan Africa
    [list] =SubSaharan_Africa();
    A=addagg(A,cl,list,'SubSaharan Africa',0);
        % North America
    [list] =NorthAmerica();
    A=addagg(A,cl,list,'North America',0);
return
```
