## Supplementary material for "COVID-19 Time-varying Reproduction Numbers Worldwide: An Empirical Analysis of Mandatory and Voluntary Social Distancing": Replication package: add_aggregatesWo.m

```
function Ano=add_aggregatesWo(Ao, Ac, Ap)
cl=Ao(2:end,1,1);
h=size(Ao,3);

[worldlist,worldlist_exChina] =world_list();
[SHlist,NHlist]=hemispheres(worldlist, Ac);
[~,NHlist_exChina]=hemispheres(worldlist_exChina, Ac);

for i=1:h
    A=Ao(:,:,i);   
    
    % add World
    A=addaggo(A,cl,worldlist,'World',0, Ap);
    A=addaggo(A,cl,worldlist_exChina,'World excl. China',0, Ap);

    % Add Southern Hemisphere
    A=addaggo(A,cl,SHlist,'Southern Hemisphere',0, Ap);

    % Add Northern Hemisphere
    A=addaggo(A,cl,NHlist,'Northern Hemisphere',0, Ap);
    A=addaggo(A,cl,NHlist_exChina,'Northern Hemisphere excl. China',0, Ap);

    % regional aggregates
        % East Asia and Pacific
    [EA_P,EA_P_exChina] =east_asia_and_pacific();
    A=addaggo(A,cl,EA_P,'East Asia and Pacific',0, Ap);
    A=addaggo(A,cl,EA_P_exChina,'East Asia and Pacific excl. China',0, Ap);

        % Eastern Europe and Central Asia
    [EE_CA] =Eastern_Europe_and_Central_Asia();
    A=addaggo(A,cl,EE_CA,'Eastern Europe and Central Asia',0, Ap);
        % Western Europe
    [WE] =Western_Europe();
    A=addaggo(A,cl,WE,'Western Europe',0, Ap);
        % Latin America and Caribbean
    [LA_C] =Latin_America_and_Caribbean();
    A=addaggo(A,cl,LA_C,'Latin America and Caribbean',0, Ap);
        % Middle East and North Africa
    [list] =Middle_East_and_North_Africa();
    A=addaggo(A,cl,list,'Middle East and North Africa',0, Ap);

        % South Asia
    [list] =South_Asia();
    A=addaggo(A,cl,list,'South Asia',0, Ap);
        % Eastern and Southern Africa
    [list] =Eastern_and_Southern_Africa();
    A=addaggo(A,cl,list,'Eastern and Southern Africa',0, Ap);
        % West and Central Africa
    [list] =West_and_Central_Africa();
    A=addaggo(A,cl,list,'West and Central Africa',0, Ap);
        % SubSaharan Africa
    [list] =SubSaharan_Africa();
    A=addaggo(A,cl,list,'SubSaharan Africa',0, Ap);
        % North America
    [list] =NorthAmerica();
    A=addaggo(A,cl,list,'North America',0, Ap);
    
    Ano(:,:,i)=A;
end

return
```
