## Supplementary material for "COVID-19 Time-varying Reproduction Numbers Worldwide: An Empirical Analysis of Mandatory and Voluntary Social Distancing": Replication package: addagg.m

```
function A=addagg(A,cl,toadd,agg_name, lat)


nt=size(toadd,1);
na=size(A,1);
nc=size(cl,1);
ind=false(na,1);

for j=1:nt
    for i=1:nc
        if strcmp(cl{i,1},toadd{j,1}) 
            ind(i+1,1)=true;
        end
    end
end

A{na+1,1}=missing();
A{na+1,2}=agg_name;
A{na+1,3}=lat;
for t=5:size(A,2)
    a=cell2mat(A(ind,t)); 
    A{na+1,t}=sum(a);
end

return
```
