## Supplementary material for "COVID-19 Time-varying Reproduction Numbers Worldwide: An Empirical Analysis of Mandatory and Voluntary Social Distancing": Replication package: addaggo.m

```
function A=addaggo(A,cl,toadd,agg_name, lat, Ap)


nt=size(toadd,1);
na=size(A,1);
nap=size(Ap,1);
nc=size(cl,1);
ind=false(na,1);
indp=false(nap,1);
indp2=false(nap,1);

clp=country_list(Ap);
ncp=size(clp,1);

S=false(nc,ncp); % transition matrix
for j=1:nt
    for i=1:nc
        if strcmp(cl{i,1},toadd{j,1}) 
            ind(i+1,1)=true;
%             % find position in Ap
%             for ii=1:ncp
%                 if strcmp(clp{ii,1},toadd{j,1}) 
%                     indp2(ii+1,1)=true;
%                     S(i+1,ii+1)=true;
%                 end
%             end
        end
    end
    for i=1:ncp
        if strcmp(clp{i,1},toadd{j,1}) 
            indp(i+1,1)=true;
        end
    end
end

% check if indp and ind are identical
eq=true;
for ii=1:size(indp,1);
   if not(indp(ii)==ind(ii) )
       eq=false;
   end
end
if not(eq)
    'error'
    dbstop
end


%A{na+1,1}=missing();
A{na+1,1}=agg_name;
A{na+1,3}=lat;

ind(end+1,1)=false;

% find total population of aggregate
pop=sum(cell2mat(Ap(indp,5)));
for t=5:size(A,2)
    % find share of countries withavailable data
     % available data in t
     aval=false(na+1,1);
     for jj=2:na
         if not(isempty(A{jj,t}))
             aval(jj,1)=true;
         end
     end
     indu=ind & aval;
     pa=sum(cell2mat(Ap(indu,5)));
     if pa>0.8*pop % if at least half population with data, then compute aggregate
         pw=cell2mat(Ap(indu,5));
         ad=cell2mat(A(indu,t));
         A{na+1,t}=pw'*ad/sum(pw);
     else
         A{na+1,t}=[];
     end
end

return
```
