## Supplementary material for "COVID-19 Time-varying Reproduction Numbers Worldwide: An Empirical Analysis of Mandatory and Voluntary Social Distancing": Replication package: betaestim.m

```
function betahat=betaestim(c0,c1,i1, lb, ub, steps)

ball=lb+[1:steps]'*ub/steps;
W=size(c0,1);
tau=ones(W,1);

objvals=zeros(steps,1);
for s=1:steps
    beta=ball(s,1);
    e=(tau-c0)./(tau-c1)-exp(-beta*i1);
    objvals(s,1)=e'*e;
end

[~,ind]=min(objvals);
betahat=ball(ind);
return
```
