## Supplementary material for "COVID-19 Time-varying Reproduction Numbers Worldwide: An Empirical Analysis of Mandatory and Voluntary Social Distancing": Replication package: compute_dc.m

```
function [dcm,dCm] =compute_dc(C,PT, W)

if nargin<3
    W=7;
end
[n,T]=size(C);
%daily cases
dC=NaN(size(C));
dC(:,2:end)=C(:,2:end)-C(:,1:end-1);
% per 100K
dcp=dC./(PT/100000);
%7DMA
    dcm=zeros(n,T);
    dCm=zeros(n,T);
    for t=W:T
        dcm(:,t)=dcp(:,t-W+1:t)*ones(W,1)/W;
        dCm(:,t)=dC(:,t-W+1:t)*ones(W,1)/W;
    end


return
```
