## Supplementary material for "COVID-19 Time-varying Reproduction Numbers Worldwide: An Empirical Analysis of Mandatory and Voluntary Social Distancing": Replication package: compute_trans_rates_hc.m

```
function [btw, re]=compute_trans_rates_hc(c, i, W,I,MAtransform, sv)

[n,T]=size(c);

if MAtransform % compute 7-day ma
    ima=zeros(n,T);
    cma=zeros(n,T);
    for t=7:T
        ima(:,t)=i(:,t-6:t)*ones(7,1)/7;
        cma(:,t)=c(:,t-6:t)*ones(7,1)/7;
    end
    i=ima;
    c=cma;
end

stbeta=3/14;
options = optimset('Display','off', 'TolX',1e-6);
btw=NaN(n,T);
lb=0;
ub=3/14;
steps=10^3;

for j=1:n
    for t=W+1:T
         iw=I(j,t-W+1:t)';        

         if computebeta(iw, sv) %&& started
            ci0=c(j,t-W+1:t)';
            ci1=c(j,t-W:t-1)';
            i1=i(j,t-W:t-1)';
            betaihat=betaestim(ci0,ci1,i1, lb, ub, steps);
          
            btw(j,t)=betaihat;
        end
    end
end
%% now compute effective R
gamma=1/14;
re=(btw/gamma).*(1-c);
return

function obj=objg(beta,c0,c1,i1)

W=size(c0,1);
tau=ones(W,1);
e=(tau-c0)./(tau-c1)-exp(-beta*i1);

obj=e'*e;

return
```
