## Supplementary material for "COVID-19 Time-varying Reproduction Numbers Worldwide: An Empirical Analysis of Mandatory and Voluntary Social Distancing": Replication package: computebeta.m

```
function cmpt=computebeta(iw,Threshold )


cmpt=true;
% first check that all days are available
if sum(isnan(iw))>0
    cmpt=false;
end
if cmpt
    % now check that data exceed threshold
    a=sum(iw>Threshold);
    W=size(iw,1);
    if a==0 % all entries do not exceed threshold
        cmpt=false;
    end
end


return
```
