## Supplementary material for "COVID-19 Time-varying Reproduction Numbers Worldwide: An Empirical Analysis of Mandatory and Voluntary Social Distancing": Replication package: correct_french_data.m

```
 function Ac=correct_french_data(Ac)

cl=country_list(Ac);


%French data from 4/4 to 4/11 from http://hgis.uw.edu/virus/assets/virus.csv
dataUW=[68605, 70478, 74390, 78167, 82048, 86334, 90676, 93790];
% % WHO data https://covid19.who.int/region/euro/country/fr
% dataWHO=[63536, 67757, 69607, 73488, 77225, 81094, 85350, 89683];
for i=1:size(cl,1)
    % find france
    if strcmp(cl(i,1),'France')
        % replace 4/4 to 4/11
        if strcmp(Ac{1,78},'4/4/20')
            if Ac{i+1,77}<Ac{i+1,78}
               'French data appears without errors - not replacing 4/4 to 4/11 data - check if errors in French data was fully corrected by JHU' 
             
            else
                Ac(i+1,78:85)=num2cell(dataUW);
            end
        else
            dbstop;
        end
    end
end
return
```
