## Supplementary material for "COVID-19 Time-varying Reproduction Numbers Worldwide: An Empirical Analysis of Mandatory and Voluntary Social Distancing": Replication package: country_list.m

```
function clist=country_list(A)
% create list of countries 
n=size(A,1)-1;
clist=cell(n,1);
for i=1:n
   if ismissing(A{1+i,1}) | size(A{1+i,1},1)==0  % not a region
       clist(i,1)=A(1+i,2);
   else % is a region of a country
       clist{i,1}=[A{1+i,2},': ',A{1+i,1}];
   end
    
end

return
```
