## Supplementary material for "COVID-19 Time-varying Reproduction Numbers Worldwide: An Empirical Analysis of Mandatory and Voluntary Social Distancing": Replication package: csave.m

```
function ycs=csave(y,s)

[n,T]=size(y);
ycs=NaN(n,T);

for t=1:T
   if sum(s(:,t))==n % all true
      %ycs(:,t)=mean(y(:,t)); 
      a=y(:,t);
      S=(ones(n,n)-eye(n))/(n-1);
      ycs(:,t)=S*a;
   end
end


return
```
