## Supplementary material for "COVID-19 Time-varying Reproduction Numbers Worldwide: An Empirical Analysis of Mandatory and Voluntary Social Distancing": Replication package: east_asia_and_pacific.m

```
function [list, list_exChina]=east_asia_and_pacific()

list={'Australia';'Brunei';'Cambodia';'China';'Korea, South';'Fiji';'Indonesia';'Japan';'Laos';...
     'Malaysia';'Marshall Islands';'Mongolia';'Burma';'New Zealand';'Papua New Guinea';'Philippines';...
     'Samoa';'Singapore';'Solomon Islands';'Thailand';'Timor-Leste';'Vanuatu';'Vietnam'};
    
list_exChina={'Australia';'Brunei';'Cambodia';'Korea, South';'Fiji';'Indonesia';'Japan';'Laos';...
     'Malaysia';'Marshall Islands';'Mongolia';'Burma';'New Zealand';'Papua New Guinea';'Philippines';...
     'Samoa';'Singapore';'Solomon Islands';'Thailand';'Timor-Leste';'Vanuatu';'Vietnam'};
return
```
