## Supplementary material for "COVID-19 Time-varying Reproduction Numbers Worldwide: An Empirical Analysis of Mandatory and Voluntary Social Distancing": Replication package: Eastern_and_Southern_Africa.m

```
function [list]=Eastern_and_Southern_Africa()

list={'Angola';'Botswana';'Burundi';'Comoros';'Djibouti';'Eritrea';'Ethiopia';...
    'Kenya';'Lesotho';'Madagascar';'Malawi';'Mozambique';'Namibia';'Rwanda';'Seychelles';'Somalia';...
    'South Africa';'South Sudan';'Sudan';'Uganda';'Tanzania''Zambia';'Zimbabwe'};


return
```
