## Supplementary material for "COVID-19 Time-varying Reproduction Numbers Worldwide: An Empirical Analysis of Mandatory and Voluntary Social Distancing": Replication package: Eastern_Europe_and_Central_Asia.m

```
function [list]=Eastern_Europe_and_Central_Asia()

list={'Albania';'Armenia';'Azerbaijan';'Belarus';'Bosnia and Herzegovina';'Bulgaria';'Croatia';'Georgia';...
    'Kazakhstan';'Kyrgyzstan';'Montenegro';'Moldova';'Romania';'Russia';'Serbia';'Tajikistan';'North Macedonia';...
    'Turkey';'Ukraine';'Uzbekistan'};


return
```
