## Supplementary material for "COVID-19 Time-varying Reproduction Numbers Worldwide: An Empirical Analysis of Mandatory and Voluntary Social Distancing": Replication package: fill_entries.m

```
function B=fill_entries(A,al,cl)

[~,T]=size(A);
n=size(cl,1);

B=NaN(n,T);
for i=1:n
   ind=strcmp(cl{i},al);
   if(sum(ind))>1
       'more than one country ID found'
       dbstop

   end
   if sum(ind)==1
      B(i,:)=A(ind,:); 
   end
end

return
```
