## Supplementary material for "COVID-19 Time-varying Reproduction Numbers Worldwide: An Empirical Analysis of Mandatory and Voluntary Social Distancing": Replication package: find_go_sp.m

```
function [gm, foundc,foundcd,i]=find_go_sp(Ago,country_to_find,date_to_find, sp, indX)

c1=str2num(datestr(date_to_find, 'yyyymmdd'));

nr=size(Ago,1);
gm=cell(1,1);
gm{1,1}=[]; %missing;
foundc=false;
foundcd=false;

i=sp-1;
while not(foundcd || i>nr-1)
i=i+1;    
     if strcmp(Ago{i,1},country_to_find) % found country
         foundc=true;
         c2=Ago{i,6}; %datestr(Agg{i,8});
         if c1==c2 % found also date
            foundcd=true;
            gm=ox_select(Ago(i,:), indX);
         end
     end
end
         


return
```
