## Supplementary material for "COVID-19 Time-varying Reproduction Numbers Worldwide: An Empirical Analysis of Mandatory and Voluntary Social Distancing": Replication package: find_go_start.m

```
function [sp, foundc]=find_go_start(Ago,country_to_find)

nr=size(Ago,1);
foundc=false;
sp=NaN;
i=1;
while not(foundc || i>nr-1)
i=i+1;    
     if strcmp(Ago{i,1},country_to_find) % found country
         foundc=true;
         sp=i;
     end
end
         


return
```
