## Supplementary material for "COVID-19 Time-varying Reproduction Numbers Worldwide: An Empirical Analysis of Mandatory and Voluntary Social Distancing": Replication package: hemispheres.m

```
function [SHlist,NHlist]=hemispheres(worldlist, Ac)

n=size(worldlist,1);
cl=country_list(Ac);
na=size(cl,1);
SHlist={};sh=0;
NHlist={};nh=0;

for j=1:n
    % find country position
    ind=false(na,1);
     for i=1:na
        if strcmp(cl{i,1},worldlist{j,1}) 
            ind(i+1,1)=true;
        end
    end
    lat=Ac{ind,3};
    if lat<0
        sh=sh+1;
        SHlist{sh,1}=worldlist{j,1};
    else
        nh=nh+1;
        NHlist{nh,1}=worldlist{j,1};
    end
end


return
```
