## Supplementary material for "COVID-19 Time-varying Reproduction Numbers Worldwide: An Empirical Analysis of Mandatory and Voluntary Social Distancing": Replication package: Latin_America_and_Caribbean.m

```
function [list]=Latin_America_and_Caribbean()

list={'Antigua and Barbuda';'Argentina';'Bahamas';'Barbados';'Belize';'Bolivia';'Brazil';'Chile';'Colombia';...
    'Costa Rica';'Cuba';'Dominica';'Dominican Republic';'Ecuador';'El Salvador';'Grenada';'Guatemala';...
    'Guyana';'Haiti';'Honduras';'Jamaica';'Mexico';'Nicaragua';'Panama';'Paraguay';'Peru';...
    'Saint Kitts and Nevis';'Saint Lucia';'Saint Vincent and the Grenadines';'Suriname';...
    'Trinidad and Tobago';'Uruguay';'Venezuela'};


return
```
