## Supplementary material for "COVID-19 Time-varying Reproduction Numbers Worldwide: An Empirical Analysis of Mandatory and Voluntary Social Distancing": Replication package: MGestim.m

```
function [A, ycs, s]=MGestim(c, i, I,MAtransform, sv, Xo, lags, country_names, MF, last_is_csave, Xnames)


gamma=1/14;
nl=size(lags,2);

[n,T]=size(c);
nindex=[1:n]'*ones(1,T);
Tindex=ones(n,1)*[1:T];
%% MA transformation
if MAtransform>0 % compute 7-day ma
    ima=zeros(n,T);
    cma=zeros(n,T);
    Ima=zeros(n,T);
    %sma=zeros(n,T);
    for t=MAtransform:T
        ima(:,t)=i(:,t-MAtransform+1:t)*ones(MAtransform,1)/MAtransform;
        cma(:,t)=c(:,t-MAtransform+1:t)*ones(MAtransform,1)/MAtransform;
        Ima(:,t)=I(:,t-MAtransform+1:t)*ones(MAtransform,1)/MAtransform;
       % sma(:,t)=s(:,t-6:t)*ones(7,1)/7;
    end
    i=ima;
    c=cma;
    I=Ima;
end


%% individual regressions
% define cell structure for saving
A=cell(10,10);
% if last_is_csave
%     A(1,1)={['Individual country and Mean Group estimation results using specifications augmented by cross section averages (domestic country is excluded from CS average) and using Multiplication Factor MF=',int2str(MF)]};
% else
%     A(1,1)={['Individual country and Mean Group estimation results using specifications NOT augmented by CS averages, and using Multiplication Factor MF=',int2str(MF)]};
% 
% end

for ell=1:nl
 lagorder=lags(1,ell);
 k=size(Xo,3);
 X=lagx(Xo,lagorder*ones(k,1));
 
 sh=(ell-1)*(n+7);
 if last_is_csave
  %A(3+sh,1)={['Individual country estimation results using specifications augmented by CS averages, lag order p=',int2str(lagorder),' and multiplication factor MF=',int2str(MF)]};   
  A(3+sh,1)={['Individual country estimation results using specifications augmented by 7-day lagged dependent variable for Western Europe aggregate, lag order p=',int2str(lagorder),' and multiplication factor MF=',int2str(MF)]};   

 else
  %A(3+sh,1)={['Individual country estimation results using specifications not augmented by CS averages, lag order p=',int2str(lagorder),' and multiplication factor MF=',int2str(MF)]};
  A(3+sh,1)={['Individual country estimation results using specifications not augmented by Western Europe aggregate, lag order p=',int2str(lagorder),' and multiplication factor MF=',int2str(MF)]};
  end
 A(5+sh,1)={'Country'};
 A(4+sh,2)={'Constant'};A(4+sh,3)={'robust s.e.'};A(4+sh,4)={'robust t-ratio'};
                        A(4+sh,5)={'standard s.e.'};A(4+sh,6)={'standard t-ratio'};
 for j=1:k
     A(4+sh,2+j*5)=Xnames(j,1);
     A(4+sh,3+j*5)={'robust s.e.'};  A(4+sh,4+j*5)={'robust t-ratio'};
     A(4+sh,5+j*5)={'standard s.e.'};A(4+sh,6+j*5)={'standard t-ratio'};
     if last_is_csave && j==k
         % A(4+sh,2+j*5)={'CS average of dep. var'};
         A(4+sh,2+j*5)={'Western Europe dep. var. lagged by 7 days'};
     end
 end
 A(4+sh,7+k*5)={'R2'};A(4+sh,8+k*5)={'T'};
 
 
 %% determine sample
 % first sufficient I>sv exists
 s1=I>sv;
 % next regressors exist

 s2=true(size(c));
 for j=1:k
     s2=s2 & not(isnan(X(:,:,j)));
 end
 %% dependent variable
 y=NaN(size(c));
 y(:,1:end-1)=-log((1-c(:,2:end))./(1-c(:,1:end-1)))./i(:,1:end-1);
 sy=not(isnan(y));
 s=s1&s2&sy;
   % compute CS ave
   scs=sy&s1;
   ycs=csave(y/gamma*100,scs );
   
 for ii=1:n
    yi=y(ii,:)';
    Xi=squeeze(X(ii,:,:));
    si=s(ii,:)';
    
    yi=yi(si)/gamma;
    Xi=Xi(si,:);
    % now country estimations
    Te=size(Xi,1);
    Xai=[ones(Te,1),Xi/100];
    b=Xai\yi;
    ba(ii,:)=b';
    mdl = fitlm([Xi/100],yi);
    Q=inv(Xai'*Xai);
    u=yi-Xai*b;
    sb=diag(Q*(u'*u)/Te).^0.5;
    
    % now compute panel NW standard errors
    k=size(Xai,2);    
    Q=zeros(k,k);
    Sa=zeros(k,k);
    T=size(Xai,1);
    m=floor(T^(1/3));
    Ti=size(Xai,1);
    Qi=(Xai'*Xai)/Ti;
    Q=Q+Qi;
    % now Si
    Xui=Xai.*(u*ones(1,k));
    Om=(Xui'*Xui)/Ti;
    Si=Om;
    for ell=1:m
       % now Om,i,ell
       Xuic=Xui(ell+1:end,:);
       Xuil=Xui(1:end-ell,:);
       Omiell= (Xuic'*Xuil)/Ti;
       w=1-ell/(m+1);
       Si=Si+w*(Omiell+Omiell');
    end
    Sa=Sa+Si;
    iQ=inv(Q);
    V=iQ*Sa*iQ'/T;
    seb=diag(V).^0.5;
    % save country results into A
    A(4+ii+sh,1)=country_names(ii,1);
    for j=1:k
        A(4+ii+sh,j*5-3)={b(j,1)}; A(4+ii+sh,j*5-2)={seb(j,1)}; A(4+ii+sh,j*5-1)={b(j,1)/seb(j,1)};
                               A(4+ii+sh,j*5)={sb(j,1)}; A(4+ii+sh,j*5+1)={b(j,1)/sb(j,1)};
    end
    A(4+ii+sh,k*5+2)={mdl.Rsquared.Ordinary};
    A(4+ii+sh,k*5+3)={Te};
 end % ii
 % now MG estimations
  if last_is_csave
    %A(7+n+sh,1)={['Mean Group estimation using specifications augmented by CS averages, lag order p=',int2str(lagorder),' and multiplication factor MF=',int2str(MF)]};
    A(7+n+sh,1)={['Mean Group estimation using specifications augmented by Western Europe dependent variable (7-day lag), lag order p=',int2str(lagorder),' and multiplication factor MF=',int2str(MF)]};

  else
    %A(7+n+sh,1)={['Mean Group estimation using specifications not augmented by CS averages, lag order p=',int2str(lagorder),' and multiplication factor MF=',int2str(MF)]};  
    A(7+n+sh,1)={['Mean Group estimation using specifications not augmented by Western Europe, lag order p=',int2str(lagorder),' and multiplication factor MF=',int2str(MF)]};  
  end
  
    A(6+n+sh,2)={'Constant'};A(6+n+sh,3)={'s.e.'};A(6+n+sh,4)={'t-ratio'};
    bmg=mean(ba);
    vm=ba-ones(n,1)*bmg;
    semg=(diag((vm'*vm/n)/n)).^0.5;
    
    A(7+n+sh,2)={bmg(1,1)}; A(7+n+sh,3)={semg(1,1)};A(7+n+sh,4)={bmg(1,1)/semg(1,1)};
    for j=1:k-1
     A(6+n+sh,2+j*5)={['X',int2str(j)]};
     A(7+n+sh,2+j*5)={bmg(1,j+1)};
     A(6+n+sh,3+j*5)={'s.e.'};  A(6+n+sh,4+j*5)={'t-ratio'};
     A(7+n+sh,3+j*5)={semg(j+1,1)};  A(7+n+sh,4+j*5)={bmg(1,j+1)/semg(j+1,1)};
        if last_is_csave && j==k-1
            %A(6+n+sh,2+j*5)={'CS average of dep. var'};
            A(6+n+sh,2+j*5)={'Western Europe dep. var. lagged 7 days'};
        end
    end
    A(9+n+sh,1)={''};                    
end % ell

return
```
