## Supplementary material for "COVID-19 Time-varying Reproduction Numbers Worldwide: An Empirical Analysis of Mandatory and Voluntary Social Distancing": Replication package: Middle_East_and_North_Africa.m

```
function [list]=Middle_East_and_North_Africa()

list={'Algeria';'Bahrain';'Egypt';'Iran';'Iraq';'Israel';'Jordan'; 'Kuwait';'Lebanon';...
    'Libya';'Morocco';'Oman';'Qatar';'Saudi Arabia';'Syria';'Tunisia';'United Arab Emirates';'Yemen'};


return
```
