## Supplementary material for "COVID-19 Time-varying Reproduction Numbers Worldwide: An Empirical Analysis of Mandatory and Voluntary Social Distancing": Replication package: panel_estimations.m

```
function panel_estimations(SC,D, Thvar, sv, save_file, ycsind, do_thresholds)


% preliminaries
lags=[14,21]; % lag orders
ma=0; % moving average transformation of data - set 0 for none

c=D.c;
i2=D.i2;
I2=D.I2;
X=D.X;
Ac=D.Ac;
Xnames=D.Xnames;

% select sample
[csel,isel,Isel,Xsel,Rsel, Acsel,Thsel.data ]=select_sample(c,i2,I2,X,D.R,SC.list,Ac,Thvar.data);
Thsel.name=Thvar.name;
Thsel.grid=Thvar.grid;
Thsel.usegrid=Thvar.usegrid;

if ycsind==1
    ycs=select_AGGforCSave(c,i2,I2,{'Western Europe'}, SC.list, Ac, sv)*100; 
    csname={'Western Europe d.v. (7-day lag)'};
end
if ycsind==2
    ycs=select_AGGforCSave(c,i2,I2,{'World excl. China'}, SC.list, Ac, sv)*100; 
    csname={'World excl. China d.v. (7-day lag)'};
end

% first individual estimations and MG
MF=5; last_is_csave=false;
[A1, ycs1, s]=MGestim(MF*csel, MF*isel,MF*Isel, ma, MF*sv, Xsel,lags, SC.list, MF, last_is_csave, Xnames);


MF=3;
A2=MGestim(MF*csel, MF*isel,MF*Isel, ma, MF*sv, Xsel,lags, SC.list, MF, last_is_csave, Xnames);
% now with CS averages
MF=5; last_is_csave=true;
Xselcs=Xsel;
Xselcs(:,:,end+1)=ycs;
XnamesCS=Xnames;
XnamesCS(end+1,1)=csname;
A3=MGestim(MF*csel, MF*isel,MF*Isel, ma, MF*sv, Xselcs,lags, SC.list, MF,last_is_csave, XnamesCS);
MF=3;
A4=MGestim(MF*csel, MF*isel,MF*Isel, ma, MF*sv, Xselcs,lags, SC.list, MF, last_is_csave, XnamesCS);

% save A in xls
disp('saving individual and MG estimation results');
Aa=[A1;A2];
Ab=[A3;A4];k=size(Ab,2)-size(Aa,2);
Aa(:,end+1:end+k)={''};
A=[Aa;Ab];
writecell(A,save_file, 'Sheet', 'Individual and MG', 'Range', 'A1');

%% now pooled estimation without threshold

% common intercept, no CS

[n,T,k]=size(Xsel);
Xcommon=NaN(n,T,k+1);
Xcommon(:,:,1:end-1)=Xsel/100;
Xcommon(:,:,end)=1;
Xspec=NaN(n,T,0);
regressor_names.common(1:k,1)=Xnames;
regressor_names.common(k+1,1)={['Intercept']};
regressor_names.specific={};
LX.common=[true(size(Xnames,1),1);false];
LX.spec=[];

MF=5; 
A1=pooled_estim(MF*csel, MF*isel,MF*Isel, ma, MF*sv, Xcommon, Xspec, lags, LX,MF,regressor_names, SC.list);
MF=3; 
A2=pooled_estim(MF*csel, MF*isel,MF*Isel, ma, MF*sv, Xcommon, Xspec, lags, LX,MF,regressor_names, SC.list);

% country-specific intercept, no CS
Xcommon=Xsel/100;
Xspec=ones(n,T,1); %createFE(n,T);
regressor_names.common=Xnames;
regressor_names.specific={'Fixed Effects'};
LX.common=true(size(Xnames,1),1);
LX.spec=[false];

MF=5; 
A3=pooled_estim(MF*csel, MF*isel,MF*Isel, ma, MF*sv, Xcommon, Xspec, lags, LX,MF,regressor_names, SC.list);
MF=3; 
A4=pooled_estim(MF*csel, MF*isel,MF*Isel, ma, MF*sv, Xcommon, Xspec, lags, LX,MF,regressor_names, SC.list);

% common intercept, pooled CS

Xcommon=Xsel/100;
Xcommon(:,:,end+1)=1;
Xcommon(:,:,end+1)=ycs/100;
Xspec=NaN(n,T,0);
regressor_names.common(1:k,1)=Xnames;
regressor_names.common(k+1,1)={['Intercept']};
regressor_names.common(k+2,1)={['Western Europe d.v. (7-day lag)']};
regressor_names.specific={};
LX.common=[true(size(Xnames,1),1); false; false];
LX.spec=[];

MF=5; 
A5=pooled_estim(MF*csel, MF*isel,MF*Isel, ma, MF*sv, Xcommon, Xspec, lags, LX,MF,regressor_names, SC.list);
MF=3; 
A6=pooled_estim(MF*csel, MF*isel,MF*Isel, ma, MF*sv, Xcommon, Xspec, lags, LX,MF,regressor_names, SC.list);

% country-specific intercept, het CS
Xcommon=Xsel/100;
Xspec=ones(n,T,1); %createFE(n,T);
Xspec(:,:,end+1)=ycs/100;
regressor_names.common=Xnames;
regressor_names.specific={'Fixed Effects';['Western Europe d.v. (7-day lag)']};
LX.common=true(size(Xnames,1),1);
LX.spec=[false; false];

MF=5; 
A7=pooled_estim(MF*csel, MF*isel,MF*Isel, ma, MF*sv, Xcommon, Xspec, lags, LX,MF,regressor_names, SC.list);
MF=3; 
A8=pooled_estim(MF*csel, MF*isel,MF*Isel, ma, MF*sv, Xcommon, Xspec, lags, LX,MF,regressor_names, SC.list);

A=[A1;A2;A3;A4;A5;A6;A7;A8];
disp('saving Pooled results without threshold');
writecell(A,save_file, 'Sheet', 'Pooled without threshold', 'Range', 'A1');

%% now pooled regressions with single threshold

if do_thresholds
% common intercept, no CS

[n,T,k]=size(Xsel);
Xcommon=NaN(n,T,k+1);
Xcommon(:,:,1:end-1)=Xsel/100;
Xcommon(:,:,end)=1;
Xspec=NaN(n,T,0);
regressor_names.common(1:k,1)=Xnames;
regressor_names.common(k+1,1)={['Intercept']};
regressor_names.specific={};
LX.common=[true(size(Xnames,1),1);false];
LX.spec=[];

MF=5; 
A1=pooled_estim_threshold(MF*csel, MF*isel,MF*Isel, ma, MF*sv, Xcommon, Xspec, lags, LX,MF,regressor_names, SC.list, Thsel);
MF=3; 
A2=pooled_estim_threshold(MF*csel, MF*isel,MF*Isel, ma, MF*sv, Xcommon, Xspec, lags, LX,MF,regressor_names, SC.list, Thsel);

% country-specific intercept, no CS
Xcommon=Xsel/100;
Xspec=ones(n,T,1); %createFE(n,T);
regressor_names.common=Xnames;
regressor_names.specific={'Fixed Effects'};
LX.common=[true(size(Xnames,1),1)];
LX.spec=[false];

MF=5; 
A3=pooled_estim_threshold(MF*csel, MF*isel,MF*Isel, ma, MF*sv, Xcommon, Xspec, lags, LX,MF,regressor_names, SC.list, Thsel);
MF=3; 
A4=pooled_estim_threshold(MF*csel, MF*isel,MF*Isel, ma, MF*sv, Xcommon, Xspec, lags, LX,MF,regressor_names, SC.list, Thsel);

% common intercept, pooled CS

Xcommon=Xsel/100;
Xcommon(:,:,end+1)=1;
Xcommon(:,:,end+1)=ycs/100;
Xspec=NaN(n,T,0);
regressor_names.common(1:k,1)=Xnames;
regressor_names.common(k+1,1)={['Intercept']};
regressor_names.common(k+2,1)={['Western Europe d.v. (7-day lag)']};
regressor_names.specific={};
LX.common=[true(size(Xnames,1),1); false; false];
LX.spec=[];

MF=5; 
A5=pooled_estim_threshold(MF*csel, MF*isel,MF*Isel, ma, MF*sv, Xcommon, Xspec, lags, LX,MF,regressor_names, SC.list, Thsel);
MF=3; 
A6=pooled_estim_threshold(MF*csel, MF*isel,MF*Isel, ma, MF*sv, Xcommon, Xspec, lags, LX,MF,regressor_names, SC.list, Thsel);

% country-specific intercept, het CS
Xcommon=Xsel/100;
Xspec=ones(n,T,1); %createFE(n,T);
Xspec(:,:,end+1)=ycs/100;
regressor_names.common=Xnames;
regressor_names.specific={'Fixed Effects';['Western Europe d.v. (7-day lag)']};
LX.common=[true(size(Xnames,1),1)];
LX.spec=[false; false];

MF=5; 
A7=pooled_estim_threshold(MF*csel, MF*isel,MF*Isel, ma, MF*sv, Xcommon, Xspec, lags, LX,MF,regressor_names, SC.list, Thsel);
MF=3; 
A8=pooled_estim_threshold(MF*csel, MF*isel,MF*Isel, ma, MF*sv, Xcommon, Xspec, lags, LX,MF,regressor_names, SC.list, Thsel);

A=[A1;A2;A3;A4;A5;A6;A7;A8];
disp('saving Pooled results with single threshold');
writecell(A,save_file, 'Sheet', 'Pooled with single threshold', 'Range', 'A1');

% %% now pooled regressions with two thresholds
% 
% Period1=false(1,T); Period1(1,1:131)=true; % start of sample until end May)
% Period2=false(1,T); Period2(1,132:end)=true; % start of sample until end May)
% 
% Thmulti{1}.data=zeros(size(Thsel.data));
% Thmulti{1}.data(:,Period1)=Thsel.data(:,Period1);
% Thmulti{1}.name='Period 1 (until end May) confirmed cases per day per 100k population (7dma)';
% Thmulti{2}.data=zeros(size(Thsel.data));
% Thmulti{2}.data(:,Period2)=Thsel.data(:,Period2);
% Thmulti{2}.name='Period 2 (from the begining of June) confirmed cases per day per 100k population (7dma)';
% 
% Thmulti{1}.grid=Thsel.grid(1:55); % max out at 10, about 98% quantile
% Thmulti{1}.usegrid=Thsel.usegrid;
% Thmulti{2}.grid=Thsel.grid;
% 
% % common intercept, no CS
% 
% [n,T,k]=size(Xsel);
% Xcommon=NaN(n,T,k+1);
% Xcommon(:,:,1:end-1)=Xsel/100;
% Xcommon(:,:,end)=1;
% Xspec=NaN(n,T,0);
% regressor_names.common(1:k,1)=Xnames;
% regressor_names.common(k+1,1)={['Intercept']};
% regressor_names.specific={};
% LX.common=[true(size(Xnames,1),1);false];
% LX.spec=[];
% 
% MF=5; 
% A1=pooled_estim_threshold2(MF*csel, MF*isel,MF*Isel, ma, MF*sv, Xcommon, Xspec, lags, LX,MF,regressor_names, SC.list, Thmulti);
% 'a1 th2 done';
% MF=3; 
% A2=pooled_estim_threshold2(MF*csel, MF*isel,MF*Isel, ma, MF*sv, Xcommon, Xspec, lags, LX,MF,regressor_names, SC.list, Thmulti);
% 
% % country-specific intercept, no CS
% Xcommon=Xsel/100;
% Xspec=ones(n,T,1); %createFE(n,T);
% regressor_names.common=Xnames;
% regressor_names.specific={'Fixed Effects'};
% LX.common=[true(size(Xnames,1),1)];
% LX.spec=[false];
% 
% MF=5; 
% A3=pooled_estim_threshold2(MF*csel, MF*isel,MF*Isel, ma, MF*sv, Xcommon, Xspec, lags, LX,MF,regressor_names, SC.list, Thmulti);
% MF=3; 
% A4=pooled_estim_threshold2(MF*csel, MF*isel,MF*Isel, ma, MF*sv, Xcommon, Xspec, lags, LX,MF,regressor_names, SC.list, Thmulti);
% 
% % common intercept, pooled CS
% 
% Xcommon=Xsel/100;
% Xcommon(:,:,end+1)=1;
% Xcommon(:,:,end+1)=ycs/100;
% Xspec=NaN(n,T,0);
% regressor_names.common(1:k,1)=Xnames;
% regressor_names.common(k+1,1)={['Intercept']};
% regressor_names.common(k+2,1)={['Western Europe d.v. (7-day lag)']};
% regressor_names.specific={};
% LX.common=[true(size(Xnames,1),1); false; false];
% LX.spec=[];
% 
% MF=5; 
% A5=pooled_estim_threshold2(MF*csel, MF*isel,MF*Isel, ma, MF*sv, Xcommon, Xspec, lags, LX,MF,regressor_names, SC.list, Thmulti);
% MF=3; 
% A6=pooled_estim_threshold2(MF*csel, MF*isel,MF*Isel, ma, MF*sv, Xcommon, Xspec, lags, LX,MF,regressor_names, SC.list, Thmulti);
% 
% % country-specific intercept, het CS
% Xcommon=Xsel/100;
% Xspec=ones(n,T,1); %createFE(n,T);
% Xspec(:,:,end+1)=ycs/100;
% regressor_names.common=Xnames;
% regressor_names.specific={'Fixed Effects';['Western Europe d.v. (7-day lag)']};
% LX.common=true(size(Xnames,1),1);
% LX.spec=[false; false];
% 
% MF=5; 
% A7=pooled_estim_threshold2(MF*csel, MF*isel,MF*Isel, ma, MF*sv, Xcommon, Xspec, lags, LX,MF,regressor_names, SC.list, Thmulti);
% MF=3; 
% A8=pooled_estim_threshold2(MF*csel, MF*isel,MF*Isel, ma, MF*sv, Xcommon, Xspec, lags, LX,MF,regressor_names, SC.list, Thmulti);
% 
% A=[A1;A2;A3;A4;A5;A6;A7;A8];
% disp('saving Pooled results with two thresholds');
% writecell(A,save_file, 'Sheet', 'Pooled with two thresholds', 'Range', 'A1');

end % if dothresholds
return
```
