## Supplementary material for "COVID-19 Time-varying Reproduction Numbers Worldwide: An Empirical Analysis of Mandatory and Voluntary Social Distancing": Replication package: pooled_estim.m

```
function [A, ycs]=pooled_estim(c, i, I,MAtransform, sv, Xcommon, Xspec,lags, LX, MF, regressor_names,country_names)


gamma=1/14;
nl=size(lags,2);

[n,T]=size(c);
nindex=[1:n]'*ones(1,T);
Tindex=ones(n,1)*[1:T];
%% MA transformation
if MAtransform>0 % compute 7-day ma
    ima=zeros(n,T);
    cma=zeros(n,T);
    Ima=zeros(n,T);
    %sma=zeros(n,T);
    for t=MAtransform:T
        ima(:,t)=i(:,t-MAtransform+1:t)*ones(MAtransform,1)/MAtransform;
        cma(:,t)=c(:,t-MAtransform+1:t)*ones(MAtransform,1)/MAtransform;
        Ima(:,t)=I(:,t-MAtransform+1:t)*ones(MAtransform,1)/MAtransform;
       % sma(:,t)=s(:,t-6:t)*ones(7,1)/7;
    end
    i=ima;
    c=cma;
    I=Ima;
end


%% individual regressions
% define cell structure for saving
A=cell(10,10);
kc=size(Xcommon,3);
pooled_regresors=[''];
for ii=1:kc
    if ii<kc
        pooled_regresors=[pooled_regresors,' ',regressor_names.common{ii,1},','];
    else
        pooled_regresors=[pooled_regresors,' ',regressor_names.common{ii,1},'.'];
    end
end
ks=size(Xspec,3);
if ks==0
    regresors_with_country_specific_slopes=['None.'];
else
    regresors_with_country_specific_slopes=[''];
    for ii=1:ks
        if ii<ks
            regresors_with_country_specific_slopes=[regresors_with_country_specific_slopes,' ',regressor_names.specific{ii,1},','];
        else
            regresors_with_country_specific_slopes=[regresors_with_country_specific_slopes,' ',regressor_names.specific{ii,1},'.'];
        end
    end
end

for ell=1:nl
 lagorder=lags(1,ell);
 lagscommon=zeros(kc,1);
 for ii=1:kc
     if LX.common(ii)
         lagscommon(ii,1)=lagorder;
     end
 end
 Xcl=lagx(Xcommon,lagscommon);
 
lagsspec=zeros(ks,1);
 if ks>0
  for ii=1:ks
     if LX.spec(ii)
         lagsspec(ii,1)=lagorder;
     end
 end
 Xsl=lagx(Xspec,lagsspec);
 Xslcc=create_country_specific_vars(Xsl);
 else
     Xslcc=Xspec;
 end
X=NaN(n,T, kc+ks);
X(:,:,1:kc)=Xcl;
X(:,:,kc+1:kc+ks*n)=Xslcc;
 
 
 sh=(ell-1)*(9+kc+n*ks);
 A(1+sh,1)={['Pooled panel estimation results. Multiplication Factor MF=',int2str(MF),...
          '. Lag order=', int2str(lagorder), ... 
          '. Pooled regressors: ',pooled_regresors, ...
          ' Regressors with country-specific slopes: ', regresors_with_country_specific_slopes] };

 for ii=1:kc
     if LX.common(ii)
        A(2+sh+ii,1)={[regressor_names.common{ii,1}, ' (lagged ', int2str(lagorder), 'days)']};
     else
        A(2+sh+ii,1)=regressor_names.common(ii,1);
     end
 end
 
 A(2+sh,2)={'Coeficient estimate'}; 
 A(2+sh,3)={'standard s.e.'};
 A(2+sh,4)={'standard t-ratio'}; 
 A(2+sh,5)={'robust s.e. (serial correlation)'};
 A(2+sh,6)={'robust t-ratio (serial correlation)'};
 A(2+sh,7)={'robust s.e. (serial and CS correlation)'};
 A(2+sh,8)={'robust t-ratio (serial and CS correlation)'};

  
 if ks>0
  for j=1:ks
     for jj=1:n 
        A(2+kc+sh+jj+(j-1)*n,1)={[regressor_names.specific{j},': ',country_names{jj}]};
     end
  end
 end
 A(3+kc+n*ks+sh,1)={'R2'};
 A(4+kc+n*ks+sh,1)={'No. of observations'};
 A(5+kc+n*ks+sh,1)={'n'};  A(5+kc+n*ks+sh,2)={int2str(n)};
 A(6+kc+n*ks+sh,1)={'Tmin'};
 A(7+kc+n*ks+sh,1)={'Tmax'};
 
 %% determine sample
 % first sufficient I>sv exists
s1=I>sv;
% next regressors exist
k=size(X,3);
s2=true(size(c));
for j=1:k
    s2=s2 & not(isnan(X(:,:,j)));
end


%% dependent variable
y=NaN(size(c));
y(:,1:end-1)=-log((1-c(:,2:end))./(1-c(:,1:end-1)))./i(:,1:end-1);


sy=not(isnan(y));
s=s1&s2&sy;

% stack dependent and explanatory variables, and sample selector
ys=reshape(y',n*T,1);
Xs=reshape(permute(X,[2,1,3]),n*T,k);
ss=reshape(s',n*T,1);
ninds=reshape(nindex',n*T,1);
Tinds=reshape(Tindex',n*T,1);

gamma=1/14;
yr=ys(ss)/gamma;
Xr=Xs(ss,:);
nr=ninds(ss,:);
Tr=Tinds(ss,:);

Te=size(Xr,1);
b=[Xr]\yr;
mdl = fitlm([Xr],yr,'Intercept',false);
R=mdl;
u=R.Residuals{1:end,1};
S.data_and_resid=[nr,Tr,yr,Xr,u];
%S.datas=[ninds,Tinds,ys,Xs];

% for checking
Xall=[Xr];
%inv(Xall'*Xall)*Xall'*yr
Q=inv(Xall'*Xall);
u=yr-Xall*b;
sb=diag(Q*(u'*u)/Te).^0.5;

%% now compute panel NW standard errors
kpm=1/3;
    % compute Q and Sa
Xall=[Xr];
k=size(Xall,2);    
 Q=zeros(k,k);
 Qn=zeros(k,k);
 Sa=zeros(k,k);
 nT=size(Xall,1);
Tiall=zeros(n,1);

for ii=1:n
    % get Xi
    %ind=true(nT,1);
    ind=nr==ii; %
    Xi=Xall(ind,:);
    Ti=size(Xi,1);
    Tiall(ii,1)=Ti;
    Qi=(Xi'*Xi)/Ti;
    
    Q=Q+Qi*(Ti/nT);
    Qn=Qn+Qi*Ti;
     m=floor(Ti^kpm);
    % now Si
    ui=u(ind,1);
    Xui=Xi.*(ui*ones(1,k));
    Om=(Xui'*Xui)/Ti;
    Si=Om;
    for el=1:m
       % now Om,i,ell
       Xuic=Xui(el+1:end,:);
       Xuil=Xui(1:end-el,:);
       Omiell= (Xuic'*Xuil)/Ti;
       w=1-el/(m+1);
       Si=Si+w*(Omiell+Omiell');
    end
    Sa=Sa+Si*(Ti/nT);
end
iQ=inv(Q);
V=iQ*Sa*iQ'/nT;
seb=diag(V).^0.5;

S.seb=seb;


%% now compute s.e's robust to cs and serial correlation


% get ht
Tmin=min(Tr);
Tmax=max(Tr);
H=zeros(Tmax-Tmin+1,k);
for t=Tmin:Tmax
    ind=Tr==t; %
    Xt=Xall(ind,:);
    ut=u(ind,1);
    xut=Xt.*(ut*ones(1,k));
    ht=sum(xut);
    H(t-Tmin+1,:)=ht';
end

% now n-w
m=floor((Tmax-Tmin+1)^kpm);
    Om=(H'*H);
    SS=Om;
    for el=1:m
       % now Om,i,ell
       Hc=H(el+1:end,:);
       Hl=H(1:end-el,:);
       Omiell= (Hc'*Hl);
       w=1-el/(m+1);
       SS=SS+w*(Omiell+Omiell');
    end
iQn=inv(Qn);
Vn=iQn*SS*iQn';
sec=diag(Vn).^0.5;    


%% save results in A
for ii=1:kc
    A(2+sh+ii,2)={b(ii)};    
    A(2+sh+ii,3)={sb(ii)};
    A(2+sh+ii,4)={b(ii)/sb(ii)};
    A(2+sh+ii,5)={seb(ii)};
    A(2+sh+ii,6)={b(ii)/seb(ii)};
    A(2+sh+ii,7)={sec(ii)};
    A(2+sh+ii,8)={b(ii)/sec(ii)};

end
for ii=1:ks
    for jj=1:n
        A(2+sh+kc+(ii-1)*n+jj,2)={b(kc+(ii-1)*n+jj)};
        A(2+sh+kc+(ii-1)*n+jj,3)={sb(kc+(ii-1)*n+jj)};
        A(2+sh+kc+(ii-1)*n+jj,4)={b(kc+(ii-1)*n+jj)/sb(kc+(ii-1)*n+jj)};
        A(2+sh+kc+(ii-1)*n+jj,5)={seb(kc+(ii-1)*n+jj)};
        A(2+sh+kc+(ii-1)*n+jj,6)={b(kc+(ii-1)*n+jj)/seb(kc+(ii-1)*n+jj)};
        A(2+sh+kc+(ii-1)*n+jj,7)={sec(kc+(ii-1)*n+jj)};
        A(2+sh+kc+(ii-1)*n+jj,8)={b(kc+(ii-1)*n+jj)/sec(kc+(ii-1)*n+jj)};
    end
end
 
 A(3+kc+n*ks+sh,2)={R.Rsquared.Ordinary};
 A(4+kc+n*ks+sh,2)={nT};
 A(5+kc+n*ks+sh,2)={int2str(n)};
 A(6+kc+n*ks+sh,2)={int2str(min(Tiall))};
 A(7+kc+n*ks+sh,2)={int2str(max(Tiall))};
 A(9+kc+n*ks+sh,1)={''};
end % ell

return
```
