## Supplementary material for "COVID-19 Time-varying Reproduction Numbers Worldwide: An Empirical Analysis of Mandatory and Voluntary Social Distancing": Replication package: ReadMe.txt

This subfolder contains Matlab codes for the panel data estimations reported in Section 5 of "COVID-19 Time-varying Reproduction Numbers Worldwide: An Empirical Analysis of Mandatory and Voluntary Social Distancing," by A. Chudik, M. H. Pesaran and A. Rebucci, 15 March 2021. 

The script to run is: "RunMe.m". This script loads data from the following files:

- confirmed_cases_txt (Covid-19 data)
- population.xlsx (Population data)
- OxCGRT.csv (Oxford indices).

Panel estimation results are saved in Excel file panel_estimation_results_Europe.xlsx. Sheets Table1, Table2, and Table3, contain linked estimation results reported in Tables 1-3 of the paper, respectively.
