## Supplementary material for "COVID-19 Time-varying Reproduction Numbers Worldwide: An Empirical Analysis of Mandatory and Voluntary Social Distancing": Replication package: reorderAP.m

```
function Apn=reorderAP(Ap,Ac)
cla=country_list(Ap);
clc=country_list(Ac);

na=size(cla,1);
nc=size(clc,1);
Apn(1,:)=Ap(1,:);

for i=1:nc
   pom=0;
   for j=1:na 
        if strcmp(cla{j,1},clc{i,1})
            pom=pom+1;
            Apn(i+1,:)=Ap(j+1,:);
        end
   end
   if pom==0
       Apn(i+1,1:3)=Ac(i+1,1:3);
   end
   if pom>1
       'error'
       dbstop
   end
end
return
```
