## Supplementary material for "COVID-19 Time-varying Reproduction Numbers Worldwide: An Empirical Analysis of Mandatory and Voluntary Social Distancing": Replication package: run_me.m

```
%% This script replicates panel data estimation results in Section 5 of:
% "COVID-19 Time-varying Reproduction Numbers Worldwide: An Empirical Analysis
% of Mandatory and Voluntary Social Distancing," by A. Chudik, M. H. Pesaran and A. Rebucci, 15 March 2021. 
%
% Data is loaded first, using the following input files: 
% - Covid-19 data in confirmed_cases_txt, 
% - Population data in population.xlsx, and
% - Oxford indices data in OxCGRT.csv.
%
% Panel estimation results will be saved in Excel file save_file_europe='panel_estimation_results_Europe.xlsx' specified below.
% See also ReadMe.txt for further information.

clc 
clear
%% load data

    disp('loading covid-19 data from local txt file...')
 % covid-19 data   
 Ac = readcell('confirmed_cases_txt', 'DatetimeType','text');
 % population data
 disp('loading population data from xlsx file...')
 Ap = readcell('population.xlsx');
  % delete missing population entries
  j=2;
  for i=2:size(Ap,1)
      if ismissing(Ap{j,5})
          Ap(j,:)=[];
      else
          j=j+1;
      end
  end
Ap=reorderAP(Ap,Ac); % make sure order is identical;

Ac=correct_french_data(Ac); % correct the obvious data error in JHU French data in the period 4/4 to 4/11 
  
% add unreported country aggregates to Ac
Ac=add_aggregates(Ac);
Ap=add_aggregates(Ap);

 
 % Oxford data
   disp('loading Oxford indices data from csv file...');
  Ago = readcell('OxCGRT.csv');
  % clear regions data to save space
  ind=false(size(Ago,1),1);
  for i=2:size(Ago,1)
      a=Ago{i,3};
      if size(a,2)>1
        ind(i,1)=true;%not(ismissing(a));
      end
  end
   Ago(ind,:)=[];
  % replace names to match JUH data labels
  Ago=replace_nameso(Ago,'South Korea','Korea, South');
  Ago=replace_nameso(Ago,'Myanmar','Burma');
  Ago=replace_nameso(Ago,'Democratic Republic of Congo','Congo (Kinshasa)');
  Ago=replace_nameso(Ago,'Congo','Congo (Brazzaville)');
  Ago=replace_nameso(Ago,'Czech Republic','Czechia');
  Ago=replace_nameso(Ago,'Slovak Republic','Slovakia');
  Ago=replace_nameso(Ago,'Taiwan','Taiwan*');
  Ago=replace_nameso(Ago,'United States','US');

dt=Ac(1,5:end); % date
cl=country_list(Ac); % country list

%% further processing of data
disp('processing Oxford data');

indX=[40,48]; % selector of variables - check the file to make sure correct columns are selected
Data.Xnames={['Stringency Index']; ['Ec. Support Index']};
save_file_europe='panel_estimation_results_Europe.xlsx';

Data.Xnames={['Stringency Index']; ['Ec. Support Index']};
Ao=cell(size(Ac));
Ao(2:end,1)=cl;
Ao(1,5:end)=dt;
for i=1:size(cl,1)

   country_to_find=cl{i,1};
   [sp,foundc]=find_go_start(Ago,country_to_find);
   if foundc
    for j=1:size(dt,2)
        date_to_find=dt{1,j};
        [ox,~,foundd,sp1]= find_go_sp(Ago,country_to_find,date_to_find, sp, indX);
        for h=1:size(ox,2)
            Ao(i+1,j+4,h)=ox(1,h);
%            date_to_find
%            ox
        end
        if foundd
            sp=sp1;
        end
    end
   else
   % disp(['...country ',country_to_find,' not found in the Oxford database']);
   end
end

Ao=add_aggregatesWo(Ao, Ac, Ap);
Ac=add_aggregatesW(Ac, Ac);
Ap=add_aggregatesW(Ap, Ac);


%% computations
disp(['computing and saving results to Excel file ',save_file_europe,' ...'])

% data transformations
 % get number of countries/regions and time dimensions
n=size(Ac,1)-1; %-1 due to the first row being descriptions
T=size(Ac,2)-4; %-4 due to descriptions

cl=country_list(Ac);
pl=country_list(Ap);

C=cell2mat(Ac(2:end,5:end));
P=fill_entries(cell2mat(replace_br_with_nans(Ap(2:end,5:end))),pl,cl);

%now compute synthetic R series, assuming gamma
gamma=1/14; % do not change! There are several other instances in the codes that set gamma=1/14;
Rem2=createRe(C,gamma);

% compute number of active infections
I2=C-Rem2;
PT=P*ones(1,size(C,2));
s=(PT-C)./PT;

%compute c and i
c=C./PT;
i2=I2./PT;

% compute daily cases per 100k, 7dma
[dcpm, dCm]=compute_dc(C,PT);

matransform=true;W=14; % Set Moving Average transformation, and estimation window size
sv=50; % minimum threshold for computing cases
MF=2; [b_1wm_mf2, r_1wm_mf2]=compute_trans_rates_hc(MF*c, MF*i2,W,dCm, matransform, sv); 
MF=3; [b_1wm_mf3, r_1wm_mf3]=compute_trans_rates_hc(MF*c, MF*i2,W,dCm, matransform, sv); 
MF=2.5; [b_1wm_mf2_5, r_1wm_mf2_5]=compute_trans_rates_hc(MF*c, MF*i2,W,dCm, matransform, sv); 
MF=5; [b_1wm_mf5, r_1wm_mf5]=compute_trans_rates_hc(MF*c, MF*i2,W,dCm, matransform, sv);  
MF=7; [b_1wm_mf7, r_1wm_mf7]=compute_trans_rates_hc(MF*c, MF*i2,W,dCm, matransform, sv); 

cl=country_list(Ac); % country list
%% panel estimations

k=size(Ao,3);
for i=1:k
    Apom=Ao(2:end,5:end,i);
    Apom=replace_br_with_nans(Apom);
    X(:,:,i)=cell2mat(Apom);
end
MF=5; % Multiple factor
sv=100; % for selection of panel estimation sample

% define samples of countries
selected_countries={'Belgium';'France'; 'Germany';'Italy';'Netherlands';'Poland';'Portugal';...
    'Spain';'United Kingdom'};
selected_countries_short={'Bel';'Fra';'Ger';'Ita';'Neth';'Pol';'Por';...
    'Spain';'UK'};


Thvar.data=dcpm; 
Thvar.name=['Confirmed cases per day per 100k population (7day ma)'];
% define grid
gr(1:10,1)=[1:10]/1000;
gr(11:29,1)=0.01+[1:19]/100;
gr(30:37,1)=0.2+[1:8]/10;
gr(38:55,1)=1+[1:18]/2;
gr(56:80,1)=10+[1:25]*2;
Thvar.grid=gr;
Thvar.usegrid=true;

SC.list=selected_countries;
SC.short_names=selected_countries_short;
Data.c=c;
Data.i2=i2;
Data.I2=I2;
Data.X=X;
Data.Ac=Ac;
Data.b=b_1wm_mf5;
Data.R=r_1wm_mf5;
Data.dates=dt;

do_thresholds=true;
save_file='panel_estimation_results_Europe.xlsx';
panel_estimations(SC,Data, Thvar, sv, save_file,1, do_thresholds); 

disp('All done. Please check ReadMe.txt regarding the description of the results xlsx file.')
```
