## Supplementary material for "COVID-19 Time-varying Reproduction Numbers Worldwide: An Empirical Analysis of Mandatory and Voluntary Social Distancing": Replication package: select_AGGforCSave.m

```
function ycslag=select_AGGforCSave(ca,ia,Ia,selected_country, cslist, Ac, sv)

[c,i,I,ACsel]=select_sample_for_cs(ca,ia,Ia,selected_country,Ac);

s1=I>sv;
gamma=1/14;
y=NaN(size(c));
y(:,1:end-1)=-log((1-c(:,2:end))./(1-c(:,1:end-1)))./i(:,1:end-1)/gamma;
sy=not(isnan(y));
s=s1&sy;

% compute CS ave for countries in cslist, excluding the domestic economy
n=size(cslist,1);
T=size(c,2);

ycs=NaN(n,T);
ycslag=NaN(n,T);

na=size(y,1);

ys=y;
ys(:,not(s))=NaN;

for ii=1:n
   ycs(ii,:)=ys;
end
% lag by one week

ycslag(:,8:end)=ycs(:,1:end-7);


return
```
