## Supplementary material for "COVID-19 Time-varying Reproduction Numbers Worldwide: An Empirical Analysis of Mandatory and Voluntary Social Distancing": Replication package: select_sample.m

```
function [csel,isel,Isel,Xsel,bsel,ACsel, Thsel]=select_sample(c,ii,I,X,b,selected_countries,Ac, Th)
cl=country_list(Ac);
nc=size(cl,1);
ind=false(nc,1);
ind1=false(nc+1,1); ind1(1,1)=true;
ns=size(selected_countries,1);

csel=zeros(ns,size(c,2));
bsel=zeros(ns,size(c,2));
isel=zeros(ns,size(ii,2));
Isel=zeros(ns,size(I,2));
ACsel=Ac(1,:);
Xsel=zeros(ns,size(X,2),size(X,3));

% create moving aver for Threg
W=1;
Thma=NaN(size(Th));
    for t=W:size(Th,2)
        Thma(:,t)=Th(:,t-W+1:t)*ones(W,1)/W;
    end
Thsel=zeros(0,size(Thma,2));    
k=size(X,3);    

for j=1:ns
    for i=1:nc
        if strcmp(cl{i,1},selected_countries{j,1}) 
            ind(i,1)=true;
            ind1(i+1,1)=true;
            % add country to the set of selected data
                csel(j,:)=c(i,:);
                bsel(j,:)=b(i,:);
                isel(j,:)=ii(i,:);
                Isel(j,:)=I(i,:);
                ACsel(j+1,:)=Ac(i+1,:);
                for jj=1:k
                    Xsel(j,:,jj)=X(i,:,jj);
                end
                Thsel(j,:)=Thma(i,:);   
        end
    end
end

% csel=c(ind,:);
% isel=ii(ind,:);
% Isel=I(ind,:);
% ACsel=Ac(ind1,:);
% k=size(X,3);
% for j=1:k
%     Xsel(:,:,j)=X(ind,:,j);
% end


%Thsel=Thma(ind,:);    
    
return
```
