## Supplementary material for "COVID-19 Time-varying Reproduction Numbers Worldwide: An Empirical Analysis of Mandatory and Voluntary Social Distancing": Replication package: select_sample_for_cs.m

```
function [csel,isel,Isel,ACsel]=select_sample_for_cs(c,ii,I,selected_countries,Ac)
cl=country_list(Ac);
nc=size(cl,1);
ind=false(nc,1);
ind1=false(nc+1,1); ind1(1,1)=true;
ns=size(selected_countries,1);

for j=1:ns
    for i=1:nc
        if strcmp(cl{i,1},selected_countries{j,1}) 
            ind(i,1)=true;
            ind1(i+1,1)=true;
        end
    end
end

csel=c(ind,:);
isel=ii(ind,:);
Isel=I(ind,:);
ACsel=Ac(ind1,:);

    
return
```
