## Supplementary material for "COVID-19 Time-varying Reproduction Numbers Worldwide: An Empirical Analysis of Mandatory and Voluntary Social Distancing": Replication package: SubSaharan_Africa.m

```
function [list]=SubSaharan_Africa()

% west and central ... eastern and southern
list={'Benin';'Burkina Faso';'Cabo Verde';'Cameroon';'Central African Republic';'Chad';...
   'Congo (Brazzaville)';'Congo (Kinshasa)';'Cote d''Ivoire';'Croatia';'Equatorial Guinea';'Gabon';'Gambia';...
  'Ghana';'Guinea';'Guinea-Bissau';'Liberia';'Mali';'Mauritania';'Niger';'Nigeria';'Sao Tome and Principe';...
  'Senegal';'Sierra Leone';'Togo';...
  'Angola';'Botswana';'Burundi';'Comoros';'Djibouti';'Eritrea';'Ethiopia';...
    'Kenya';'Lesotho';'Madagascar';'Malawi';'Mozambique';'Namibia';'Rwanda';'Seychelles';'Somalia';...
    'South Africa';'South Sudan';'Sudan';'Uganda';'Tanzania''Zambia';'Zimbabwe'};


return
```
