## Supplementary material for "COVID-19 Time-varying Reproduction Numbers Worldwide: An Empirical Analysis of Mandatory and Voluntary Social Distancing": Replication package: Western_Europe.m

```
function [list]=Western_Europe()

list={'Andorra';'Austria';'Belgium';'Cyprus';'Czechia';'Denmark';'Estonia';'Finland';'France';...
    'Germany';'Greece';'Holy See';'Hungary';'Iceland';'Ireland';'Italy';'Latvia';'Liechtenstein';...
    'Lithuania';'Luxembourg';'Malta';'Monaco';'Netherlands';'Norway';'Poland';'Portugal';'San Marino';...
   'Slovakia';'Slovenia';'Spain';'Sweden';'Switzerland';'United Kingdom'};


return
```
