## Supplementary material for "COVID-19 Time-varying Reproduction Numbers Worldwide: An Empirical Analysis of Mandatory and Voluntary Social Distancing": Replication package: world_list.m

```
function [worldlist, worldlist_exChina]=world_list()

worldlist={'Afghanistan';'Albania';'Algeria';'Andorra';'Angola';'Antigua and Barbuda';'Argentina';...
    'Armenia';'Austria';'Azerbaijan';'Bahamas';'Bahrain';'Bangladesh';'Barbados';'Belarus';'Belgium';...
    'Belize';'Benin';'Bhutan';'Bolivia';'Bosnia and Herzegovina';'Botswana';'Brazil';'Brunei';'Bulgaria';...
    'Burkina Faso';'Burma';'Burundi';'Cabo Verde';'Cambodia';'Cameroon';'Central African Republic';...
    'Chad';'Chile';'Colombia';'Comoros';'Congo (Brazzaville)';'Congo (Kinshasa)';'Costa Rica';...
    'Cote d''Ivoire';'Croatia';'Cuba';'Cyprus';'Czechia';'Denmark';'Djibouti';'Dominica';...
    'Dominican Republic';'Ecuador';'Egypt';'El Salvador';'Equatorial Guinea';'Eritrea';'Estonia';...
    'Eswatini';'Ethiopia';'Fiji';'Finland';'France';'Gabon';'Gambia';'Georgia';'Germany';'Ghana';...
    'Greece';'Grenada';'Guatemala';'Guinea';'Guinea-Bissau';'Guyana';'Haiti';'Holy See';'Honduras';...
    'Hungary';'Iceland';'India';'Indonesia';'Iran';'Iraq';'Ireland';'Israel';'Italy';'Jamaica';'Japan';...
    'Jordan';'Kazakhstan';'Kenya';'Korea, South';'Kosovo';'Kuwait';'Kyrgyzstan';'Laos';'Latvia';'Lebanon';...
    'Lesotho';'Liberia';'Libya';'Liechtenstein';'Lithuania';'Luxembourg';'MS Zaandam';'Madagascar';'Malawi';...
    'Malaysia';'Maldives';'Mali';'Malta';'Marshall Islands';'Mauritania';'Mauritius';'Mexico';'Moldova';...
    'Monaco';'Mongolia';'Montenegro';'Morocco';'Mozambique';'Namibia';'Nepal';'Netherlands';'New Zealand';...
    'Nicaragua';'Niger';'Nigeria';'North Macedonia';'Norway';'Oman';'Pakistan';'Panama';'Papua New Guinea';...
    'Paraguay';'Peru';'Philippines';'Poland';'Portugal';'Qatar';'Romania';'Russia';'Rwanda';...
    'Saint Kitts and Nevis';'Saint Lucia';'Saint Vincent and the Grenadines';'Samoa';...
    'San Marino';'Sao Tome and Principe';'Saudi Arabia';'Senegal';'Serbia';'Seychelles';'Sierra Leone';...
    'Singapore';'Slovakia';'Slovenia';'Solomon Islands';'Somalia';'South Africa';'South Sudan';'Spain';...
    'Sri Lanka';'Sudan';'Suriname';'Sweden';'Switzerland';'Syria';'Taiwan*';'Tajikistan';'Tanzania';'Thailand';...
    'Timor-Leste';'Togo';'Trinidad and Tobago';'Tunisia';'Turkey';'US';'Uganda';'Ukraine';'United Arab Emirates';...
    'United Kingdom';'Uruguay';'Uzbekistan';'Vanuatu';'Venezuela';'Vietnam';'West Bank and Gaza';'Yemen';'Zambia';...
    'Zimbabwe';'Canada';'China';'Australia'};

worldlist_exChina={'Afghanistan';'Albania';'Algeria';'Andorra';'Angola';'Antigua and Barbuda';'Argentina';...
    'Armenia';'Austria';'Azerbaijan';'Bahamas';'Bahrain';'Bangladesh';'Barbados';'Belarus';'Belgium';...
    'Belize';'Benin';'Bhutan';'Bolivia';'Bosnia and Herzegovina';'Botswana';'Brazil';'Brunei';'Bulgaria';...
    'Burkina Faso';'Burma';'Burundi';'Cabo Verde';'Cambodia';'Cameroon';'Central African Republic';...
    'Chad';'Chile';'Colombia';'Comoros';'Congo (Brazzaville)';'Congo (Kinshasa)';'Costa Rica';...
    'Cote d''Ivoire';'Croatia';'Cuba';'Cyprus';'Czechia';'Denmark';'Djibouti';'Dominica';...
    'Dominican Republic';'Ecuador';'Egypt';'El Salvador';'Equatorial Guinea';'Eritrea';'Estonia';...
    'Eswatini';'Ethiopia';'Fiji';'Finland';'France';'Gabon';'Gambia';'Georgia';'Germany';'Ghana';...
    'Greece';'Grenada';'Guatemala';'Guinea';'Guinea-Bissau';'Guyana';'Haiti';'Holy See';'Honduras';...
    'Hungary';'Iceland';'India';'Indonesia';'Iran';'Iraq';'Ireland';'Israel';'Italy';'Jamaica';'Japan';...
    'Jordan';'Kazakhstan';'Kenya';'Korea, South';'Kosovo';'Kuwait';'Kyrgyzstan';'Laos';'Latvia';'Lebanon';...
    'Lesotho';'Liberia';'Libya';'Liechtenstein';'Lithuania';'Luxembourg';'MS Zaandam';'Madagascar';'Malawi';...
    'Malaysia';'Maldives';'Mali';'Malta';'Marshall Islands';'Mauritania';'Mauritius';'Mexico';'Moldova';...
    'Monaco';'Mongolia';'Montenegro';'Morocco';'Mozambique';'Namibia';'Nepal';'Netherlands';'New Zealand';...
    'Nicaragua';'Niger';'Nigeria';'North Macedonia';'Norway';'Oman';'Pakistan';'Panama';'Papua New Guinea';...
    'Paraguay';'Peru';'Philippines';'Poland';'Portugal';'Qatar';'Romania';'Russia';'Rwanda';...
    'Saint Kitts and Nevis';'Saint Lucia';'Saint Vincent and the Grenadines';'Samoa';...
    'San Marino';'Sao Tome and Principe';'Saudi Arabia';'Senegal';'Serbia';'Seychelles';'Sierra Leone';...
    'Singapore';'Slovakia';'Slovenia';'Solomon Islands';'Somalia';'South Africa';'South Sudan';'Spain';...
    'Sri Lanka';'Sudan';'Suriname';'Sweden';'Switzerland';'Syria';'Taiwan*';'Tajikistan';'Tanzania';'Thailand';...
    'Timor-Leste';'Togo';'Trinidad and Tobago';'Tunisia';'Turkey';'US';'Uganda';'Ukraine';'United Arab Emirates';...
    'United Kingdom';'Uruguay';'Uzbekistan';'Vanuatu';'Venezuela';'Vietnam';'West Bank and Gaza';'Yemen';'Zambia';...
    'Zimbabwe';'Canada';'Australia'};

return
```
