## Supplementary material for "COVID-19 Time-varying Reproduction Numbers Worldwide: An Empirical Analysis of Mandatory and Voluntary Social Distancing": Replication package: ReadMe.txt

This subfolder contains codes for the estimation of R numbers and transmission rates for individual countries with available case data as reported by JHU data hub. 

The script to run is: "RunMe.m". This file contains comments that will help user. 

There are two options how to run this script:

(i) When "download=1" on line 10 (default option), this script will download the latest data from John Hopkins University and automatically save the data in the local file confirmed_cases_txt.
(ii) When "download=0" on line 10, the script will not download data from the web and it will proceed with loading localy saved data from file confirmed_cases_txt. 

In addition to COVID-19 case data, script also loads population data from Excel file "population.xlsx".

The outcome of the estimations is saved in Excel file "Estimates.xlsx". This file contains the following sheets:
- Sheets "R_mfX" (red), where X stands for different values of multiplication factor considered, namely X=2, 2.5, 3, 5, and 7 is reported. These sheets report the estimated R values for individual countries and given multiplication factor. First row contains dates.
- Sheets "bg_mfX" (blue), where X stands for different values of multiplication factor considered, namely X=2, 2.5, 3, 5, and 7 is again reported. These sheets contain the estimates of effective transmsission rate beta x 14 (in which 14=1/gamma).
- Sheet "dc" (yellow) contains 7-day moving average of new cases per 100k population. 

Estimates of R numbers and effective transmission rates are subject to the chosen upper bound set equal to 3.
