## Supplementary material for "COVID-19 Time-varying Reproduction Numbers Worldwide: An Empirical Analysis of Mandatory and Voluntary Social Distancing": Replication package: run_me.m

```
%% This program estimates R numbers and transmistion rates using the methodology in the paper:
% "COVID-19 Time-varying Reproduction Numbers Worldwide: An Empirical Analysis of Mandatory and Voluntary Social Distancing,"
% by A. Chudik, M. H. Pesaran and A. Rebucci, March 15, 2021
%
% Program below can either download the latest data from John Hopkins University (JHU), or use the saved data (to be selected by user on line 10 below)
% The outcome of the estimations is saved in Excel file "Estimates.xlsx". See ReadMe.txt on the information about this file.

clc 
clear
download=1; % Set to 1 to download the latest data from JHU. This option will overwrite local confirmed_cases_txt data file with the latest COVID-19 case data. 
            % Set to 0 to download data from previously saved local file confirmed_cases_txt.

%% load data            

files_load=1; % do not change this

if download==1
  %download case count data and save them from internet
  disp('downloading COVID-19 data from internet...')
  fileC='https://github.com/CSSEGISandData/COVID-19/raw/master/csse_covid_19_data/csse_covid_19_time_series/time_series_covid19_confirmed_global.csv';
  FC=urlread(fileC);
 
  %save into csv files
  disp('saving downloaded data into local file...');  
  fileID = fopen('confirmed_cases_txt','w');fprintf(fileID,FC);fclose(fileID);
  clear FC ;
end %download

 %read data from files in matlab cellarray
if files_load==1
    disp('loading COVID-19 data from local txt files...')
  % covid-19 data   
  Ac = readcell('confirmed_cases_txt', 'DatetimeType','text');
  % population data
  disp('loading population data from xlsx file...')
  Ap = readcell('population.xlsx');
  % delete missing population entries
  j=2;
  for i=2:size(Ap,1)
      if ismissing(Ap{j,5})
          Ap(j,:)=[];
      else
          j=j+1;
      end
  end
  Ap=reorderAP(Ap,Ac); % make sure order is identical;

  Ac=correct_french_data(Ac); % if executed, it will correct the obvious data error in JHU French data in the period 4/4 to 4/11. This might no longer be relevant in the latest data vintage. 
  
  % add not reported Canada, China and Australia country aggregates to Ac
  Ac=add_aggregates(Ac);
  % add the same aggregates to Ap
  Ap=add_aggregates(Ap);
end

dt=Ac(1,5:end); % date

% add global region aggregates
Ac=add_aggregatesW(Ac, Ac);
Ap=add_aggregatesW(Ap, Ac);


%% computations
disp('computing ... please wait a minute...')


% data transformations
 % get number of countries/regions and time dimensions
n=size(Ac,1)-1; %-1 due to the first row being descriptions
T=size(Ac,2)-4; %-4 due to descriptions

cl=country_list(Ac);
pl=country_list(Ap);

C=cell2mat(Ac(2:end,5:end));
P=fill_entries(cell2mat(replace_br_with_nans(Ap(2:end,5:end))),pl,cl);

%now compute synthetic R series, assuming gamma
gamma=1/14; % do not change! There are several other instances in the codes that set gamma=1/14;
Rem2=createRe(C,gamma);

% compute number of active infections
I2=C-Rem2;
PT=P*ones(1,size(C,2));
s=(PT-C)./PT;

%compute c and i
c=C./PT;
i2=I2./PT;

% compute daily cases per 100k, 7dma
[dcpm, dCm]=compute_dc(C,PT);

matransform=true; W=14; % Set Moving Average transformation, and estimation window size
sv=50; % minimum threshold for computing cases
MF=2; [b_1wm_mf2, r_1wm_mf2]=compute_trans_rates_hc(MF*c, MF*i2,W,dCm, matransform, sv); 
MF=3; [b_1wm_mf3, r_1wm_mf3]=compute_trans_rates_hc(MF*c, MF*i2,W,dCm, matransform, sv); 
MF=2.5; [b_1wm_mf2_5, r_1wm_mf2_5]=compute_trans_rates_hc(MF*c, MF*i2,W,dCm, matransform, sv); 
MF=5; [b_1wm_mf5, r_1wm_mf5]=compute_trans_rates_hc(MF*c, MF*i2,W,dCm, matransform, sv);  
MF=7; [b_1wm_mf7, r_1wm_mf7]=compute_trans_rates_hc(MF*c, MF*i2,W,dCm, matransform, sv); 


%% save results
disp('saving estimation results to Excel file...')
filenameF='Estimates.xlsx';

A=cell(n+1,T+1);          
A(2:end,1)=cl;
A(1,2:T+1)=dt;

A(2:end,2:end)=num2cell(r_1wm_mf5);
writecell(A,filenameF, 'Sheet', 'R_mf5', 'Range', 'A1');
A(2:end,2:end)=num2cell(r_1wm_mf2_5);
writecell(A,filenameF, 'Sheet', 'R_mf2_5', 'Range', 'A1');
A(2:end,2:end)=num2cell(r_1wm_mf7);
writecell(A,filenameF, 'Sheet', 'R_mf7', 'Range', 'A1');
A(2:end,2:end)=num2cell(r_1wm_mf3);
writecell(A,filenameF, 'Sheet', 'R_mf3', 'Range', 'A1');
A(2:end,2:end)=num2cell(r_1wm_mf2);
writecell(A,filenameF, 'Sheet', 'R_mf2', 'Range', 'A1');


A(2:end,2:end)=num2cell(dcpm);
writecell(A,filenameF, 'Sheet', 'dc', 'Range', 'A1');

A(2:end,2:end)=num2cell(b_1wm_mf5/gamma);
writecell(A,filenameF, 'Sheet', 'bg_mf5', 'Range', 'A1');
A(2:end,2:end)=num2cell(b_1wm_mf2_5/gamma);
writecell(A,filenameF, 'Sheet', 'bg_mf2_5', 'Range', 'A1');
A(2:end,2:end)=num2cell(b_1wm_mf7/gamma);
writecell(A,filenameF, 'Sheet', 'bg_mf7', 'Range', 'A1');
A(2:end,2:end)=num2cell(b_1wm_mf3/gamma);
writecell(A,filenameF, 'Sheet', 'bg_mf3', 'Range', 'A1');
A(2:end,2:end)=num2cell(b_1wm_mf2/gamma);
writecell(A,filenameF, 'Sheet', 'bg_mf2', 'Range', 'A1');

disp(['All done. Results were saved to ',filenameF,'. Check ReadMe.txt for additional info.'])
```
