## Supplementary material for "COVID-19 Time-varying Reproduction Numbers Worldwide: An Empirical Analysis of Mandatory and Voluntary Social Distancing": Replication package: ReadMe.txt

This folder contains codes and data for paper entitled "COVID-19 Time-varying Reproduction Numbers Worldwide: An Empirical Analysis
of Mandatory and Voluntary Social Distancing," by A. Chudik, M. H. Pesaran and A. Rebucci, 15 March 2021. It is organized in two self-contained separate subfolders:

- Subfolder "R_estimates" is for the estimation of R numbers (and transmission rates) reported in Section 4 of the paper.
- Subfolder "European_Panel_Estimates" is for the replication of the panel regressions results reported in Section 5 of the paper.

Each subfolder contains its "ReadMe.txt" file, where it is explained how to run the codes.
